# Impact of Intended Isocaloric Early vs. Late Time-Restricted Eating on Plasma Lipidome in Women with Overweight or Obesity: Secondary Analysis of the ChronoFast Trial

**DOI:** 10.1101/2025.04.01.25324944

**Authors:** Kristof Szekely, Mathias J. Gerl, Beeke Peters, Julia Schwarz, Bettina Schuppelius, Markus Damm, Jorge R. Soliz-Rueda, Ratika Sehgal, Michail Lazaratos, Christian Klose, Kai Simons, Andreas F. H. Pfeiffer, Annette Schürmann, Achim Kramer, Andreas Michalsen, Olga Pivovarova-Ramich

**Author notes:** **Correspondence:** Olga Pivovarova-Ramich, German Institute of Human Nutrition Potsdam-Rehbruecke, Arthur-Scheunert-Allee 114-116, 14558 Nuthetal, Germany.

## Abstract

Time-restricted eating (TRE) is a promising strategy to prevent obesity and type 2 diabetes, but its effects on lipid metabolism remain controversial. The aim of the present research is to assess and compare the impact of isocaloric early (eTRE) vs. late (lTRE) TRE on the plasma lipidomic profile. This randomized crossover study examines 31 women with overweight or obesity who follow a two-week eTRE and a two-week lTRE in an intended isocaloric setting. Blood plasma and subcutaneous adipose tissue biopsies are analyzed using shotgun lipidomics and transcriptomics, respectively. Between interventions and within the lTRE, lipid species and classes, as well as enzyme activity indices, are not substantially changed. Within the eTRE, changes are observed for 103 lipid species, including a reduction of ceramide and phosphatidylcholine classes, and for the desaturation indices D5D, D6D, and D9D, as well as elongation index ELOVL6. Combined analysis of plasma lipidome and adipose tissue reveals alterations in the glycerophospholipid pathway and in the expression of phospholipase enzymes PLB1, PLA2G6, and PLAG4B, dependent on TRE timing. These results suggest that eating timing during TRE might be crucial for remodeling the plasma lipidome and adipose tissue transcriptome and highlight the need of future lipidomic research in TRE.

## 1. Introduction

In recent years, intermittent fasting (IF) has become increasingly popular within the scientific community and beyond. One type of IF is time-restricted eating (TRE), in which food intake is limited to 6 to 10 hours per day. The reason for the popularity of this approach, in addition to its numerous beneficial effects, is an easy implementation.^[1]^ The effects of TRE have been intensively studied in recent years, and improvements have been observed for body weight^[2–17]^, body fat^[3, 4, 7, 8, 10, 12, 13, 15, 16]^, waist circumference^[9, 10, 15, 18]^, inflammatory markers^[8, 16]^, oxidative stress^[7, 19]^, blood pressure^[6, 10, 12, 19]^, blood glucose parameters^[3, 7, 8, 11, 14, 19, 20]^, sleep, and overall quality of life.^[2, 9]^ Therefore, TRE is a promising prevention and treatment approach improving the metabolic state in obesity and type 2 diabetes. In these diseases, dysfunctions of carbohydrate and lipid metabolism are closely interrelated. In particular, dyslipidemia has been described in type 2 diabetes mellitus.^[21]^ However, the effects of TRE on lipid metabolism are partly controversial.

Several TRE trials have found improvements in triglyceride^[3, 4, 8, 14, 22]^, total and low-density-lipoprotein (LDL) cholesterol levels.^[10, 22]^ Nevertheless, these results vary depending on the study design and study cohort, with some TRE studies showing no change or even a worsening of lipid metabolism.^[5–7, 9–13, 19, 20, 23]^ Further, it remains generally unclear whether shortening of the eating window provides additional metabolic effects compared to continuous caloric restriction and/or changes in dietary composition, and which timing of eating during TRE is more beneficial. Remarkably, most clinical studies suggest metabolic benefits of eTRE ^[11, 19, 20, 24]^, which are hypothesized to be superior compared to the lTRE. ^[25]^ However, trials directly comparing eTRE and lTRE are very limited ^[14, 26–28]^, and their results are also inconsistent. While some trials found a decrease in triglycerides and LDL cholesterol in both interventions, showing no difference between the early and late eating windows ^[14, 28]^, other studies observed an aggravation of LDL cholesterol within both diets ^[27]^.

The development of novel research techniques, such as high throughput shotgun lipidomics, offers a significantly more accurate approach to assess a broader range of plasma lipids, opening new horizons in lipid research compared to lipid parameters used in everyday clinical practice.^[29]^ In particular, the lipidome analysis can provide more insights into the mechanisms of TRE. Using this method, a reduction in sphingosines and different sphingomyelins was observed after some IF interventions, e.g. the Ramadan fast.^[30]^ In our previous research, shotgun lipidomic analysis allowed to characterize in detail the daytime-dependent changes in postprandial lipid metabolism^[31]^ and confirmed that the timing of meal intake modulates dietary-induced lipid responses. However, so far, no lipidomic study has directly compared the effects of early and late TRE, especially in a crossover trial. Therefore, the aim of the present research was to assess and compare the impact of isocaloric eTRE vs. late lTRE on the plasma lipidomic profile. Based on previous literature, we expected that eTRE would induce more pronounced effects on plasma lipidome compared to the lTRE. The study was a secondary analysis of the crossover ChronoFast trial (NCT04351672) conducted in women with overweight or obesity.^[32]^ Because an additional aim was to assess the contribution of adipose tissue in lipid changes upon TRE, we combined the analysis of plasma lipidome with gene expression profiling in the subcutaneous adipose tissue (SAT).

## 2. Results

### 2.1. Study Population and Adherence to Dietary Interventions

31 female subjects with overweight or obesity (age: 62 (53-65) years; BMI: 30.5 (2.9) kg/m^2^) - 13 with impaired fasting glucose and/or impaired glucose tolerance (IFG/IGT) and 18 with normal glucose tolerance (NGT) - completed this study. 26 participants were postmenopausal, and 5 were premenopausal. Five participants has taken lipid-lowering medication, which was continued during the whole study period. Baseline characteristics of study participants are shown in **Table 1**.

**Table 1.**
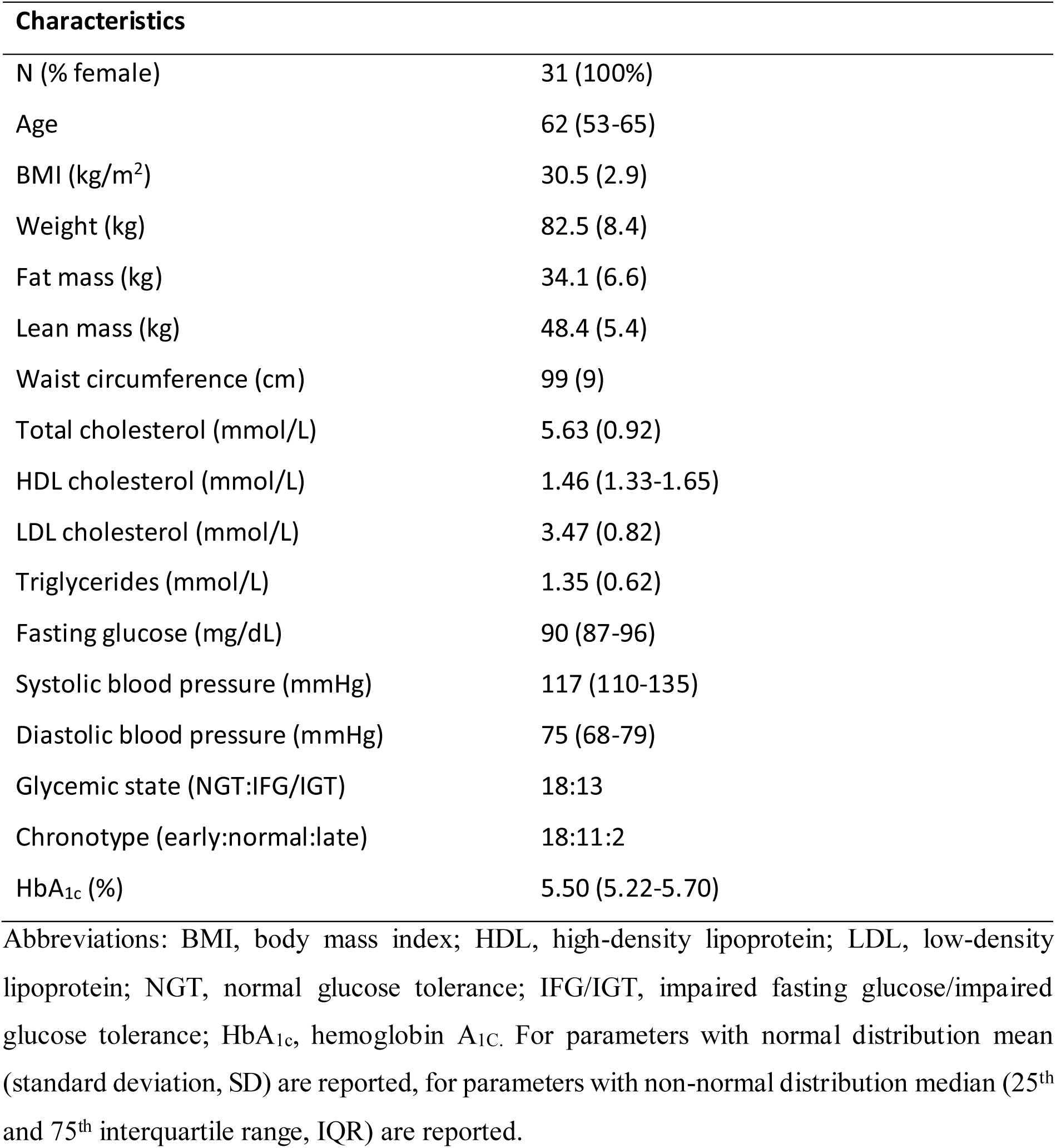
Baseline characteristics of study participants.

Details of this trial are described in **Figure 1A**, **Table S1**, the **Experimental section/Methods**, and Peters et al.^[32, 33]^ In brief, after a two-to-four-week baseline period, participants followed a two-week eTRE and a two-week lTRE intervention in a crossover design. They showed a high adherence to the 8-hour eating window in both TRE interventions. During the baseline phase, the participants’ median eating window was 11:48 (10:50 - 13:15) h per day, which decreased to 7:09 (6:57 - 7:21) h during the eTRE intervention, and to 6:57 (6:39 - 7:16) h during the lTRE intervention. The subjects maintained a high timely compliance of 96.5 % (6.3 %) in eTRE and 97.7 % (6.1 %) in lTRE. There was no difference in macronutrient composition and physical activity between and within the two TRE interventions ( **Table S2**). Despite intensive real-time dietary monitoring to ensure an isocaloric intake ^[33]^, a minor, but statistically significant, decrease in daily energy intake was observed within the eTRE intervention (−167 kcal; *P* < 0.001), but not within lTRE (−97 kcal; *P* = 0.06). Minimal weight loss was observed within both eTRE (−1.08 kg; *P* < 0.001) and lTRE (−0.44 kg; *P* = 0.01), with a between-intervention difference of 0.65 kg (*P* = 0.012). Fat mass loss (−0.61 kg, *P* = 0.002) and lean mass loss (−0.57 kg; *P* = 0.04) were found within eTRE only, but their percentage to body weight was not changed within and between both interventions (**Table S3**). Total cholesterol, LDL cholesterol, and triglyceride concentrations were not affected by both interventions. High-density lipoprotein (HDL) cholesterol declined within both eTRE (−0.10 mmol/L; *P* <0.001) and lTRE (−0.07 mmol/L; *P* = 0.003), with no difference between the interventions. Body weight, body composition, and clinical lipid parameters were comparable at the beginning of eTRE and lTRE (**Table S3**).

**Figure 1.**
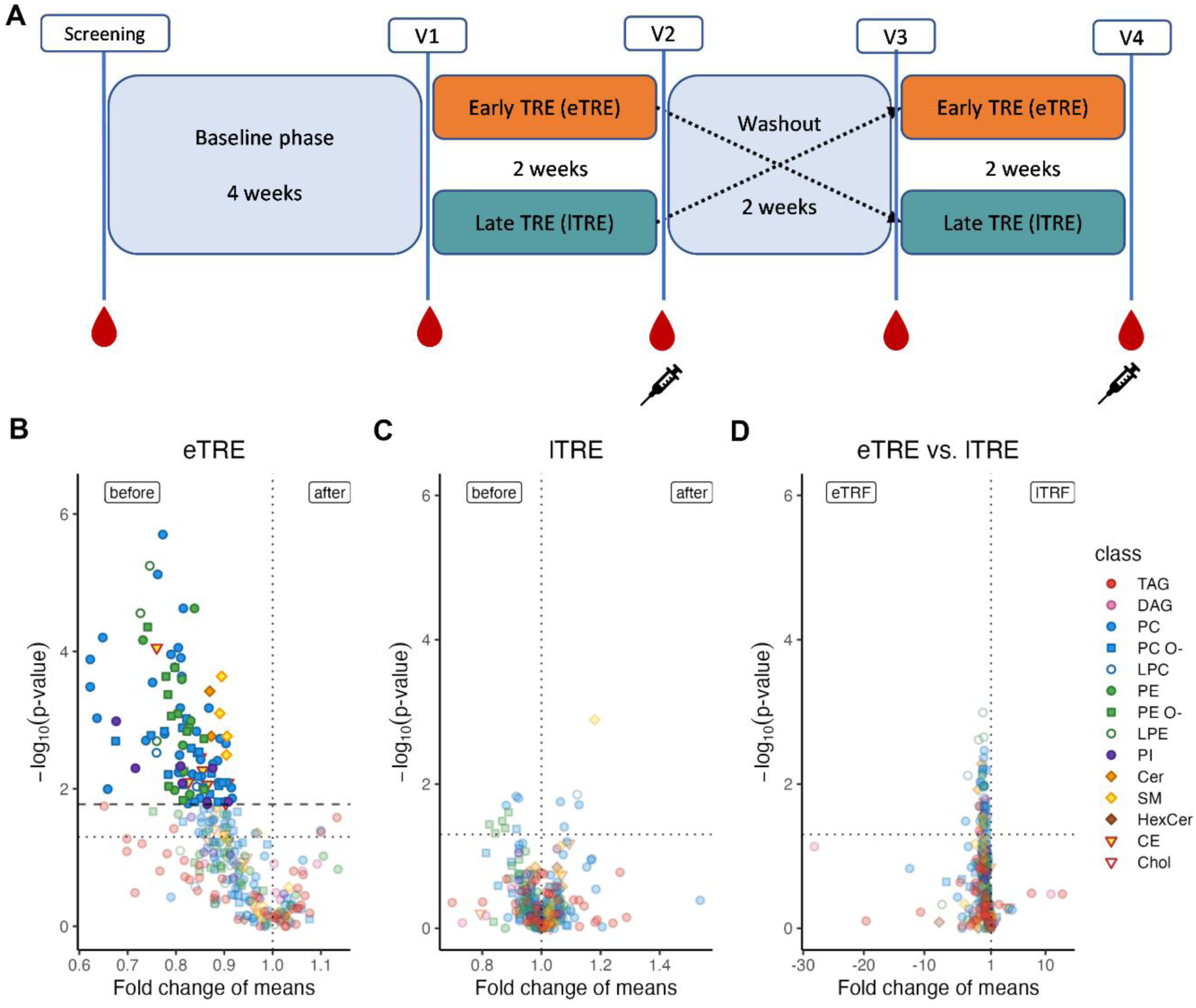
Impact of eTRE and lTRE on lipid species profiles in plasma. A) Study design. Early TRE (eTRE, orange), eating window between 8 a.m. and 4 p.m.; late TRE (lTRE, petrol), eating window between 1 pm and 9 pm; red drops indicate the collection of blood samples for lipidomic analysis and syringes indicate the SAT samples; V, visit. B) Changes in lipid species profiles within the eTRE intervention. C) Changes in lipid species profiles within the lTRE intervention. D) Changes in lipid species profiles between eTRE and lTRE intervention. In (B) and (C): Volcano plots show comparisons of lipid species before vs. after the intervention (N=30 for eTRE and N=31 for lTRE). (D): Volcano plot show comparisons of lipid species between the two interventions (eTRE vs. lTRE). Only lipid species with p <0.05 in the paired Mann-Whitney U Test (before correction for multiple testing) are shown. The y-axis displays the negative logarithm of p-values (uncorrected), and the x-axis shows fold changes of mean amounts. A dotted vertical line at a fold change of 1 indicates no change in lipid abundance. Titles indicate the intervention, and the labels at the top of each plot show the direction of fold change: ‘before vs. after’ for (B) and (C), and ‘eTRE vs. lTRE’ for (D). Lipid (sub-)species not significant after Benjamini-Hochberg correction are shown translucent; significant species are shown opaque with an outline. In (B), the horizontal line indicates the highest p-value significant after Benjamini-Hochberg correction. In (C) and (D), no significance threshold could be established. All plots include a dotted horizontal line at p-value = 0.05 (uncorrected) as a visual reference.

### 2.2. Detected Plasma Lipids

Lipid profiles in plasma samples could be determined in 30 subjects for the eTRE intervention and in 31 subjects for the lTRE intervention. Lipidomics analysis yielded on average 10534 ± 2768 pmol (*n* = 154) of lipids per μL of sample. Lipid species from 14 lipid classes (cholesterol (Chol), cholesterol esters (CE), triacylglycerols (TAG), diacylglycerols (DAG), phosphatidylcholines (PCs), phosphatidylcholine ethers (PC O-), phosphatidylethanolamines (PE), phosphatidylethanolamine ethers (PE O-), phosphatidylinositols (PI), lysophosphatidylcholines (LPC), lysophosphatidylcholine ethers (LPC O-), lysophosphatidylethanolamines (LPE), sphingomyelins (SM), and ceramides (CER) were identified and quantified **(Table S4)**. Lipid species present in less than 70% of all samples were excluded, resulting in a total of 300 lipid species for further analysis. This procedure accounted for 99% (98.9 ± 0.6 (*n* = 154)) of the total lipid content.

Plasma lipid patterns were analyzed for changes of (i) lipid species, (ii) lipid classes, (iii) fatty acids; (iv) indices of desaturase and elongase activities; (v) double bonds; (vi) total carbon lengths –compared effects of both interventions and within the eTRE and lTRE.

### 2.3. Effects of eTRE and lTRE on Plasma Lipid Species and Classes

Between eTRE and lTRE interventions, no alterations of lipid species were revealed (here and elsewhere: only changes of lipid parameters which remained significant after the correction for multiple testing, are described) **(Figure 1B-D, Table S5)**. Despite the lack of between-group differences, we conducted an additional exploratory analysis of the within-intervention changes. Within the eTRE, a decrease in 103 lipid species compared to the beginning of the intervention was found, while no lipid species were altered within the lTRE **(Figure 1B, Table S5)**.

Analysis of lipid classes revealed no alterations between interventions and within the lTRE **(Figure 2A, Table S6)**. Within the eTRE intervention, a reduction of ceramides (*P* = 0.043) and phosphatidylcholines (*P* = 0.043) was observed **(Figure 2A, Table S6)**.

**Figure 2.**
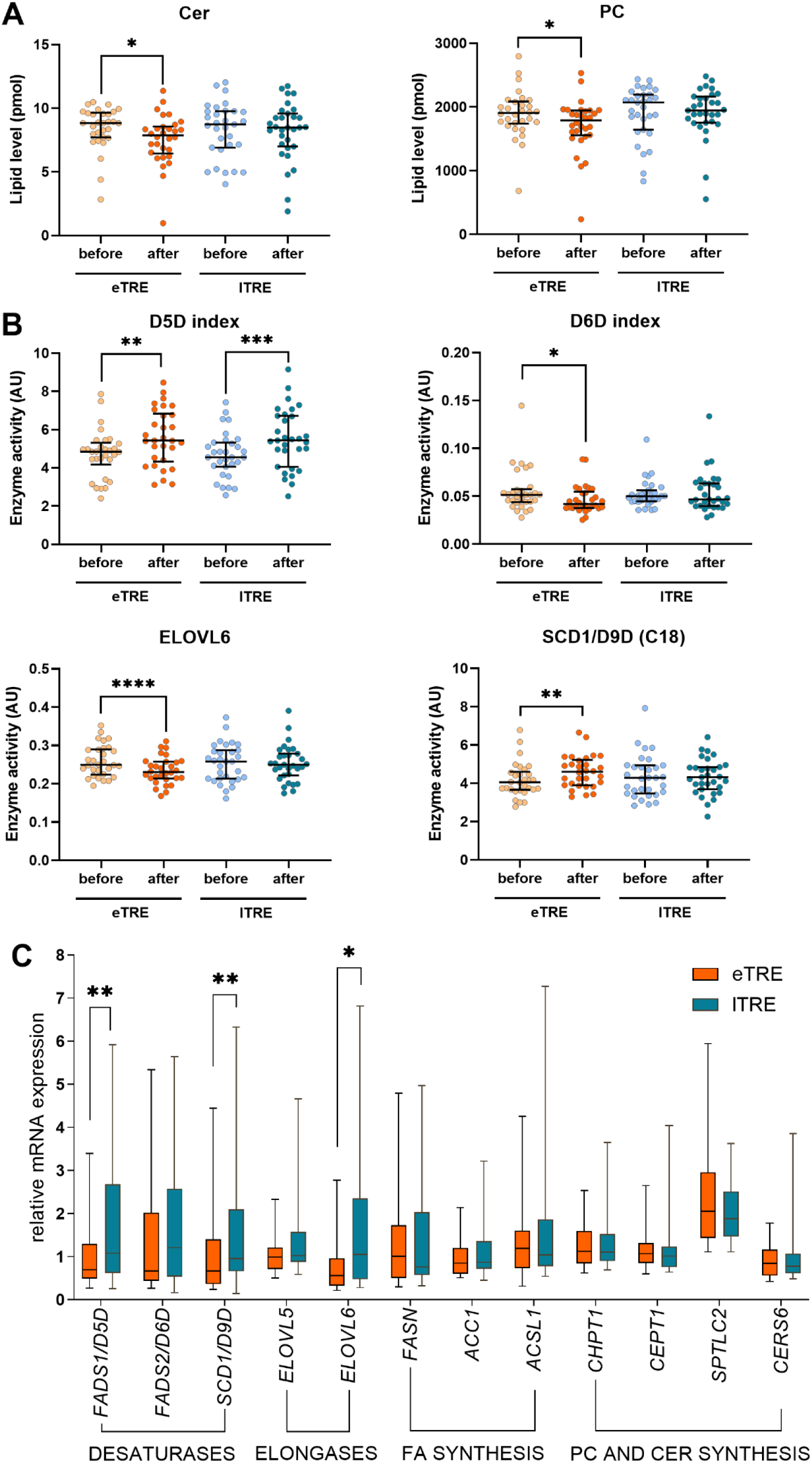
Lipid classes and enzyme activity indices in plasma and gene expression in adipose tissue. A) Lipid classes showed significant alterations within eTRE (lipid levels before and after the intervention are shown with beige and orange dots, respectively) or lTRE (lipid levels before and after the intervention are shown with blue and petrol dots, respectively) intervention. B) Activity indices of key lipid metabolism enzymes showed significant alterations within eTRE (beige/orange) or lTRE (blue/petrol) intervention. In (A) and (B): N=30 for eTRE and N=31 for lTRE; data are shown as dot plots with median and IQR. The data were analyzed using the Wilcoxon signed-rank test. The Benjamini-Hochberg method was used to control for FDR. Only lipid indices with significant differences after adjusting for multiple testing are shown. Significance levels are encoded as follows: *p < 0.05, **p < 0.01, ***p < 0.001. C) Expression of key lipid enzymes in subcutaneous adipose tissue (SAT). Relative mRNA expression of the desaturases *FADS1/D5D, FADS2/D6D, SCD1/D9D*; elongases *ELOVL5, ELOVL6*; FA synthesis enzymes *FASN, ACC1, ACSL1*; PC and CER synthesis enzymes *CHPT1, CEPT1, SPTLC2* and *CERS6* in SAT samples collected after eTRE (orange dots) and lTRE (petrol dots) intervention is shown. All data were normalized to the geometric mean of the housekeeper genes beta-glucuronidase (*GUSB*) and P0 of 60S ribosomal protein large subunit *RPLP0* after quantification by qPCR. N=25; data are shown as dot plots with median and IQR. Significances are encoded as follows: *p < 0.05, **p < 0.01, ***p < 0.001, by the Wilcoxon signed-rank test.

### 2.4. Effects of eTRE and lTRE on the Elongation Indices and Lipid Carbon Chain Length

Based on the levels of specific fatty acids (FA) within complex lipids (**Table S7**), we further quantified indices of desaturation and elongation as surrogate markers of corresponding enzyme activity **(Table S8)**. Between interventions, no differences of elongation indices ELOVL5 and ELOVL6 were observed **(Table S8)**, and two lipid subgroups across two lipid classes showed alterations in total carbon chain length, with each subgroup containing all lipid species from the same lipid class with identical carbon chain length ( **Table S10**). Within the eTRE, the elongation index ELOVL6 (calculated as FA 18:0 to FA 16:0 ratio^[34]^) was decreased (*P* < 0.001), whereas the ELOVL5 index (calculated as FA 20:3 to FA 18:3 ratio^[34]^) was not altered **(Figure 2B, Table S9)**. In agreement with this, the level of stearic acid 18:0 decreased within the eTRE intervention (*P* = 0.010) **(Table S7)**. The total carbon chain length was altered in 22 lipid class subgroups across 10 lipid classes **(Table S10)**. Within lTRE, FA-derived elongation indices were not altered **(Table S8)**, whereas total carbon chain length was changed in three lipid subgroups across two lipid classes (**Table S10**).

### 2.5. Effects of eTRE and lTRE on the Desaturation Indices and Lipid Double Bond Number

Between interventions, no difference of desaturation indices D5D/FADS1, D6D/FADS2, and D9D/SCD1 were observed **(Table S8)**, and two lipid subgroups changed the number of double bonds **(Table S11).** Within eTRE, the desaturation indices delta-5 desaturase (D5D/FADS1, calculated as FA 20:4 to FA 20:3 ratio ^[34–36]^, P = 0.002) and delta-9 desaturase (D9D/SCD1) for C18 (calculated as FA 18:1 to FA 18:0 ratio^[34, 37, 38]^, *P* = 0.007) increased, while the indices of delta-6 desaturase (D6D/FADS2, calculated as FA 18:3 to FA 18:2 ratio^[34]^, *P* = 0.035) decreased **(Figure 2B, Table S9)**. In agreement with this, the level of alpha-linolenic acid 18:3, used for the calculation of the D6D index), decreased following the eTRE intervention (*P* = 0.026) **(Table S7)**. In general, the number of double bonds showed alterations in 39 lipid subgroups across 12 lipid classes, with each subgroup containing all lipid species from the same lipid class with identical number of double bonds **(Table S11)**. Within lTRE, only the D5D/FADS1 index was increased (*P* < 0.001) **(Figure 2B, Table S9)**, and the number of double bonds showed alterations in six lipid subgroups across five lipid classes **(Table S11).** No FA levels were changed within lTRE (**Table S7**).

### 2.6. Effects of eTRE and lTRE on Key Lipid Enzymes in Adipose Tissue

We further hypothesized that the expression of key lipid metabolism genes in adipose tissue can be affected by eTRE and lTRE and is consistent with the alterations of ceramides and phosphatidylcholines observed in plasma. To investigate this, we assessed the expression of key genes involved in phosphatidylcholine biosynthesis (*CHPT1, CETP1*) and ceramide biosynthesis (*SPTLC2, CERS6*) in SAT samples collected after the eTRE and lTRE using qPCR **(Table S13)**. No differences of the expression of these genes were found between both interventions **(Figure 2C)**.

In addition to these genes, we have investigated the expression of genes responsible for key lipid metabolism processes in SAT, i.e. 1) desaturases (*FADS1* also known as *D5D*, *FADS2* also known as *D6D,* and *SCD1* also known as *D9D*), 2) elongases (*ELOVL5* and *ELOVL6*), and 3) genes of fatty acid synthesis (*FASN, ACC1,* and *ACSL1*). Most of the selected genes (as well as ceramide and phosphatidylcholine biosynthesis genes mentioned above) demonstrated circadian rhythms in previous human research ^[39–44]^. We therefore hypothesized that the 24-hour rhythms of these lipid species can be influenced by the timing of the eating window during eTRE and lTRE interventions. The mRNA expression after the lTRE intervention was higher than after eTRE for the desaturases *FADS1* (*P* = 0.008) and *SCD1* (*P* = 0.008) as well as for the elongase *ELOVL6* (*P* = 0.013) **(Figure 2D)**. For all other genes examined, no differences between eTRE and lTRE expression levels were found.

### 2.7. Lipid Pathway Enrichment combined with Adipose Tissue Analysis

To elucidate further lipid metabolism pathways which underwent remodeling by the TRE interventions, we conducted a lipid pathway enrichment analysis of the altered plasma lipidome as described in **Experimental section/Methods**. This analysis identified that the glycerophospholipid pathway was significantly affected by eTRE **(Table S14)**. Based on the SAT RNAseq dataset (GSE287198) collected in the same trial after both interventions, we have further analyzed the changes in expression profile of adipose tissue genes involved in the glycerophospholipid pathway **(Figure 3A).** Using the metaKEGG tool [https://metakegg.apps.dzd-ev.org, https://github.com/dife-bioinformatics/metaKEGG], we visualized observed pathway changes on both lipidomic and the transcriptomic layers. Genes found in the RNAseq dataset were directly mapped on the pathway and colored according to their log_2_(FC) **(Figure S1)**. This allowed to identify three genes, involved in the glycerophospholipid pathway and coding phospholipase enzymes, which were regulated differently after eTRE and lTRE interventions **(Figure 3B-C)**. *PLB1* and *PLA2G6* were downregulated, while *PLAG4B* was upregulated after the eTRE intervention compared to the lTRE.

**Figure 3.**
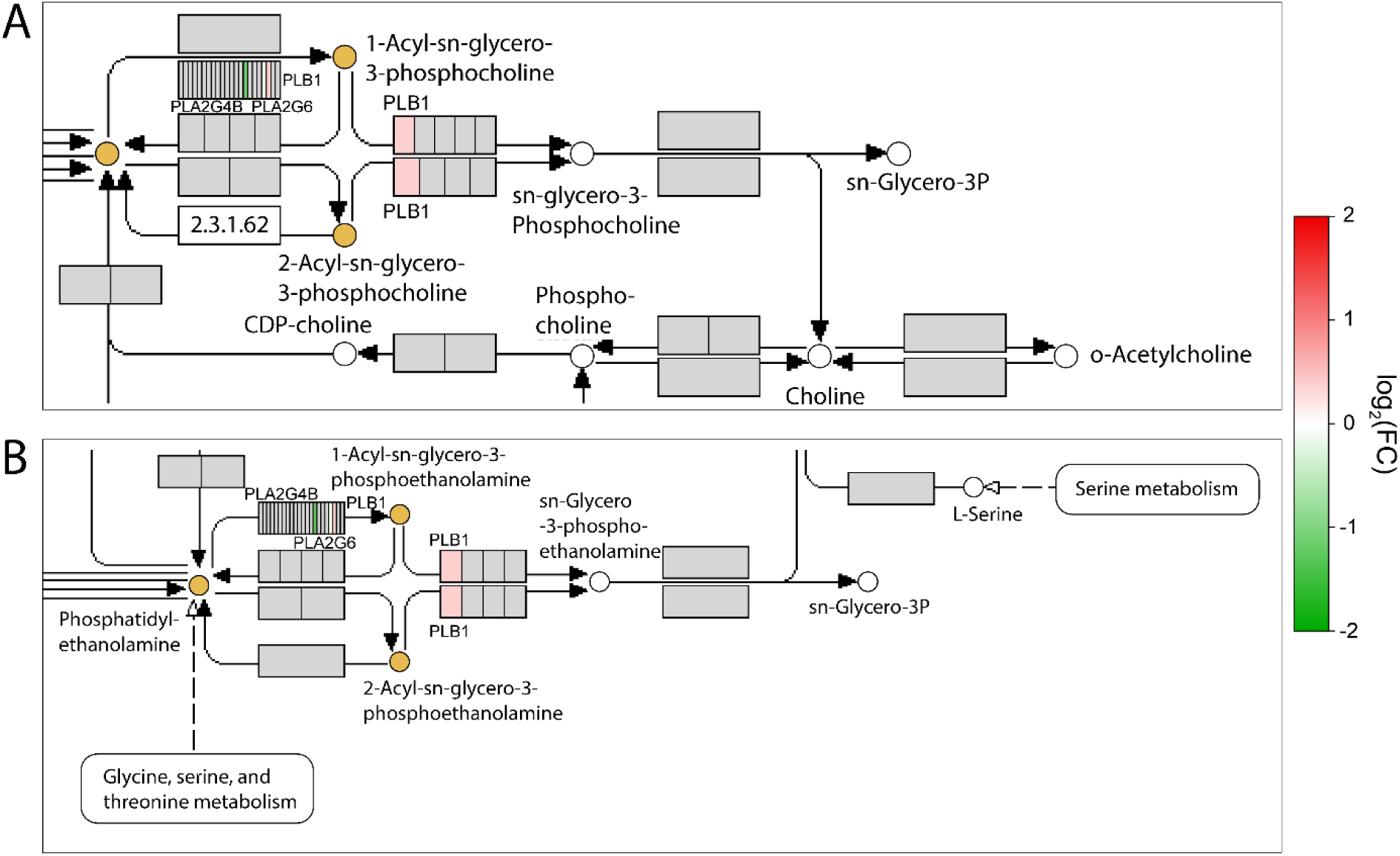
Selected areas of the glycerophospholipid pathway combining lipid and gene expression data. A) Upper zoomed area of the glycerophospholipid pathway. B) Lower zoomed area of the glycerophospholipid pathway. In (A) and (B): Glycerophospholipid pathway (KEGG ID: *hsa00564*) was identified by the lipid pathway enrichment analysis of plasma lipidome, and its full map is presented in **Figure S1**. The output of the lipid pathway enrichment analysis was subjected to the metaKEGG tool, and the KEGG compounds belonging to the enriched pathway were highlighted. Genes found in the SAT transcriptome dataset were directly mapped on the pathway and colored according to their log_2_(FC) translated to a color scale. In the color scale, red color means a positive log_2_(FC), i.e. the transcript was upregulated in ITRF compared to the eTRE, whereas green color means a negative log_2_(FC). This allowed to identify three genes coding phospholipase enzymes PLB1, PLA2G6, and PLAG4B, which were regulated differently after eTRE and lTRE interventions.

## 3. Discussion

In this work, we assessed and compared, for the first time, effects of eTRE and lTRE regimens on the human plasma lipidome using high throughput lipidomic analysis. Importantly, the TRE interventions were intended to be isocaloric, and study adherence ^[33]^ was carefully controlled to ensure that plasma lipid changes were caused by the shortening and/or timing of the eating window itself, independent of calorie deficit, changes in food composition, or physical activity. We achieved high timely adherence to both interventions, unchanged dietary composition and physical activity, although minimal caloric reduction could not be completely avoided within eTRE.

The main finding of this study is that no differences in effects on plasma lipidome were found between eTRE and lTRE, considering lipid species and classes, enzyme activity indices, carbon chain length, and the number of double bonds. This aligns with previously published studies that have found no difference in the impact of eTRE and lTRE on clinical lipid metabolism parameters, such as cholesterol and triglycerides, when directly comparing both interventions.^[14, 27, 28]^ To date, only one study by Hutchison et al. has compared eTRE and lTRE in a crossover design ^[14]^, finding that fasting triglycerides and postprandial glucose decreased similarly with both diets. Another parallel-arm trial showed comparable beneficial effects of early, late and self-selected TRE combined with a Mediterranean diet compared to a Mediterranean diet alone on blood lipids as well as visceral adipose tissue volume in individuals with overweight or obesity.^[28]^ A recent meta-analysis also confirmed comparable results of eTRE vs. lTRE on clinical lipid metabolism parameters.^[45]^ Further, our lipidomics data are consistent with data on blood lipids, glycemic control, and other cardiometabolic parameters from the same ChronoFast trial, which demonstrated no clinically relevant improvents between and within interventions in a nearly isocaloric setting.^[33]^

Nevertheless, despite the lack of between-group comparisons, we cautiously interpreted and discussed the within-intervention data, considering them as exploratory and hypothesis-generating. In within-intervention comparisons, eTRE had more pronounced effects on the plasma lipidome compared to lTRE. Indeed, eTRE caused a decrease in 103 of the analyzed lipid species compared to the levels before intervention, whereas lTRE showed no effect on lipid species. Similar differences were also observed for lipid classes and enzyme activity indices. Specifically, eTRE caused a reduction of ceramides and phosphatidylcholines, as well as alterations of several elongation and desaturation indices, while lTRE caused a change in the D5D index only. Accordingly, eTRE induced more changes in the number of total double bonds and total carbon chain length per class than lTRE.

The marked within-intervention impact of eTRE on the plasma lipidome aligns with published data showing superior metabolic effects of eTRE compared to lTRE.^[11, 19, 20, 24]^ Numerous eTRE studies have shown that restricting food intake to the beginning of the day enhances insulin sensitivity, beta-cell responsiveness, and reduces blood pressure, inflammation, and oxidative stress.^[11, 19, 20, 24]^ Conversely, lTRE, where food intake is restricted to the late afternoon or evening (after 4 pm), has either shown no effect or worsened blood glucose, beta-cell responsiveness, and lipid levels.^[5, 7, 8, 13, 46]^ The superior metabolic effects of eTRE compared to the lTRE may be explained by circadian rhythms of key metabolic processes, including lipid metabolism ^[25, 47]^.

Indeed, the molecular clock machinery induces circadian rhythms of essential lipid metabolism enzymes via clock-controlled transcription factors such as PPARs, PGC-1alpha, and SREBPs.^[48–51]^ Correspondingly, a large part of the plasma lipidome demonstrates circadian rhythmicity, as demonstrated in constant routine studies, confirming the essential role of the endogenous circadian clock in lipid homeostasis.^[49, 52, 53]^ Notably, circadian lipidome organization is altered both in circulation and in the adipose tissue in subjects with type 2 diabetes, suggesting a strong link between lipid rhythm dysregulation and diabetes pathogenesis.^[54]^ Further, we previously found that the plasma lipidome responds differently to the same meal depending on the time of day, with approximatey one-third of all lipids showing greater postprandial changes after a morning meal compared to an afternoon meal.^[31]^ This observation aligns with our finding that eTRE has more pronounced within-intervention effects on the plasma lipidome compared to lTRE.

Results of our study are also, in some degree, comparable with data of Madkur et al.^[30]^, which is the only published study to date investigating the impact of any form of intermittent fasting - in their case, Ramadan fasting - on the plasma lipidome using lipidomic analysis. Although Ramadan fasting and the TRE interventions studied in our trial differ in their eating timing, the two studies remain quite comparable as different forms of intermittent fasting. The study reported similar lipid pattern changes, particularly for sphingomyelins, after 29-30 days of the Ramadan fasting, but in contrast to our data, the ceramide class did not change.

Our finding on ceramide reduction by eTRE may be clinically relevant, as elevated ceramide levels are associated with metabolic disfunction. Numerous studies have described increased ceramide levels in context of type 2 diabetes and cardiovascular diseases^[55–59]^, as well as obesity and weight gain.^[60, 61]^ Ceramides are even being explored as biomarkers for cardiovascular diseases in daily clinical practice ^[55, 62–65]^, and some clinics have begun using ceramide-based scores to identify patients at risk for cardiometabolic diseases.^[66]^ Given these findings, the observed reduction in both the total ceramide class and the single ceramide species suggests that eTRE potentially induces greater cardiometabolic benefits than lTRE. Our data obtained in the lipid enrichment analysis of the plasma lipidome support this notion. In this analysis, we revealed a number of lipid species belonging to the glycerophospholipid pathway, which has been linked to cardiovascular risk and coronary artery disease progression.^[67, 68]^

For estimated lipid metabolism enzyme activities, similar tendencies regarding the effects of eTRE and lTRE were observed. Specifically, in the context of diabetes, a reduction in D5D activity and an elevation in D6D activity are often described as associated factors.^[69–73]^ Beyond diabetes, these changes in desaturase activities are associated with metabolic syndrome, elevated fasting plasma triglycerides, C-reactive protein, HOMA-IR, and HbA1c.^[72, 74–77]^ In this context, the observed increase in the FA-derived D5D index in both eTRE and lTRE, along with the reduction in the D6D index in lTRE, align with published beneficial metabolic effects of TRE. However, the D9D index, which was increased within eTRE, was described to be positively associated with BMI and triglyceride levels.^[71]^ Therefore, effects of TRE timing on the lipid metabolism enzyme activities warrant careful interpretation, especially considering that the index calculation in our work was based on FAs within complex lipids and not on the free FAs as in previously published studies.

In addition to the altered elongase and desaturase indices, we found that the expression of key lipid metabolism enzymes in adipose tissue differ between the two interventions depending on eating timing, highlighting the possible contribution of adipose tissue to the TRE-induced lipid changes in the circulation. The results of the expression analysis of the same genes in the SAT partly disagree with the enzyme indices findings in plasma. For example, in SAT, we observed higher expression of the desaturases FADS1/D5D and SCD1/D9D after the lTRE intervention compared to the eTRE, whereas no difference was detected in the corresponding enzyme indices in plasma. In turn, higher ELOVL6 expression levels in SAT after lTRE did not align with ELOVL6 index changes in plasma.

Following observed changes in plasma ceramide and phosphatidylcholines, we also quantified the mRNA expression of key enzymes involved in their metabolism. In particular, CHPT1 and SPTLC2, essential genes of ceramide and phosphatidylcholine metabolism, respectively, have been previously characterized as circadian enzymes in human muscles.^[78]^ Based on the lipidomic data and considering the *zeitgeber* role of food intake for central peripheral circadian clocks as shown in animal^[79]^ and few human trials^[33, 80, 81]^, we expected to observe expression differences between eTRE and lTRE. However, contrary to our expectations, no significant differences were found in the expression of these and other analyzed genes between samples collected after eTRE and lTRE. This discrepancy, along with previously mentioned differences between circulating lipids and adipose tissue expression levels, may be attributable to the fact that circulating lipids are primarily synthesized in the liver; thus, their levels may not accurately reflect ongoing metabolism in SAT. Further, mRNA levels do not account for the translational and post-translational regulation of corresponding enzymes. Similarly, we found no differences between eTRE and lTRE in the expression of genes involved in fatty acid synthesis (FASN, ACC1 and ACSL1), which have previously shown to exhibit circadian rhythmicity ^[43, 44]^.

By combining lipid pathway enrichment analysis and transcriptome data from SAT using the novel metaKEGG tool, we identified three genes - PLB1, PLA2G6, and PLAG4B - that appear to be involved in the effects of TRE timing on lipid homeostasis. These genes contribute to the glycerophospholipid pathway and encode phospholipase enzymes, cleaving acyl chains from the *sn-*1 and *sn*-2 positions of a phospholipid. Importantly, these three genes were regulated differently by the TRE timing: PLB1 and PLA2G6 were downregulated, while PLAG4B was upregulated after the eTRE intervention compared to the lTRE. Whereas PLA2G6 is known to be a proinflammatory mediator required for monocyte chemotaxis and potentially contributory to both atherosclerosis and diabetes^[82]^, the link between both PLAG4B & PLB1 and obesity, diabetes, or cardiovascular diseases is currently unknown. Nevertheless, our findings provide new insight into the role of phospholipases in the effects of TRE.

Several strengths and limitations should be considered when interpreting the results of this work. As mentioned above, the primary strength is an absolute novelty of the data presented regarding the TRE effects on the plasma lipidome. Secondly, this study directly compared the effects of predefined early vs. late eating windows on the plasma lipidome within a crossover design, that minimizes the impact of interindividual variability. The third strength was the maintanence of unchanged food composition, achieved through intensive nutritional counselling and careful control of food intake, as detailed previously.^[32]^ Finally, the combined analysis of the plasma lipidome and adipose tissue transcriptome represents an important strength, providing insight into the role of adipose tissue in lipid homeostasis regulation by the TRE intervention.

The important limitation of the data interpretation is an analysis of the within-intervention changes despite the lack of between-intervention difference. To notice, this doesn’t automatically mean that neither intervention work. The reason can be the relatively small effect size or insufficient power, because our study was powered for assessing the primary outcome insulin sensitivity. Remarkably, we consider our data as exploratory and hypothesis-generating, and therefore future, adequately powered lipidomic studies are needed to confirm these findings.

Another practical limitation stems from the challenges in maintaining strict caloric equivalence across the interventions. Despite intensive real-time dietary supervision, in the eTRE, a minimal caloric reduction could not be entirely avoided, although it was lower than observed in comparable TRE studies.^[6, 7, 10, 27, 83]^ In both interventions, minor weight loss of ∼ 1% was observed after two weeks, consistent with previous isocaloric or nearly isocaloric TRE trials ^[14, 84–86]^. Of note, weight loss was slightly, but significantly, greater in the eTRE compared to the lTRE. Such minor weight changes would expectedly not meaningfully affect lipid profile, because metabolic changes are usually induced by >5% weight loss. Nevertheless, we cannot completely exclude the impact of the weight loss on lipidomic changes attributed to TRE timing, which adds complexity to the data interpretation. The third limitation lies in the indirect assessment of enzyme activity levels, which were estimated based on fatty acid levels within complex lipids and should therefore be interpreted with caution. The fourth limitation is that the trial was conducted in mostly postmenopausal women, potentially limiting the generalizability of our findings to the broader population. Finally, the intervention periods were short (two weeks), which was necessary to maximize adherence to prescribed eating times and isocaloric energy intake, facilitated by real-time monitoring and individual councelling, and to ensure extensive documentation of interstitial glucose, physical activity, and sleep behavior ^[33]^. The limited sample size is also acknowledged as a potential limitation; however, its impact is mitigated by the crossover study design. Nevertheless, future research in other population groups and larger cohorts needs to confirm the results of our study.

## 3. Conclusion

In summary, our study provides new insights into the effects of TRE on plasma lipidome. Between interventions, lipid species and classes, as well as enzyme activity indices, are not substantially changed. However, the exploratory analysis of within-intervention comparisons highlighted marked differences between the effects of eTRE and lTRE on the plasma lipidome and adipose tissue. These results suggest that eating timing during TRE might be essential for remodeling of the plasma lipidome and adipose tissue transcriptome, highlighting the need of future lipidomic research in TRE.

## 4. Experimental Section/Methods

### Study design & Participant Characteristics

31 women with overweight or obesity completed the randomized 10-weeks crossover clinical trial, which was conducted at the German Institute of Human Nutrition Potsdam-Rehbruecke, Germany, from April 2020 to December 2021. Details of the participant recruitment, clinical characteristics of the study participants, dietary interventions, and adherence assessment were recently published.^[32, 33]^ Inclusion and exclusion criteria are described in **Table S1**. In brief, inclusion criteria were female sex, age between 18 and 70 years, and BMI between 25 and 35 kg/m^2^. Exclusion criteria were diabetes type 1 or type 2, shift work, and travel across more than one time zone one moths before or during the study. The study protocol and informed consent form were approved by the Medical Ethics Committee of the University of Potsdam (EA No. 8/2019) and comply with the Declaration of Helsinki of 1975 as revised in 2013. All participants provided written informed consent prior to study participation. The study was registered at clinicaltrials.gov on 17.04.2020 (Identifier: NCT04351672). Study results were reported using the CONSORT 2025 checklist guideline for reporting randomized trials.^[87]^ All resources and reagents related to this trial are listed in the supporting information (**Table S15**).

For a detailed description of the study protocol, refer to.^[32]^ In brief, after a two-to-four-week baseline phase, two intended isocaloric two-week interventions – (i) eTRE and (ii) lTRE - were conducted, separated by a two-week washout phase **(Figure 1A)**. During the baseline phase, the participants consumed their usual food quality and quantity at their usual meal times. In the eTRE intervention, participants were asked to consume the same kind and amount of food as during the baseline phase but to reduce their eating window to 8 h per day for a daytime, from 8 am to 4 pm. In the lTRE intervention, the 8-hour eating window shifted to 1 pm to 9 pm. Between the two interventions, participants returned to their usual eating times. Throughout the 8-10 weeks of the study, participants were asked to maintain their usual lifestyle habits in terms of physical activity and sleep. Eating behavior (timing, kind, and amount of food) was monitored using digital or paper-based food records, while physical activity and sleep-wake rhythms were controlled by the actigraphy (ActiGraph wGT3X-BT, ActiGraph, Netherlands) and sleep logs, respectively, for 14 consecutive days of the baseline phase and during both TRE interventions. Energy intake, macronutrient composition, and eating times were assessed using the FDDB food database (Fddb Internetportale GmbH, https://fddb.info/) as described.^[32]^ All days at which the subjects followed the required TRE eating time frame with a deviation of maximally ± 30 minutes were considered as compliant. As a measure of physical activity, the metabolic equivalent of task (MET) was analyzed using an ActiLife software version 6.13.4 (ActiGraph, Netherlands).

At intitial screening and before and after each intervention anthropometric measurements were conducted. Body composition was examined in a fasting state with a bioimpedance analyzer (BIA; Quantum S, Akern, Florence, Italy) using the BIA-related Bodygram^TM^ software (Akern, Italy). Height measurements were assessed using a stadiometer, and weight measures were assessed using a digital scale, for the calculation of BMI, defined as weight in kilograms divided by the square of the height in metres. Circumferences of waist and hip were measured with a measuring tape.

Beyond that, metabolic examination of participants was conducted after an overnight fast, using 75 g-oral glucose tolerance test (OGTT) and blood sample collection as described.^[32]^ Screening OGTT was used to characterize participants’ metabolic state as follows: individuals were classified as having impaired fasting glucose (IFG) if their fasting glucose was 100-125 mg/dL, and as having impaired glucose tolerance (IGT) if their 2-hour glucose was 140-199 mg/dL. Blood samples were collected in serum monovettes for blood glucose, HbA1c, and total lipid measurements. Participants’ chronotypes were determined using the Munich Chronotype Questionnaire (MCTQ) and the Horne-Östberg Morningness-Eveningness Questionnaire (MEQ) as described.^[32, 33]^ Chronotype classification in the analysis was performed on MSFsc value (midsleep on free days corrected for the sleep debt over the working days) from MCTQ using the following cut-off-points: MCFsc <4 was defined as an early chronotype, MCF-Sc >5 as a late chronotype, and intermediate values as a normal chronotype. Classification in the MEQ was performed as follows: a range 59-69 defined early chronotypes, a range 31-41 defined late chronotypes, and intermediate values for normal chronotypes. In case of heterogenous results in both questionnaires, the MCTQ was used for the final chronotype classification.

### Blood Plasma and Adipose Tissue Collection

For lipidomics analysis, fasting blood samples were drawn at 9.30 a.m. before and after eTRE and lTRE interventions **(visits 1-4 at Figure 1A)** using EDTA monovettes (Sarstedt, Germany). Blood plasma was frozen immediately after the centrifugation and stored at −80°C until analysis. Subcutaneous adipose tissue (SAT) was collected periumbilically at 9.30 a.m. after both interventions (visits 2 and 4). Before each biopsy, a skin area of approximately 2×2 cm was treated with lidocaine for anesthesia. A 3-mm incision was then made in the skin using a fine needle (diameter 2.1 mm) connected to a vacuum syringe containing sterile NaCl solution. The resulting negative pressure allowed small pieces of tissue to be removed. These were immediately cleaned in a NaCl solution, frozen in aliquots a 0.4 g in liquid nitrogen and stored at −80 °C until analysis.

### Lipidomics Analyses of Plasma Samples

Lipidomic measurements in blood plasma were performed using mass spectrometry-based shotgun lipidomic analysis at Lipotype GmbH (Dresden, Germany) as described ^[29]^. For lipid extraction, an equivalent of 1 μl of undiluted plasma was used, and plasma lipids were extracted with methyl tert-butyl ether/methanol (7:2, V:V).^[88]^ Samples were analyzed by direct infusion in a QExactive mass spectrometer (Thermo Scientific, Bremen, Germany) equipped with a TriVersa NanoMate ion source (Advion Biosciences, Ithaca, New York, United States of America). Samples were analyzed in both positive and negative ion modes with a resolution of R_m/z= 200_ = 280,000 for MS and _Rm/z= 200_ = 17,500 for MSMS experiments in a single acquisition. Lipids were identified and quantified employing the proprietary LipotypeXplorer software. Lipid intensities were normalized to lipid class-specific internal standards, and data were reported as molar amounts. Analytical quality was evaluated by incorporating reference and blank samples. Data were corrected for batch effects and drift based on reference samples. Lipid species present in less than 70% of all samples were excluded. The Lipidomics Standard Initiative minimal reporting checklist^[89]^ for this study can be found at https://doi.org/10.5281/zenodo.15183498.

Lipid species were annotated based on their molecular composition, indicating the sum of carbon atoms in the hydrocarbon moiety, the sum of double bonds, and the sum of hydroxyl groups. For instance, PI 34:1;0 denotes phosphatidylinositol with a total length of its fatty acids equal to 34 carbon atoms, total number of double bonds in its fatty acids equal to 1, and 0 hydroxylations. The annotation of lipid subspecies provides additional information on the exact identity of their acyl moieties and their sn-position (if available). For example, PI 18:1;0_16:0;0 denotes phosphatidylinositol with octadecenoic (18:1;0) and hexadecanoic (16:0;0) fatty acids, for which the exact position (sn-1 or sn-2) in relation to the glycerol backbone cannot be discriminated (underscore "_" separating the acyl chains). In contrast, PC O-18:1;0/16:0;0 denotes an ether-phosphatidylcholine, where an alkyl chain with 18 carbon atoms and 1 double bond (O-18:1;0) is ether-bound to sn-1 position of the glycerol, and a hexadecanoic acid (16:0;0) is connected via an ester bond to the sn-2 position of the glycerol (slash "/" separating the chains signifies that the sn-position on the glycerol can be resolved). Lipid identifiers from the SwissLipids database^[90]^ and the shorthand notation for MS-derived lipid structures^[91]^ are provided. All lipid species and lipids classes identified by this method are shown in **Table S3**.

### Assessment of Lipid Enzyme Activity Indices

Based on the level of specific lipids, the overall amount of fatty acids (FA) within complex lipid was assessed. Enzyme activity indices were calculated as ratio between two FAs and used as surrogate activity markers of corresponding enzyme responsible for the conversion from one FA to another. Following indices were assessed in this study: delta-5 desaturase (D5D/FADS1), delta-6 desaturase (D6D/FADS2) ^[34]^, elongation of very long chain fatty acids protein 5 (ELOVL5)^[34]^, elongation of very long chain fatty acids protein 6 (ELOVL6)^[34]^, stearoyl-CoA desaturase 1/delta-9 desaturase - C16 (SCD1/D9D (C16)), stearoyl-CoA desaturase 1/delta-9 desaturase - C18 (SCD1/D9D (C18)), stearoyl-CoA desaturase 1/delta-9 desaturase - C18+16 (SCD1/D9D (C18+16))^[36–38]^ **(Table S4)**.

### Gene Expression Analyses of Adipose Tissue

Total RNA was isolated from subcutaneous adipose tissue (SAT) samples using the RNeasy Lipid Tissue Mini Kit (Qiagen, Germany). RNA concentration was determined utilizing an ND-1000 spectrophotometer (Nanodrop, PeqLab), and the RNA quality was determined by the Bioanalyzer (Agilent Technologies, USA). Extracted RNA samples all had RIN values of ≥ 8. For the qPCR analysis, single-stranded cDNA was generated using the QuantiTect Reverse Transcription Kit (Qiagen, Germany). Quantitative PCR (qPCR) was conducted on the ViiA 7 sequence detection system using Power SYBR Green PCR Master Mix (Applied Biosystems, USA) and specific primers **(Table S5)**. Gene expression was evaluated using the standard curve method and normalized to the geometric mean of housekeeper genes beta-glucuronidase (GUSB) and P0 protein of 60S ribosomal protein large subunit (RPLPO). Whole-genome transcriptome analysis was performed by RNA sequencing by BGI Genomics (BGI-Copenhagen, Denmark) using BGISEQ platform with 100 bp paired-end reads. RNAseq data was submitted to the GEO database with an assession number GSE287198.

### RNAseq Data Processing

RNAseq data were delivered by BGI in form of adapter and low-quality reads cleaned sequencing data. FastQC v0.12.1 was employed to assess quality of the samples. Reads were aligned to the reference genome (GRCh38.110/hg38) using STAR 2.7.11a, and fragments per kilobase per million (FPKM) values for transcripts were determined by STRINGTIE v2.2.1, both with default options for paired reads. Differential gene expression analysis was performed via the R-package DESeq2 (v1.34.0) pipeline. Transcripts with mean expression values > 1 FPKM, and p < 0.05 were considered statistically significant. RNAseq data processing was performed with R v4.1.2.

### Lipid Pathway Enrichment combined with SAT Transcriptome Analysis

All the differentially abundant lipid species were subjected to lipid pathway enrichment analysis using online tool LIPEA (https://hyperlipea.org/). The lipid species belonging to the same lipid class were merged and KEGG pathway enrichment analysis^[92–96]^ output included the lipid/compound ID mapping to the enriched pathways.

The output of the lipid pathway enrichment analysis was subjected to metaKEGG tool [https://metakegg.apps.dzd-ev.org, https://github.com/dife-bioinformatics/metaKEGG] to visualize lipidomic and transcriptomic layers. For this application, we used the analysis pipeline “Bulk RNAseq mapping” in order to directly map the KEGG pathway that emerged from the lipid pathway enrichment analysis, bypassing the need for a gene pathway enrichment analysis. The pathway “Glycerophospholipid metabolism” (KEGG ID: *hsa00564*) was queried, using log_2_(FC) values from only significantly (1 FPKM & p < 0.05) differentially expressed genes of the comparison of eTRF vs. lTRF. The internal KEGG compound IDs for lipid species identified in enriched pathways were provided as an additional input parameter for the metaKEGG visualization pipeline. Genes found in the SAT RNAseq dataset were directly mapped on the pathway, colored according to their log_2_(FC) translated to a color scale, while KEGG compounds were assigned a single color to highlighted their presence in the pathway. *Statistical Analysis:* Statistical power of the trial was calculated for the main study outcome, insulin sensitivity, as described ^[32]^. For this secondary analysis we refer to previously published nutritional lipidomic studies with a similar or even lower sample size and crossover design.^[97, 98]^ Statistical analyses were performed with SPSS v.25 (SPSS, Chicago, IL) and R version 4.2.3 (2023-03-15) using tidyverse packages (version 2.0.0). P < 0.05 was considered statistically significant in all analyses. Comparison of lipid data between two groups was conducted using a paired Mann-Whitney U Test (Wilcoxon signed-rank test). For the multiple testing correction, the Benjamini–Hochberg (BH) method was used. Data are presented as means (standard deviation, SD) for parameters with normal distribution and medians (25th and 75th interquartile range, IQR) for parameters with non-normal distribution if not stated otherwise. Changes within eTRE or lTRE intervention (calculated as a delta of the value after the intervention minus the value before the intervention) and between-intervention differences (calculated as a delta of the change within lTRE minus the change within eTRE) were expressed as mean (95% CI).

## Supporting Information

Supporting Information is available from the Wiley Online Library or from the author.

## Acknowledgements

We acknowledge all study participants for their cooperation. We gratefully thank Juliane Roeder, Melanie Hannemann, and Lothar Napieralski for their work with study subjects; Katja Treu and Christiana Gerbracht for the help in the preparation of nutritional counselling; Marion Urbich and Nadine Huckauf for excellent technical assistance in the sample analysis. We also thank all students who contributed to the preparation and conduction of the clinical trial. The study was supported by the German Research Foundation (DFG RA 3340/3-1, project number 434112826 and DFG RA 3340/4-1, project number 530918029 to OP-R), by the German Diabetes Association (Allgemeine Projektförderung der DDG 2020 and 2023 to OP-R); and by the European Association for Study of Diabetes 2020 (Morgagni Prize 2020 to OP-R). The DZD is funded by the German Federal Ministry for Education and Research (01GI0925).

## Author Contributions

M.J.G. performed lipidomics experiments; M.J.G. and S.K. performed the data analyses; A.M., A.F.H.P., A.K., and O. P-R conceptualized the clinical trial; B.P., J.S., and B.S. conducted the trial and provided the samples; C.K. and K.Si contributed to the lipidomics conceptualization; M.L. and R.S. conducted RNAseq data processing and lipid pathway enrichment analysis; K.S. drafted the manuscript; O.P-R. edited the manuscript. All authors readand approved the final manuscript.

## Data Availability Statement

The lipidomic and clinical data that support the findings of this study are available from the corresponding author upon reasonable request. RNAseq data is publicly available in the GEO database (GSE287198, uploaded, will be reaseased after the manuscript acception) as listed in the key resources table **(Table S15)**.

## References

1. Barnosky, A. R., K. K. Hoddy, T. G. Unterman and K. A. Varady. "Intermittent fasting vs daily calorie restriction for type 2 diabetes prevention: A review of human findings." Transl Res 164 (2014): 302–11. 10.1016/j.trsl.2014.05.013.

2. Gill, S. and S. Panda. "A smartphone app reveals erratic diurnal eating patterns in humans that can be modulated for health benefits." Cell Metab 22 (2015): 789–98. 10.1016/j.cmet.2015.09.005. https://www.ncbi.nlm.nih.gov/pubmed/26411343.

3. Chow, L. S., E. N. C. Manoogian, A. Alvear, J. G. Fleischer, H. Thor, K. Dietsche, Q. Wang, J. S. Hodges, N. Esch, S. Malaeb, et al. "Time-restricted eating effects on body composition and metabolic measures in humans who are overweight: A feasibility study." Obesity (Silver Spring) 28 (2020): 860–69. 10.1002/oby.22756. https://www.ncbi.nlm.nih.gov/pubmed/32270927.

4. Cai, H., Y. L. Qin, Z. Y. Shi, J. H. Chen, M. J. Zeng, W. Zhou, R. Q. Chen and Z. Y. Chen. "Effects of alternate-day fasting on body weight and dyslipidaemia in patients with non-alcoholic fatty liver disease: A randomised controlled trial." BMC Gastroenterol 19 (2019): 219. 10.1186/s12876-019-1132-8. https://www.ncbi.nlm.nih.gov/pubmed/31852444.

5. Lowe, D. A., N. Wu, L. Rohdin-Bibby, A. H. Moore, N. Kelly, Y. E. Liu, E. Philip, E. Vittinghoff, S. B. Heymsfield, J. E. Olgin, et al. "Effects of time-restricted eating on weight loss and other metabolic parameters in women and men with overweight and obesity: The treat randomized clinical trial." JAMA Intern Med 180 (2020): 1491–99. 10.1001/jamainternmed.2020.4153. https://www.ncbi.nlm.nih.gov/pubmed/32986097.

6. Gabel, K., K. K. Hoddy, N. Haggerty, J. Song, C. M. Kroeger, J. F. Trepanowski, S. Panda and K. A. Varady. "Effects of 8-hour time restricted feeding on body weight and metabolic disease risk factors in obese adults: A pilot study." Nutr Healthy Aging 4 (2018): 345–53. 10.3233/NHA-170036. https://www.ncbi.nlm.nih.gov/pubmed/29951594.

7. Cienfuegos, S., K. Gabel, F. Kalam, M. Ezpeleta, E. Wiseman, V. Pavlou, S. Lin, M. L. Oliveira and K. A. Varady. "Effects of 4- and 6-h time-restricted feeding on weight and cardiometabolic health: A randomized controlled trial in adults with obesity." Cell Metab 32 (2020): 366–78.e3. 10.1016/j.cmet.2020.06.018.

8. Moro, T., G. Tinsley, A. Bianco, G. Marcolin, Q. F. Pacelli, G. Battaglia, A. Palma, P. Gentil, M. Neri and A. Paoli. "Effects of eight weeks of time-restricted feeding (16/8) on basal metabolism, maximal strength, body composition, inflammation, and cardiovascular risk factors in resistance-trained males." J Transl Med 14 (2016): 290. 10.1186/s12967-016-1044-0. https://www.ncbi.nlm.nih.gov/pubmed/27737674.

9. Kesztyus, D., P. Cermak, M. Gulich and T. Kesztyus. "Adherence to time-restricted feeding and impact on abdominal obesity in primary care patients: Results of a pilot study in a pre-post design." Nutrients 11 (2019): 10.3390/nu11122854. https://www.ncbi.nlm.nih.gov/pubmed/31766465.

10. Wilkinson, M. J., E. N. C. Manoogian, A. Zadourian, H. Lo, S. Fakhouri, A. Shoghi, X. Wang, J. G. Fleischer, S. Navlakha, S. Panda, et al. "Ten-hour time-restricted eating reduces weight, blood pressure, and atherogenic lipids in patients with metabolic syndrome." Cell Metab 31 (2020): 92–104.e5. 10.1016/j.cmet.2019.11.004.

11. Jones, R., P. Pabla, J. Mallinson, A. Nixon, T. Taylor, A. Bennett and K. Tsintzas. "Two weeks of early time-restricted feeding (etrf) improves skeletal muscle insulin and anabolic sensitivity in healthy men." Am J Clin Nutr 112 (2020): 1015–28. 10.1093/ajcn/nqaa192. https://www.ncbi.nlm.nih.gov/pubmed/32729615.

12. McAllister, M. J., B. L. Pigg, L. I. Renteria and H. S. Waldman. "Time-restricted feeding improves markers of cardiometabolic health in physically active college-age men: A 4-week randomized pre-post pilot study." Nutr Res 75 (2020): 32–43. 10.1016/j.nutres.2019.12.001. https://www.ncbi.nlm.nih.gov/pubmed/31955013.

13. Tinsley, G. M., J. S. Forsse, N. K. Butler, A. Paoli, A. A. Bane, P. M. La Bounty, G. B. Morgan and P. W. Grandjean. "Time-restricted feeding in young men performing resistance training: A randomized controlled trial." Eur J Sport Sci 17 (2017): 200–07. 10.1080/17461391.2016.1223173. https://www.ncbi.nlm.nih.gov/pubmed/27550719.

14. Hutchison, A. T., P. Regmi, E. N. C. Manoogian, J. G. Fleischer, G. A. Wittert, S. Panda and L. K. Heilbronn. "Time-restricted feeding improves glucose tolerance in men at risk for type 2 diabetes: A randomized crossover trial." Obesity (Silver Spring) 27 (2019): 724–32. 10.1002/oby.22449. https://www.ncbi.nlm.nih.gov/pubmed/31002478.

15. Karras, S. N., T. Koufakis, L. Adamidou, V. Antonopoulou, P. Karalazou, K. Thisiadou, E. Mitrofanova, H. Mulrooney, A. Petroczi, P. Zebekakis, et al. "Effects of orthodox religious fasting versus combined energy and time restricted eating on body weight, lipid concentrations and glycaemic profile." Int J Food Sci Nutr 72 (2021): 82–92. 10.1080/09637486.2020.1760218. https://www.ncbi.nlm.nih.gov/pubmed/32362210.

16. Li, C., C. Xing, J. Zhang, H. Zhao, W. Shi and B. He. "Eight-hour time-restricted feeding improves endocrine and metabolic profiles in women with anovulatory polycystic ovary syndrome." J Transl Med 19 (2021): 148. 10.1186/s12967-021-02817-2. https://www.ncbi.nlm.nih.gov/pubmed/33849562.

17. Peeke, P. M., F. L. Greenway, S. K. Billes, D. Zhang and K. Fujioka. "Effect of time restricted eating on body weight and fasting glucose in participants with obesity: Results of a randomized, controlled, virtual clinical trial." Nutr Diabetes 11 (2021): 6. 10.1038/s41387-021-00149-0. https://www.ncbi.nlm.nih.gov/pubmed/33446635.

18. Phillips, N. E., J. Mareschal, N. Schwab, E. N. C. Manoogian, S. Borloz, G. Ostinelli, Gauthier-Jaques, S. Umwali, E. Gonzalez Rodriguez, D. Aeberli, et al. "The effects of time-restricted eating versus standard dietary advice on weight, metabolic health and the consumption of processed food: A pragmatic randomised controlled trial in community-based adults." Nutrients 13 (2021): 10.3390/nu13031042. https://www.ncbi.nlm.nih.gov/pubmed/33807102.

19. Sutton, E. F., R. Beyl, K. S. Early, W. T. Cefalu, E. Ravussin and C. M. Peterson. "Early time-restricted feeding improves insulin sensitivity, blood pressure, and oxidative stress even without weight loss in men with prediabetes." Cell Metab 27 (2018): 1212–21.e3. 10.1016/j.cmet.2018.04.010.

20. Jamshed, H., R. A. Beyl, D. L. Della Manna, E. S. Yang, E. Ravussin and C. M. Peterson. "Early time-restricted feeding improves 24-hour glucose levels and affects markers of the circadian clock, aging, and autophagy in humans." Nutrients 11 (2019): 10.3390/nu11061234. https://www.ncbi.nlm.nih.gov/pubmed/31151228.

21. Goldberg, I. J. "Diabetic dyslipidemia: Causes and consequences." The Journal of Clinical Endocrinology & Metabolism 86 (2001): 965–71. 10.1210/jcem.86.3.7304. https://doi.org/10.1210/jcem.86.3.7304.

22. Zeb, F., X. Wu, L. Chen, S. Fatima, I. U. Haq, A. Chen, F. Majeed, Q. Feng and M. Li. "Effect of time-restricted feeding on metabolic risk and circadian rhythm associated with gut microbiome in healthy males." Br J Nutr 123 (2020): 1216–26. 10.1017/S0007114519003428. https://www.ncbi.nlm.nih.gov/pubmed/31902372.

23. Parr, E. B., B. L. Devlin, K. H. C. Lim, L. N. Z. Moresi, C. Geils, L. Brennan and J. A. Hawley. "Time-restricted eating as a nutrition strategy for individuals with type 2 diabetes: A feasibility study." Nutrients 12 (2020): 10.3390/nu12113228. https://www.ncbi.nlm.nih.gov/pubmed/33105701.

24. Ravussin, E., R. A. Beyl, E. Poggiogalle, D. S. Hsia and C. M. Peterson. "Early time-restricted feeding reduces appetite and increases fat oxidation but does not affect energy expenditure in humans." Obesity (Silver Spring) 27 (2019): 1244–54. 10.1002/oby.22518. https://www.ncbi.nlm.nih.gov/pubmed/31339000.

25. Schuppelius, B., B. Peters, A. Ottawa and O. Pivovarova-Ramich. "Time restricted eating: A dietary strategy to prevent and treat metabolic disturbances." Front Endocrinol (Lausanne) 12 (2021): 683140. 10.3389/fendo.2021.683140. https://www.ncbi.nlm.nih.gov/pubmed/34456861.

26. Xie, Z., Y. Sun, Y. Ye, D. Hu, H. Zhang, Z. He, H. Zhao, H. Yang and Y. Mao. "Randomized controlled trial for time-restricted eating in healthy volunteers without obesity." Nat Commun 13 (2022): 1003. 10.1038/s41467-022-28662-5. https://www.ncbi.nlm.nih.gov/pubmed/35194047.

27. Zhang, L. M., Z. Liu, J. Q. Wang, R. Q. Li, J. Y. Ren, X. Gao, S. S. Lv, L. Y. Liang, F. Zhang, B. W. Yin, et al. "Randomized controlled trial for time-restricted eating in overweight and obese young adults." iScience 25 (2022): 104870. 10.1016/j.isci.2022.104870. https://www.ncbi.nlm.nih.gov/pubmed/36034217.

28. Dote-Montero, M., A. Clavero-Jimeno, E. Merchán-Ramírez, M. Oses, J. Echarte, A. Camacho-Cardenosa, M. Concepción, F. J. Amaro-Gahete, J. M. A. Alcántara, A. López-Vázquez, et al. "Effects of early, late and self-selected time-restricted eating on visceral adipose tissue and cardiometabolic health in participants with overweight or obesity: A randomized controlled trial." Nat Med 31 (2025): 524–33. 10.1038/s41591-024-03375-y.

29. Surma, M. A., R. Herzog, A. Vasilj, C. Klose, N. Christinat, D. Morin-Rivron, K. Simons, M. Masoodi and J. L. Sampaio. "An automated shotgun lipidomics platform for high throughput, comprehensive, and quantitative analysis of blood plasma intact lipids." Eur J Lipid Sci Technol 117 (2015): 1540–49. 10.1002/ejlt.201500145.

30. Madkour, M. I., M. T. Islam, T. S. Tippetts, K. H. Chowdhury, L. A. Lesniewski, S. A. Summers, F. Zeb, D. N. Abdelrahim, R. AlKurd, H. M. Khraiwesh, et al. "Ramadan intermittent fasting is associated with ameliorated inflammatory markers and improved plasma sphingolipids/ceramides in subjects with obesity: Lipidomics analysis." Sci Rep 13 (2023): 17322. 10.1038/s41598-023-43862-9. https://www.nature.com/articles/s41598-023-43862-9.pdf.

31. Kessler, K., M. J. Gerl, S. Hornemann, M. Damm, C. Klose, K. J. Petzke, M. Kemper, D. Weber, N. Rudovich, T. Grune, et al. "Shotgun lipidomics discovered diurnal regulation of lipid metabolism linked to insulin sensitivity in nondiabetic men." J Clin Endocrinol Metab 105 (2020): 10.1210/clinem/dgz176.

32. Peters, B., D. A. Koppold-Liebscher, B. Schuppelius, N. Steckhan, A. F. H. Pfeiffer, A. Kramer, A. Michalsen and O. Pivovarova-Ramich. "Effects of early vs. Late time-restricted eating on cardiometabolic health, inflammation, and sleep in overweight and obese women: A study protocol for the chronofast trial." Front Nutr 8 (2021): 765543. 10.3389/fnut.2021.765543. https://www.ncbi.nlm.nih.gov/pubmed/34869534.

33. Peters, B., J. Schwarz, B. Schuppelius, A. Ottawa, D. A. Koppold, D. Weber, N. Steckhan, K. Mai, T. Grune, A. F. H. Pfeiffer, et al. "Effects of isocaloric early vs. Late time-restricted eating on insulin sensitivity, cardiometabolic health, and internal circadian time in women with overweight or obesity." medRxiv (2024): 2024.10.05.24314120. 10.1101/2024.10.05.24314120. https://www.medrxiv.org/content/medrxiv/early/2024/10/07/2024.10.05.24314120.full.pdf.

34. Kjellqvist, S., C. Klose, M. A. Surma, G. Hindy, I. G. Mollet, A. Johansson, P. Chavaux, J. Gottfries, K. Simons, O. Melander, et al. "Identification of shared and unique serum lipid profiles in diabetes mellitus and myocardial infarction." J Am Heart Assoc 5 (2016): 10.1161/jaha.116.004503.

35. Gieger, C., L. Geistlinger, E. Altmaier, M. Hrabé de Angelis, F. Kronenberg, T. Meitinger, H. W. Mewes, H. E. Wichmann, K. M. Weinberger, J. Adamski, et al. "Genetics meets metabolomics: A genome-wide association study of metabolite profiles in human serum." PLoS Genet 4 (2008): e1000282. 10.1371/journal.pgen.1000282.

36. Warensjö, E., M. Ohrvall and B. Vessby. "Fatty acid composition and estimated desaturase activities are associated with obesity and lifestyle variables in men and women." Nutr Metab Cardiovasc Dis 16 (2006): 128–36. 10.1016/j.numecd.2005.06.001.

37. Vessby, B., I. B. Gustafsson, S. Tengblad, M. Boberg and A. Andersson. "Desaturation and elongation of fatty acids and insulin action." Ann N Y Acad Sci 967 (2002): 183–95. 10.1111/j.1749-6632.2002.tb04275.x.

38. Attie, A. D., R. M. Krauss, M. P. Gray-Keller, A. Brownlie, M. Miyazaki, J. J. Kastelein, A. J. Lusis, A. F. Stalenhoef, J. P. Stoehr, M. R. Hayden, et al. "Relationship between stearoyl-coa desaturase activity and plasma triglycerides in human and mouse hypertriglyceridemia." J Lipid Res 43 (2002): 1899–907. 10.1194/jlr.m200189-jlr200.

39. Chen, M., Y. Lin, Y. Dang, Y. Xiao, F. Zhang, G. Sun, X. Jiang, L. Zhang, J. Du, S. Duan, et al. "Reprogramming of rhythmic liver metabolism by intestinal clock." J Hepatol 79 (2023): 741–57. 10.1016/j.jhep.2023.04.040.

40. Gréchez-Cassiau, A., C. Feillet, S. Guérin and F. Delaunay. "The hepatic circadian clock regulates the choline kinase α gene through the bmal1-rev-erbα axis." Chronobiol Int 32 (2015): 774–84. 10.3109/07420528.2015.1046601.

41. Jang, Y. S., Y. J. Kang, T. J. Kim and K. Bae. "Temporal expression profiles of ceramide and ceramide-related genes in wild-type and mper1/mper2 double knockout mice." Mol Biol Rep 39 (2012): 4215–21. 10.1007/s11033-011-1207-2.

42. Zhou, X., D. Wan, Y. Zhang, Y. Zhang, C. Long, S. Chen, L. He, B. Tan, X. Wu and Y. Yin. "Diurnal variations in polyunsaturated fatty acid contents and expression of genes involved in their de novo synthesis in pigs." Biochem Biophys Res Commun 483 (2017): 430–34. 10.1016/j.bbrc.2016.12.126.

43. Chou, C. F., X. Zhu, Y. Y. Lin, K. L. Gamble, W. T. Garvey and C. Y. Chen. "Ksrp is critical in governing hepatic lipid metabolism through controlling per2 expression." J Lipid Res 56 (2015): 227–40. 10.1194/jlr.M050724.

44. Sun, S., F. Hanzawa, D. Kim, M. Umeki, S. Nakajima, K. Sakai, S. Ikeda, S. Mochizuki and H. Oda. "Circadian rhythm-dependent induction of hepatic lipogenic gene expression in rats fed a high-sucrose diet." J Biol Chem 294 (2019): 15206–17. 10.1074/jbc.RA119.010328.

45. Liu, J., P. Yi and F. Liu. "The effect of early time-restricted eating vs later time-restricted eating on weight loss and metabolic health." J Clin Endocrinol Metab 108 (2023): 1824–34. 10.1210/clinem/dgad036.

46. Martens, C. R., M. J. Rossman, M. R. Mazzo, L. R. Jankowski, E. E. Nagy, B. A. Denman, J. J. Richey, S. A. Johnson, B. P. Ziemba, Y. Wang, et al. "Short-term time-restricted feeding is safe and feasible in non-obese healthy midlife and older adults." Geroscience 42 (2020): 667–86. 10.1007/s11357-020-00156-6. https://www.ncbi.nlm.nih.gov/pubmed/31975053.

47. Poggiogalle, E., H. Jamshed and C. M. Peterson. "Circadian regulation of glucose, lipid, and energy metabolism in humans." Metabolism 84 (2018): 11–27. 10.1016/j.metabol.2017.11.017. https://www.ncbi.nlm.nih.gov/pubmed/29195759.

48. Gooley, J. J. "Circadian regulation of lipid metabolism." Proc Nutr Soc 75 (2016): 440–50. 10.1017/S0029665116000288. https://www.ncbi.nlm.nih.gov/pubmed/27225642.

49. Gooley, J. J. and E. C. Chua. "Diurnal regulation of lipid metabolism and applications of circadian lipidomics." J Genet Genomics 41 (2014): 231–50. 10.1016/j.jgg.2014.04.001.

50. Adamovich, Y., R. Aviram and G. Asher. "The emerging roles of lipids in circadian control." Biochim Biophys Acta 1851 (2015): 1017–25. 10.1016/j.bbalip.2014.11.013. https://www.ncbi.nlm.nih.gov/pubmed/25483623.

51. Gnocchi, D., M. Pedrelli, E. Hurt-Camejo and P. Parini. "Lipids around the clock: Focus on circadian rhythms and lipid metabolism." Biology (Basel) 4 (2015): 104–32. 10.3390/biology4010104. https://www.ncbi.nlm.nih.gov/pubmed/25665169.

52. Sinturel, F., W. Spaleniak and C. Dibner. "Circadian rhythm of lipid metabolism." Biochem Soc Trans 50 (2022): 1191–204. 10.1042/bst20210508.

53. Petrenko, V., F. Sinturel, H. Riezman and C. Dibner. "Lipid metabolism around the body clocks." Prog Lipid Res 91 (2023): 101235. 10.1016/j.plipres.2023.101235.

54. Sinturel, F., S. Chera, M. C. Brulhart-Meynet, J. P. Montoya, D. J. Stenvers, P. H. Bisschop, A. Kalsbeek, I. Guessous, F. R. Jornayvaz, J. Philippe, et al. "Circadian organization of lipid landscape is perturbed in type 2 diabetic patients." Cell Rep Med 4 (2023): 101299. 10.1016/j.xcrm.2023.101299. https://www.ncbi.nlm.nih.gov/pubmed/38016481.

55. Hilvo, M., P. J. Meikle, E. R. Pedersen, G. S. Tell, I. Dhar, H. Brenner, B. Schöttker, M. Lääperi, D. Kauhanen, K. M. Koistinen, et al. "Development and validation of a ceramide- and phospholipid-based cardiovascular risk estimation score for coronary artery disease patients." Eur Heart J 41 (2020): 371–80. 10.1093/eurheartj/ehz387.

56. Fretts, A. M., P. N. Jensen, A. Hoofnagle, B. McKnight, B. V. Howard, J. Umans, C. Yu, C. Sitlani, D. S. Siscovick, I. B. King, et al. "Plasma ceramide species are associated with diabetes risk in participants of the strong heart study." J Nutr 150 (2020): 1214–22. 10.1093/jn/nxz259.

57. Peterson, L. R., V. Xanthakis, M. S. Duncan, S. Gross, N. Friedrich, H. Völzke, S. B. Felix, H. Jiang, R. Sidhu, M. Nauck, et al. "Ceramide remodeling and risk of cardiovascular events and mortality." J Am Heart Assoc 7 (2018): 10.1161/jaha.117.007931.

58. Lemaitre, R. N., C. Yu, A. Hoofnagle, N. Hari, P. N. Jensen, A. M. Fretts, J. G. Umans, B. V. Howard, C. M. Sitlani, D. S. Siscovick, et al. "Circulating sphingolipids, insulin, homa-ir, and homa-b: The strong heart family study." Diabetes 67 (2018): 1663–72. 10.2337/db17-1449.

59. Meeusen, J. W., L. J. Donato, S. C. Bryant, L. M. Baudhuin, P. B. Berger and A. S. Jaffe. "Plasma ceramides." Arterioscler Thromb Vasc Biol 38 (2018): 1933–39. 10.1161/atvbaha.118.311199.

60. Wittenbecher, C., R. Cuadrat, L. Johnston, F. Eichelmann, S. Jäger, O. Kuxhaus, M. Prada, M. F. Del Greco, A. A. Hicks, P. Hoffman, et al. "Dihydroceramide- and ceramide-profiling provides insights into human cardiometabolic disease etiology." Nat Commun 13 (2022): 936. 10.1038/s41467-022-28496-1.

61. Hammerschmidt, P. and J. C. Brüning. "Contribution of specific ceramides to obesity-associated metabolic diseases." Cell Mol Life Sci 79 (2022): 395. 10.1007/s00018-022-04401-3.

62. Havulinna, A. S., M. Sysi-Aho, M. Hilvo, D. Kauhanen, R. Hurme, K. Ekroos, V. Salomaa and R. Laaksonen. "Circulating ceramides predict cardiovascular outcomes in the population-based finrisk 2002 cohort." Arterioscler Thromb Vasc Biol 36 (2016): 2424–30. 10.1161/atvbaha.116.307497.

63. Tarasov, K., K. Ekroos, M. Suoniemi, D. Kauhanen, T. Sylvänne, R. Hurme, I. Gouni-Berthold, H. K. Berthold, M. E. Kleber, R. Laaksonen, et al. "Molecular lipids identify cardiovascular risk and are efficiently lowered by simvastatin and pcsk9 deficiency." J Clin Endocrinol Metab 99 (2014): E45–52. 10.1210/jc.2013-2559.

64. Meikle, P. J., G. Wong, D. Tsorotes, C. K. Barlow, J. M. Weir, M. J. Christopher, G. L. MacIntosh, B. Goudey, L. Stern, A. Kowalczyk, et al. "Plasma lipidomic analysis of stable and unstable coronary artery disease." Arterioscler Thromb Vasc Biol 31 (2011): 2723–32. 10.1161/atvbaha.111.234096.

65. Fernandez, C., M. Sandin, J. L. Sampaio, P. Almgren, K. Narkiewicz, M. Hoffmann, T. Hedner, B. Wahlstrand, K. Simons, A. Shevchenko, et al. "Plasma lipid composition and risk of developing cardiovascular disease." PLoS One 8 (2013): e71846. 10.1371/journal.pone.0071846.

66. Summers, S. A. "Could ceramides become the new cholesterol?" Cell Metab 27 (2018): 276–80. 10.1016/j.cmet.2017.12.003.

67. Iliou, A., E. Mikros, I. Karaman, F. Elliott, J. L. Griffin, I. Tzoulaki and P. Elliott. "Metabolic phenotyping and cardiovascular disease: An overview of evidence from epidemiological settings." Heart 107 (2021): 1123–29. 10.1136/heartjnl-2019-315615. https://www.ncbi.nlm.nih.gov/pubmed/33608305.

68. Chen, H., Z. Wang, M. Qin, B. Zhang, L. Lin, Q. Ma, C. Liu, X. Chen, H. Li, W. Lai, et al. "Comprehensive metabolomics identified the prominent role of glycerophospholipid metabolism in coronary artery disease progression." Front Mol Biosci 8 (2021): 632950. 10.3389/fmolb.2021.632950. https://www.ncbi.nlm.nih.gov/pubmed/33937325.

69. Prada, M., F. Eichelmann, C. Wittenbecher, O. Kuxhaus and M. B. Schulze. "Plasma lipidomic n-6 polyunsaturated fatty acids and type 2 diabetes risk in the epic-potsdam prospective cohort study." Diabetes Care 46 (2023): 836–44. 10.2337/dc22-1435.

70. Forouhi, N. G., F. Imamura, S. J. Sharp, A. Koulman, M. B. Schulze, J. Zheng, Z. Ye, I. Sluijs, M. Guevara, J. M. Huerta, et al. "Association of plasma phospholipid n-3 and n-6 polyunsaturated fatty acids with type 2 diabetes: The epic-interact case-cohort study." PLoS Med 13 (2016): e1002094. 10.1371/journal.pmed.1002094.

71. Wolters, M., H. Schlenz, C. Börnhorst, P. Risé, C. Galli, L. A. Moreno, V. Pala, A. Siani, T. Veidebaum, M. Tornaritis, et al. "Desaturase activity is associated with weight status and metabolic risk markers in young children." J Clin Endocrinol Metab 100 (2015): 3760-9. 10.1210/jc.2015-2693.

72. Jacobs, S., J. Kröger, A. Floegel, H. Boeing, D. Drogan, T. Pischon, A. Fritsche, C. Prehn, J. Adamski, B. Isermann, et al. "Evaluation of various biomarkers as potential mediators of the association between coffee consumption and incident type 2 diabetes in the epic-potsdam study." Am J Clin Nutr 100 (2014): 891–900. 10.3945/ajcn.113.080317.

73. A, I. S. S., A. B. C and J. S. A. "Changes in plasma free fatty acids associated with type-2 diabetes." Nutrients 11 (2019): 10.3390/nu11092022. https://mdpi-res.com/d_attachment/nutrients/nutrients-11-02022/article_deploy/nutrients-11-02022-v2.pdf?version=1567592929.

74. Do, H. J., H. K. Chung, J. Moon and M. J. Shin. "Relationship between the estimates of desaturase activities and cardiometabolic phenotypes in koreans." J Clin Biochem Nutr 49 (2011): 131–5. 10.3164/jcbn.10-147.

75. Kawashima, A., S. Sugawara, M. Okita, T. Akahane, K. Fukui, M. Hashiuchi, C. Kataoka and I. Tsukamoto. "Plasma fatty acid composition, estimated desaturase activities, and intakes of energy and nutrient in japanese men with abdominal obesity or metabolic syndrome." J Nutr Sci Vitaminol (Tokyo) 55 (2009): 400–6. 10.3177/jnsv.55.400.

76. Domínguez-López, I., C. Arancibia-Riveros, A. Tresserra-Rimbau, S. Castro-Barquero, R. Casas, Z. Vázquez-Ruiz, E. Ros, M. Fitó, R. Estruch, M. C. López-Sabater, et al. "Relationship between estimated desaturase enzyme activity and metabolic syndrome in a longitudinal study." Front Nutr 9 (2022): 991277. 10.3389/fnut.2022.991277.

77. Svendsen, K., T. Olsen, T. C. Nordstrand Rusvik, S. M. Ulven, K. B. Holven, K. Retterstøl and V. H. Telle-Hansen. "Fatty acid profile and estimated desaturase activities in whole blood are associated with metabolic health." Lipids Health Dis 19 (2020): 102. 10.1186/s12944-020-01282-y.

78. Loizides-Mangold, U., L. Perrin, B. Vandereycken, J. A. Betts, J. P. Walhin, I. Templeman, S. Chanon, B. D. Weger, C. Durand, M. Robert, et al. "Lipidomics reveals diurnal lipid oscillations in human skeletal muscle persisting in cellular myotubes cultured in vitro." Proc Natl Acad Sci U S A 114 (2017): E8565–E74. 10.1073/pnas.1705821114. https://www.ncbi.nlm.nih.gov/pubmed/28973848.

79. Damiola, F., N. Le Minh, N. Preitner, B. Kornmann, F. Fleury-Olela and U. Schibler. "Restricted feeding uncouples circadian oscillators in peripheral tissues from the central pacemaker in the suprachiasmatic nucleus." Genes Dev 14 (2000): 2950–61. 10.1101/gad.183500.

80. Wehrens, S. M. T., S. Christou, C. Isherwood, B. Middleton, M. A. Gibbs, S. N. Archer, D. J. Skene and J. D. Johnston. "Meal timing regulates the human circadian system." Curr Biol 27 (2017): 1768–75 e3. 10.1016/j.cub.2017.04.059. https://www.ncbi.nlm.nih.gov/pubmed/28578930.

81. Koppold-Liebscher, D. A., C. Klatte, S. Demmrich, J. Schwarz, F. I. Kandil, N. Steckhan, R. Ring, C. S. Kessler, M. Jeitler, B. Koller, et al. "Effects of daytime dry fasting on hydration, glucose metabolism and circadian phase: A prospective exploratory cohort study in baha’i volunteers." Front Nutr 8 (2021): 662310. 10.3389/fnut.2021.662310. https://www.ncbi.nlm.nih.gov/pubmed/34395487.

82. Hui, D. Y. "Phospholipase a(2) enzymes in metabolic and cardiovascular diseases." Curr Opin Lipidol 23 (2012): 235–40. 10.1097/MOL.0b013e328351b439. https://www.ncbi.nlm.nih.gov/pubmed/22327613.

83. Deng, Y., X. Liu, Y. Sun, L. Zhou, Q. Li, Z. Lei, F. Yang, L. Chen, C. Zhang, W. Tan, et al. "Effects of time-restricted eating on intrahepatic fat and metabolic health among patients with nonalcoholic fatty liver disease." Obesity (Silver Spring) (2024): 10.1002/oby.23965.

84. Andriessen, C., C. E. Fealy, A. Veelen, S. M. M. van Beek, K. H. M. Roumans, N. J. Connell, J. Mevenkamp, E. Moonen-Kornips, B. Havekes, V. B. Schrauwen-Hinderling, et al. "Three weeks of time-restricted eating improves glucose homeostasis in adults with type 2 diabetes but does not improve insulin sensitivity: A randomised crossover trial." Diabetologia 65 (2022): 1710–20. 10.1007/s00125-022-05752-z. https://www.ncbi.nlm.nih.gov/pubmed/35871650.

85. Sutton, E. F., R. Beyl, K. S. Early, W. T. Cefalu, E. Ravussin and C. M. Peterson. "Early time-restricted feeding improves insulin sensitivity, blood pressure, and oxidative stress even without weight loss in men with prediabetes." Cell Metab 27 (2018): 1212–21 e3. 10.1016/j.cmet.2018.04.010. https://www.ncbi.nlm.nih.gov/pubmed/29754952.

86. Maruthur, N. M., S. J. Pilla, K. White, B. Wu, M. T. T. Maw, D. Duan, R. A. Turkson-Ocran, D. Zhao, J. Charleston, C. M. Peterson, et al. "Effect of isocaloric, time-restricted eating on body weight in adults with obesity : A randomized controlled trial." Ann Intern Med 177 (2024): 549–58. 10.7326/M23-3132. https://www.ncbi.nlm.nih.gov/pubmed/38639542.

87. Hopewell, S., A.-W. Chan, G. S. Collins, A. Hróbjartsson, D. Moher, K. F. Schulz, R. Tunn, R. Aggarwal, M. Berkwits, J. A. Berlin, et al. "Consort 2025 statement: Updated guideline for reporting randomised trials." Bmj 389 (2025): e081123. 10.1136/bmj-2024-081123. https://www.bmj.com/content/bmj/389/bmj-2024-081123.full.pdf.

88. Matyash, V., G. Liebisch, T. V. Kurzchalia, A. Shevchenko and D. Schwudke. "Lipid extraction by methyl-tert-butyl ether for high-throughput lipidomics." J Lipid Res 49 (2008): 1137–46. 10.1194/jlr.D700041-JLR200. https://www.ncbi.nlm.nih.gov/pubmed/18281723.

89. Kopczynski, D., C. S. Ejsing, J. G. McDonald, T. Bamba, E. S. Baker, J. Bertrand-Michel, B. Brugger, C. Coman, S. R. Ellis, T. J. Garrett, et al. "The lipidomics reporting checklist a framework for transparency of lipidomic experiments and repurposing resource data." J Lipid Res 65 (2024): 100621. 10.1016/j.jlr.2024.100621. https://www.ncbi.nlm.nih.gov/pubmed/39151590.

90. Aimo, L., R. Liechti, N. Hyka-Nouspikel, A. Niknejad, A. Gleizes, L. Gotz, D. Kuznetsov, F. P. David, F. G. van der Goot, H. Riezman, et al. "The swisslipids knowledgebase for lipid biology." Bioinformatics 31 (2015): 2860–6. 10.1093/bioinformatics/btv285. https://www.ncbi.nlm.nih.gov/pubmed/25943471.

91. Liebisch, G., E. Fahy, J. Aoki, E. A. Dennis, T. Durand, C. S. Ejsing, M. Fedorova, I. Feussner, W. J. Griffiths, H. Kofeler, et al. "Update on lipid maps classification, nomenclature, and shorthand notation for ms-derived lipid structures." J Lipid Res 61 (2020): 1539–55. 10.1194/jlr.S120001025. https://www.ncbi.nlm.nih.gov/pubmed/33037133.

92. Kanehisa, M. and S. Goto. "Kegg: Kyoto encyclopedia of genes and genomes." Nucleic Acids Res 28 (2000): 27–30. 10.1093/nar/28.1.27.

93. Kanehisa, M. "Toward understanding the origin and evolution of cellular organisms." Protein Sci 28 (2019): 1947–51. 10.1002/pro.3715.

94. Kanehisa, M., M. Furumichi, Y. Sato, Y. Matsuura and M. Ishiguro-Watanabe. "Kegg: Biological systems database as a model of the real world." Nucleic Acids Res 53 (2025): D672–d77. 10.1093/nar/gkae909.

95. Sherman, B. T., M. Hao, J. Qiu, X. Jiao, M. W. Baseler, H. C. Lane, T. Imamichi and W. Chang. "David: A web server for functional enrichment analysis and functional annotation of gene lists (2021 update)." Nucleic Acids Res 50 (2022): W216–w21. 10.1093/nar/gkac194.

96. Huang da, W., B. T. Sherman and R. A. Lempicki. "Systematic and integrative analysis of large gene lists using david bioinformatics resources." Nat Protoc 4 (2009): 44–57. 10.1038/nprot.2008.211.

97. Monfort-Pires, M., S. Lamichhane, C. Alonso, B. Egelandsdal, M. Orešič, V. O. Jordahl, O. Skjølsvold, I. Pérez-Ruiz, M. E. Blanco, S. Skeie, et al. "Classification of common food lipid sources regarding healthiness using advanced lipidomics: A four-arm crossover study." Int J Mol Sci 24 (2023): 10.3390/ijms24054941.

98. Al-Sari, N., S. Schmidt, T. Suvitaival, M. Kim, K. Trošt, A. G. Ranjan, M. B. Christensen, A. J. Overgaard, F. Pociot, K. Nørgaard, et al. "Changes in the lipidome in type 1 diabetes following low carbohydrate diet: Post-hoc analysis of a randomized crossover trial." Endocrinol Diabetes Metab 4 (2021): e00213. 10.1002/edm2.213.

