## Supplemental materials for "Impact of Intended Isocaloric Early vs. Late Time-Restricted Eating on Plasma Lipidome in Women with Overweight or Obesity: Secondary Analysis of the ChronoFast Trial"

**Table S1.** Inclusion and exclusion criteria.

| | |
|---|---|
| **Inclusion criteria** | Women |
| | Age 18–70 years |
| | BMI 25–35 kg/m$^2$ |
| **Exclusion criteria** | Shift work |
| | Travel across more than one time zone one month before a study or during the study period |
| | Weight changes of more than 5% in the last 3 months |
| | Pregnancy or breastfeeding |
| | Severe intestinal disease, bariatric surgery in the past |
| | Food allergies (individual inclusion possible after consultation with the doctor) |
| | Current special diets |
| | Poor quality of sleep (PSQI score < 10) |
| | Diabetes type 1 or type 2 |
| | Severe kidney disease |
| | Severe liver disease |
| | Cardiac infarction or stroke in the last 6 months |
| | Cancer cases in the last 2 years |
| | Glucocorticoid therapy (oral) |
| | Coagulation disorders |
| | Taking anticoagulant medication (inclusion if pausing medication is possible) |
| | Severe anaemia |
| | Systemic infections |
| | Severe psychiatric illnesses, addictions or depression |
| | Other diseases/surgeries/medications affecting glucose metabolism, appetite or immune function |

**Table S2.** Eating timing, food composition, and physical activity during study periods.

| | **Baseline** | **eTRE** | **lTRE** |
|---|---|---|---|
| Eating window duration | 11:48 (10:50 – 13:15) | 07:10 (6:53 – 7:30) | 07:07 (6:43 - 7:30) |
| Daily energy intake (kcal) | 2011.24 ± 302.57 | 1834.95 ± 262.36 * | 1915.83 ± 382.91 |
| Protein (EN%) | 15.45 ± 1.98 | 15.06 ± 1.90 | 15.44 ± 2.07 |
| Carbohydrates (EN%) | 42.86 (39.26 – 46.15) | 43.83 (41.33 – 46.20) | 43.33 (40.08 – 45.61) |
| Fat (EN%) | 39.75 (36.38 – 42.00) | 39.39 (36.41 – 42.71) | 39.39 (37.66 – 42.89) |
| Physical activity (MET) | 1.59 ± 0.15 | 1.56 ± 0.14 | 1.57 ± 0.12 |

Abbreviations: kcal, kilo calories; EN%, energy percent; MET, metabolic equivalent of task. For parameters with normal distribution mean and SD are reported, for parameters with non-normal distribution median and 25$^{th}$ IQR, and 75$^{th}$ IQR are reported. * p<0.001 in a paired T-test.

**Table S3.** Changes of the body weight, body composition, and blood lipids within eTRE and lTRE interventions.

| Characteristics | Before eTRE | After eTRE | Change eTRE after – before (95% CI) | P[a)] | Before lTRE | After lTRE | Change lTRE After – before (95% CI) | P[a)] | Difference between lTRE vs. eTRE (95% CI)[a] | P[a)] |
|---|---|---|---|---|---|---|---|---|---|---|
| BMI (kg/m$^2$) | 30.4 (2.9) | 30.0 (2.8) | -0.45 (-0.56 to -0.33) | **5.4x10$^{-9}$** | 30.3 (2.8) | 30.2 (2.9) | -0.12 (-0.21 to -0.03) | **0.01** | 0.33 (0.21 to 0.44) | **2.3x10$^{-6}$** |
| Weight (kg) | 82.3 (8.4) | 81.3 (8.0) | -1.08 (-1.40 to -0.77) | **9.1x10$^{-8}$** | 82.0 (8.1) | 81.5 (8.1) | -0.44 (-0.74 to -0.13) | **0.01** | 0.65 (0.27 to 1.03) | **0.002** |
| Fat mass (kg) | 34.6 (5.5) | 34.0 (5.8) | -0.61 (-1.01 to -0.22) | **0.002** | 34.1 (5.6) | 34.1 (5.5) | 0.02 (-0.36 to 0.39) | 0.92 | 0.65 (0.19 to 1.11) | **0.007** |
| Lean mass (kg) | 47.9 (4.5) | 47.4 (3.8) | -0.57 (-1.11 to -0.04) | **0.04** | 47.8 (4.3) | 47.5 (4.3) | -0.28 (-0.78 to 0.23) | 0.27 | 0.25 (-0.31 to 0.81) | 0.37 |
| Waist circumference (cm) | 99.1 (9.5) | 98.8 (8.2) | -0.36 (-1.65 to 0.92) | 0.57 | 98.9 (8.2) | 99.3 (8.3) | 0.48 (-0.97 to 1.92) | 0.51 | 0.84 (-1.21 to 2.89) | 0.41 |
| Total cholesterol, mmol/L | 5.54 (0.92) | 5.42 (0.83) | -0.12 (-0.32 to 0.08) | 0.24 | 5.73 (1.05) | 5.62 (0.92) | -0.12 (-0.30 to 0.06) | 0.19 | 0.001 (-0.28 to 0.28) | 0.94 |
| HDL cholesterol, mmol/L | 1.56 (0.38) | 1.46 (0.33) | -0.10 (-0.14 to -0.06) | **6.0x10$^{-5}$** | 1.57 (0.37) | 1.50 (0.38) | -0.07 (-0.11 to -0.03) | **0.003** | 0.03 (-0.04 to 0.09) | 0.45 |
| LDL cholesterol, mmol/L | 3.71 (1.12) | 3.92 (1.12) | 0.21 (-0.15 to 0.57) | 0.24 | 3.87 (1.26) | 4.09 (1.18) | 0.22 (-0.10 to 0.55) | 0.17 | 0.01 (-0.54 to 0.56) | 0.96 |
| Triglycerides, mmol/L | 1.29 (0.53) | 1.36 (0.58) | 0.06 (-0.08 to 0.21) | 0.27 | 1.36 (0.61) | 1.35 (0.70) | -0.01 (-0.15 to 0.13) | 0.56 | -0.07 (-0.22 to 0.08) | 0.34 |

Data are shown as mean (SD) or mean (95% CI).
[a)] Comparison by paired Student's t-test

**Table S4.** All identified lipid classes and species.

A. **Classes**

| |
|---|
| cholesterol (Chol) |
| cholesterol esters (CE) |
| triacylglycerols (TAG) |
| diacylglycerols (DAG) |
| phosphatidylcholines (PCs |
| phosphatidylcholine ethers (PC O-) |
| phosphatidylethanolamines (PE) |
| phosphatidylethanolamine ethers (PE O-) |
| phosphatidylinositols (PI) |
| lysophosphatidylcholines (LPC) |
| lysophosphatidylcholine ethers (LPC O-) |
| lysophosphatidylethanolamines (LPE) |
| sphingomyelins (SM) |
| ceramides (CER) |

B. **Species**

| **feature** | **Shorthand Notation** | **SwissLipids Name** | **SwissLipids ID** |
|---|---|---|---|
| CE 14:0;0 | SE 27:1/14:0 | tetradecanoyl-cholesterol | SLM:000500261 |
| CE 15:0;0 | SE 27:1/15:0 | pentadecanoyl-cholesterol | SLM:000500263 |
| CE 16:0;0 | SE 27:1/16:0 | hexadecanoyl-cholesterol | SLM:000389778 |
| CE 16:1;0 | SE 27:1/16:1 | Sterol ester (27:1/16:1) | SLM:000500345 |
| CE 17:0;0 | SE 27:1/17:0 | heptadecanoyl-cholesterol | SLM:000500268 |
| CE 17:1;0 | SE 27:1/17:1 | Sterol ester (27:1/17:1) | |
| CE 18:0;0 | SE 27:1/18:0 | octadecanoyl-cholesterol | SLM:000500276 |
| CE 18:1;0 | SE 27:1/18:1 | Sterol ester (27:1/18:1) | SLM:000500351 |
| CE 18:2;0 | SE 27:1/18:2 | Sterol ester (27:1/18:2) | SLM:000500350 |
| CE 18:3;0 | SE 27:1/18:3 | Sterol ester (27:1/18:3) | SLM:000500349 |
| CE 20:2;0 | SE 27:1/20:2 | Sterol ester (27:1/20:2) | SLM:000500357 |
| CE 20:3;0 | SE 27:1/20:3 | Sterol ester (27:1/20:3) | SLM:000500356 |

| | | | |
|---|---|---|---|
| CE 20:4;0 | SE 27:1/20:4 | Sterol ester (27:1/20:4) | SLM:000500355 |
| CE 20:5;0 | SE 27:1/20:5 | Sterol ester (27:1/20:5) | SLM:000500354 |
| CE 22:6;0 | SE 27:1/22:6 | Sterol ester (27:1/22:6) | SLM:000500361 |
| Cer 40:0;2 | Cer 40:0;O2 | Ceramide (d40:0) | SLM:000391322 |
| Cer 40:1;2 | Cer 40:1;O2 | Ceramide (d40:1) | SLM:000391319 |
| Cer 40:2;2 | Cer 40:2;O2 | Ceramide (d40:2) | SLM:000391317 |
| Cer 42:0;2 | Cer 42:0;O2 | Ceramide (d42:0) | SLM:000391349 |
| Cer 42:1;2 | Cer 42:1;O2 | Ceramide (d42:1) | SLM:000391346 |
| Cer 42:2;2 | Cer 42:2;O2 | Ceramide (d42:2) | SLM:000391345 |
| Chol | ST 27:1;O | cholesterol | SLM:000000287 |
| DAG 16:0;0_18:1;0 | DG 16:0_18:1 | Diacylglycerol (16:0_18:1) | SLM:000308862 |
| DAG 16:0;0_18:2;0 | DG 16:0_18:2 | Diacylglycerol (16:0_18:2) | SLM:000308863 |
| DAG 16:1;0_18:1;0 | DG 16:1_18:1 | Diacylglycerol (16:1_18:1) | SLM:000308894 |
| DAG 18:1;0_18:1;0 | DG 18:1_18:1 | Diacylglycerol (18:1_18:1) | SLM:000309012 |
| DAG 18:1;0_18:2;0 | DG 18:1_18:2 | Diacylglycerol (18:1_18:2) | SLM:000309013 |
| DAG 18:1;0_18:3;0 | DG 18:1_18:3 | Diacylglycerol (18:1_18:3) | SLM:000309014 |
| HexCer 40:1;2 | HexCer 40:1;O2 | Hexosyl ceramide (d40:1) | SLM:000390387 |
| HexCer 42:1;2 | HexCer 42:1;O2 | Hexosyl ceramide (d42:1) | SLM:000390412 |
| HexCer 42:2;2 | HexCer 42:2;O2 | Hexosyl ceramide (d42:2) | SLM:000390411 |
| LPC 16:0;0 | LPC 16:0 | Phosphatidylcholine (16:0_0:0) | SLM:000063723 |
| LPC 16:1;0 | LPC 16:1 | Phosphatidylcholine (16:1_0:0) | SLM:000063777 |
| LPC 18:0;0 | LPC 18:0 | Phosphatidylcholine (18:0_0:0) | SLM:000063933 |
| LPC 18:1;0 | LPC 18:1 | Phosphatidylcholine (18:1_0:0) | SLM:000063983 |
| LPC 18:2;0 | LPC 18:2 | Phosphatidylcholine (18:2_0:0) | SLM:000064032 |
| LPC 20:3;0 | LPC 20:3 | Phosphatidylcholine (20:3_0:0) | SLM:000064347 |
| LPC 20:4;0 | LPC 20:4 | Phosphatidylcholine (20:4_0:0) | SLM:000064388 |
| LPE 16:0;0 | LPE 16:0 | Phosphatidylethanolamine (16:0_0:0) | SLM:000067687 |
| LPE 18:0;0 | LPE 18:0 | Phosphatidylethanolamine (18:0_0:0) | SLM:000067897 |
| LPE 18:1;0 | LPE 18:1 | Phosphatidylethanolamine (18:1_0:0) | SLM:000067947 |

| | | | |
|---|---|---|---|
| LPE 18:2;0 | LPE 18:2 | Phosphatidylethanolamine (18:2_0:0) | SLM:000067996 |
| LPE 20:0;0 | LPE 20:0 | Phosphatidylethanolamine (20:0_0:0) | SLM:000068182 |
| LPE 20:1;0 | LPE 20:1 | Phosphatidylethanolamine (20:1_0:0) | SLM:000068226 |
| LPE 20:2;0 | LPE 20:2 | Phosphatidylethanolamine (20:2_0:0) | SLM:000068269 |
| LPE 20:4;0 | LPE 20:4 | Phosphatidylethanolamine (20:4_0:0) | SLM:000068352 |
| LPE 22:6;0 | LPE 22:6 | Phosphatidylethanolamine (22:6_0:0) | SLM:000068676 |
| PC 14:0;0_16:0;0 | PC 14:0_16:0 | Phosphatidylcholine (14:0_16:0) | SLM:000063559 |
| PC 14:0;0_18:0;0 | PC 14:0_18:0 | Phosphatidylcholine (14:0_18:0) | SLM:000063563 |
| PC 14:0;0_18:1;0 | PC 14:0_18:1 | Phosphatidylcholine (14:0_18:1) | SLM:000063564 |
| PC 14:0;0_18:2;0 | PC 14:0_18:2 | Phosphatidylcholine (14:0_18:2) | SLM:000063565 |
| PC 14:0;0_20:3;0 | PC 14:0_20:3 | Phosphatidylcholine (14:0_20:3) | SLM:000063572 |
| PC 14:0;0_20:4;0 | PC 14:0_20:4 | Phosphatidylcholine (14:0_20:4) | SLM:000063573 |
| PC 15:0;0_16:0;0 | PC 15:0_16:0 | Phosphatidylcholine (15:0_16:0) | SLM:000063670 |
| PC 15:0;0_17:0;0 | PC 15:0_17:0 | Phosphatidylcholine (15:0_17:0) | SLM:000063673 |
| PC 15:0;0_18:1;0 | PC 15:0_18:1 | Phosphatidylcholine (15:0_18:1) | SLM:000063675 |
| PC 15:0;0_18:2;0 | PC 15:0_18:2 | Phosphatidylcholine (15:0_18:2) | SLM:000063676 |
| PC 15:0;0_20:3;0 | PC 15:0_20:3 | Phosphatidylcholine (15:0_20:3) | SLM:000063683 |
| PC 15:0;0_20:4;0 | PC 15:0_20:4 | Phosphatidylcholine (15:0_20:4) | SLM:000063684 |
| PC 16:0;0_16:0;0 | PC 16:0_16:0 | Phosphatidylcholine (16:0/16:0) | SLM:000088143 |
| PC 16:0;0_16:1;0 | PC 16:0_16:1 | Phosphatidylcholine (16:0_16:1) | SLM:000063725 |
| PC 16:0;0_17:1;0 | PC 16:0_17:1 | Phosphatidylcholine (16:0_17:1) | |
| PC 16:0;0_18:0;0 | PC 16:0_18:0 | Phosphatidylcholine (16:0_18:0) | SLM:000063728 |
| PC 16:0;0_18:1;0 | PC 16:0_18:1 | Phosphatidylcholine (16:0_18:1) | SLM:000063729 |
| PC 16:0;0_18:2;0 | PC 16:0_18:2 | Phosphatidylcholine (16:0_18:2) | SLM:000063730 |
| PC 16:0;0_18:3;0 | PC 16:0_18:3 | Phosphatidylcholine (16:0_18:3) | SLM:000063731 |
| PC 16:0;0_19:1;0 | PC 16:0_19:1 | Phosphatidylcholine (16:0_19:1) | |
| PC 16:0;0_20:1;0 | PC 16:0_20:1 | Phosphatidylcholine (16:0_20:1) | SLM:000063735 |
| PC 16:0;0_20:2;0 | PC 16:0_20:2 | Phosphatidylcholine (16:0_20:2) | SLM:000063736 |

| | | | |
|---|---|---|---|
| PC 16:0;0_20:3;0 | PC 16:0_20:3 | Phosphatidylcholine (16:0_20:3) | SLM:000063737 |
| PC 16:0;0_20:4;0 | PC 16:0_20:4 | Phosphatidylcholine (16:0_20:4) | SLM:000063738 |
| PC 16:0;0_20:5;0 | PC 16:0_20:5 | Phosphatidylcholine (16:0_20:5) | SLM:000063739 |
| PC 16:0;0_22:4;0 | PC 16:0_22:4 | Phosphatidylcholine (16:0_22:4) | SLM:000063745 |
| PC 16:0;0_22:5;0 | PC 16:0_22:5 | Phosphatidylcholine (16:0_22:5) | SLM:000063746 |
| PC 16:0;0_22:6;0 | PC 16:0_22:6 | Phosphatidylcholine (16:0_22:6) | SLM:000063747 |
| PC 16:1;0_16:1;0 | PC 16:1_16:1 | Phosphatidylcholine (16:1/16:1) | SLM:000088209 |
| PC 16:1;0_17:0;0 | PC 16:1_17:0 | Phosphatidylcholine (16:1_17:0) | SLM:000063780 |
| PC 16:1;0_18:0;0 | PC 16:1_18:0 | Phosphatidylcholine (16:1_18:0) | SLM:000063781 |
| PC 16:1;0_18:1;0 | PC 16:1_18:1 | Phosphatidylcholine (16:1_18:1) | SLM:000063782 |
| PC 16:1;0_18:2;0 | PC 16:1_18:2 | Phosphatidylcholine (16:1_18:2) | SLM:000063783 |
| PC 16:1;0_20:3;0 | PC 16:1_20:3 | Phosphatidylcholine (16:1_20:3) | SLM:000063790 |
| PC 16:1;0_20:4;0 | PC 16:1_20:4 | Phosphatidylcholine (16:1_20:4) | SLM:000063791 |
| PC 17:0;0_18:1;0 | PC 17:0_18:1 | Phosphatidylcholine (17:0_18:1) | SLM:000063885 |
| PC 17:0;0_18:2;0 | PC 17:0_18:2 | Phosphatidylcholine (17:0_18:2) | SLM:000063886 |
| PC 17:0;0_18:3;0 | PC 17:0_18:3 | Phosphatidylcholine (17:0_18:3) | SLM:000063887 |
| PC 17:0;0_20:3;0 | PC 17:0_20:3 | Phosphatidylcholine (17:0_20:3) | SLM:000063893 |
| PC 17:0;0_20:4;0 | PC 17:0_20:4 | Phosphatidylcholine (17:0_20:4) | SLM:000063894 |
| PC 17:0;0_20:5;0 | PC 17:0_20:5 | Phosphatidylcholine (17:0_20:5) | SLM:000063895 |
| PC 17:0;0_20:6;0 | PC 17:0_20:6 | Phosphatidylcholine (17:0_20:6) | |
| PC 17:0;0_22:4;0 | PC 17:0_22:4 | Phosphatidylcholine (17:0_22:4) | SLM:000063901 |
| PC 17:0;0_22:5;0 | PC 17:0_22:5 | Phosphatidylcholine (17:0_22:5) | SLM:000063902 |
| PC 17:0;0_22:6;0 | PC 17:0_22:6 | Phosphatidylcholine (17:0_22:6) | SLM:000063903 |
| PC 17:1;0_18:0;0 | PC 17:1_18:0 | Phosphatidylcholine (17:1_18:0) | |
| PC 17:1;0_18:1;0 | PC 17:1_18:1 | Phosphatidylcholine (17:1_18:1) | |
| PC 17:1;0_18:2;0 | PC 17:1_18:2 | Phosphatidylcholine (17:1_18:2) | |
| PC 18:0;0_18:1;0 | PC 18:0_18:1 | Phosphatidylcholine (18:0_18:1) | SLM:000063935 |
| PC 18:0;0_18:2;0 | PC 18:0_18:2 | Phosphatidylcholine (18:0_18:2) | SLM:000063936 |
| PC 18:0;0_18:3;0 | PC 18:0_18:3 | Phosphatidylcholine (18:0_18:3) | SLM:000063937 |
| PC 18:0;0_20:2;0 | PC 18:0_20:2 | Phosphatidylcholine (18:0_20:2) | SLM:000063942 |
| PC 18:0;0_20:3;0 | PC 18:0_20:3 | Phosphatidylcholine (18:0_20:3) | SLM:000063943 |

| | | | |
|---|---|---|---|
| PC 18:0;0_20:4;0 | PC 18:0_20:4 | Phosphatidylcholine (18:0_20:4) | SLM:000063944 |
| PC 18:0;0_20:5;0 | PC 18:0_20:5 | Phosphatidylcholine (18:0_20:5) | SLM:000063945 |
| PC 18:0;0_22:4;0 | PC 18:0_22:4 | Phosphatidylcholine (18:0_22:4) | SLM:000063951 |
| PC 18:0;0_22:5;0 | PC 18:0_22:5 | Phosphatidylcholine (18:0_22:5) | SLM:000063952 |
| PC 18:0;0_22:6;0 | PC 18:0_22:6 | Phosphatidylcholine (18:0_22:6) | SLM:000063953 |
| PC 18:1;0_18:1;0 | PC 18:1_18:1 | Phosphatidylcholine (18:1/18:1) | SLM:000088473 |
| PC 18:1;0_18:2;0 | PC 18:1_18:2 | Phosphatidylcholine (18:1_18:2) | SLM:000063985 |
| PC 18:1;0_18:3;0 | PC 18:1_18:3 | Phosphatidylcholine (18:1_18:3) | SLM:000063986 |
| PC 18:1;0_20:1;0 | PC 18:1_20:1 | Phosphatidylcholine (18:1_20:1) | SLM:000063990 |
| PC 18:1;0_20:2;0 | PC 18:1_20:2 | Phosphatidylcholine (18:1_20:2) | SLM:000063991 |
| PC 18:1;0_20:3;0 | PC 18:1_20:3 | Phosphatidylcholine (18:1_20:3) | SLM:000063992 |
| PC 18:1;0_20:4;0 | PC 18:1_20:4 | Phosphatidylcholine (18:1_20:4) | SLM:000063993 |
| PC 18:1;0_20:5;0 | PC 18:1_20:5 | Phosphatidylcholine (18:1_20:5) | SLM:000063994 |
| PC 18:1;0_22:5;0 | PC 18:1_22:5 | Phosphatidylcholine (18:1_22:5) | SLM:000064001 |
| PC 18:1;0_22:6;0 | PC 18:1_22:6 | Phosphatidylcholine (18:1_22:6) | SLM:000064002 |
| PC 18:2;0_18:2;0 | PC 18:2_18:2 | Phosphatidylcholine (18:2/18:2) | SLM:000088539 |
| PC 18:2;0_18:3;0 | PC 18:2_18:3 | Phosphatidylcholine (18:2_18:3) | SLM:000064034 |
| PC 18:2;0_19:0;0 | PC 18:2_19:0 | Phosphatidylcholine (18:2_19:0) | SLM:000064036 |
| PC 18:2;0_20:0;0 | PC 18:2_20:0 | Phosphatidylcholine (18:2_20:0) | SLM:000064037 |
| PC 18:2;0_20:1;0 | PC 18:2_20:1 | Phosphatidylcholine (18:2_20:1) | SLM:000064038 |
| PC 18:2;0_20:2;0 | PC 18:2_20:2 | Phosphatidylcholine (18:2_20:2) | SLM:000064039 |
| PC 18:2;0_20:3;0 | PC 18:2_20:3 | Phosphatidylcholine (18:2_20:3) | SLM:000064040 |
| PC 18:2;0_20:4;0 | PC 18:2_20:4 | Phosphatidylcholine (18:2_20:4) | SLM:000064041 |
| PC 20:0;0_20:4;0 | PC 20:0_20:4 | Phosphatidylcholine (20:0_20:4) | SLM:000064223 |
| PC 20:1;0_20:4;0 | PC 20:1_20:4 | Phosphatidylcholine (20:1_20:4) | SLM:000064266 |
| PC 20:3;0_20:4;0 | PC 20:3_20:4 | Phosphatidylcholine (20:3_20:4) | SLM:000064349 |
| PC O-16:0;0/16:0;0 | PC O-16:0/16:0 | Phosphatidylcholine (O-16:0/16:0) | SLM:000092108 |
| PC O-16:0;0/16:1;0 | PC O-16:0/16:1 | Phosphatidylcholine (O-16:0/16:1) | SLM:000092109 |
| PC O-16:0;0/18:0;0 | PC O-16:0/18:0 | Phosphatidylcholine (O-16:0/18:0) | SLM:000092112 |
| PC O-16:0;0/18:1;0 | PC O-16:0/18:1 | Phosphatidylcholine (O-16:0/18:1) | SLM:000092113 |
| PC O-16:0;0/18:2;0 | PC O-16:0/18:2 | Phosphatidylcholine (O-16:0/18:2) | SLM:000092114 |

| | | | |
|---|---|---|---|
| PC O-16:0;0/18:3;0 | PC O-16:0/18:3 | Phosphatidylcholine (O-16:0/18:3) | SLM:000092115 |
| PC O-16:0;0/20:3;0 | PC O-16:0/20:3 | Phosphatidylcholine (O-16:0/20:3) | SLM:000092121 |
| PC O-16:0;0/20:4;0 | PC O-16:0/20:4 | Phosphatidylcholine (O-16:0/20:4) | SLM:000092122 |
| PC O-16:0;0/20:5;0 | PC O-16:0/20:5 | Phosphatidylcholine (O-16:0/20:5) | SLM:000092123 |
| PC O-16:0;0/22:4;0 | PC O-16:0/22:4 | Phosphatidylcholine (O-16:0/22:4) | SLM:000092129 |
| PC O-16:0;0/22:5;0 | PC O-16:0/22:5 | Phosphatidylcholine (O-16:0/22:5) | SLM:000092130 |
| PC O-16:0;0/22:6;0 | PC O-16:0/22:6 | Phosphatidylcholine (O-16:0/22:6) | SLM:000092131 |
| PC O-16:1;0/16:0;0 | PC O-16:1/16:0 | Phosphatidylcholine (O-16:1_16:0) | SLM:000065984 |
| PC O-16:1;0/16:1;0 | PC O-16:1/16:1 | Phosphatidylcholine (O-16:1_16:1) | SLM:000065985 |
| PC O-16:1;0/18:0;0 | PC O-16:1/18:0 | Phosphatidylcholine (O-16:1_18:0) | SLM:000065988 |
| PC O-16:1;0/18:1;0 | PC O-16:1/18:1 | Phosphatidylcholine (O-16:1_18:1) | SLM:000065989 |
| PC O-16:1;0/18:2;0 | PC O-16:1/18:2 | Phosphatidylcholine (O-16:1_18:2) | SLM:000065990 |
| PC O-16:1;0/20:3;0 | PC O-16:1/20:3 | Phosphatidylcholine (O-16:1_20:3) | SLM:000065997 |
| PC O-16:1;0/20:4;0 | PC O-16:1/20:4 | Phosphatidylcholine (O-16:1_20:4) | SLM:000065998 |
| PC O-16:1;0/22:4;0 | PC O-16:1/22:4 | Phosphatidylcholine (O-16:1_22:4) | SLM:000066005 |
| PC O-16:1;0/22:5;0 | PC O-16:1/22:5 | Phosphatidylcholine (O-16:1_22:5) | SLM:000066006 |
| PC O-16:2;0/16:0;0 | PC O-16:2/16:0 | Phosphatidylcholine (O-16:2_16:0) | |
| PC O-16:2;0/18:0;0 | PC O-16:2/18:0 | Phosphatidylcholine (O-16:2_18:0) | |
| PC O-16:2;0/18:1;0 | PC O-16:2/18:1 | Phosphatidylcholine (O-16:2_18:1) | |
| PC O-17:0;0/15:0;0 | PC O-17:0/15:0 | Phosphatidylcholine (O-17:0/15:0) | SLM:000092172 |
| PC O-17:0;0/17:0;0 | PC O-17:0/17:0 | Phosphatidylcholine (O-17:0/17:0) | SLM:000092176 |
| PC O-17:0;0/17:1;0 | PC O-17:0/17:1 | Phosphatidylcholine (O-17:0/17:1) | |
| PC O-17:1;0/17:0;0 | PC O-17:1/17:0 | Phosphatidylcholine (O-17:1_17:0) | SLM:000066117 |
| PC O-17:2;0/17:0;0 | PC O-17:2/17:0 | Phosphatidylcholine (O-17:2_17:0) | |
| PC O-18:0;0/14:0;0 | PC O-18:0/14:0 | Phosphatidylcholine (O-18:0/14:0) | SLM:000092235 |
| PC O-18:0;0/16:0;0 | PC O-18:0/16:0 | Phosphatidylcholine (O-18:0/16:0) | SLM:000092238 |
| PC O-18:0;0/16:1;0 | PC O-18:0/16:1 | Phosphatidylcholine (O-18:0/16:1) | SLM:000092239 |
| PC O-18:0;0/18:2;0 | PC O-18:0/18:2 | Phosphatidylcholine (O-18:0/18:2) | SLM:000092244 |
| PC O-18:0;0/18:3;0 | PC O-18:0/18:3 | Phosphatidylcholine (O-18:0/18:3) | SLM:000092245 |
| PC O-18:0;0/20:4;0 | PC O-18:0/20:4 | Phosphatidylcholine (O-18:0/20:4) | SLM:000092252 |
| PC O-18:0;0/20:5;0 | PC O-18:0/20:5 | Phosphatidylcholine (O-18:0/20:5) | SLM:000092253 |

| | | | |
|---|---|---|---|
| PC O-18:0;0/20:6;0 | PC O-18:0/20:6 | Phosphatidylcholine (O-18:0/20:6) | |
| PC O-18:0;0/22:5;0 | PC O-18:0/22:5 | Phosphatidylcholine (O-18:0/22:5) | SLM:000092260 |
| PC O-18:0;0/22:6;0 | PC O-18:0/22:6 | Phosphatidylcholine (O-18:0/22:6) | SLM:000092261 |
| PC O-18:1;0/16:0;0 | PC O-18:1/16:0 | Phosphatidylcholine (O-18:1_16:0) | SLM:000066244 |
| PC O-18:1;0/16:1;0 | PC O-18:1/16:1 | Phosphatidylcholine (O-18:1_16:1) | SLM:000066245 |
| PC O-18:1;0/18:1;0 | PC O-18:1/18:1 | Phosphatidylcholine (O-18:1_18:1) | SLM:000066249 |
| PC O-18:1;0/18:2;0 | PC O-18:1/18:2 | Phosphatidylcholine (O-18:1_18:2) | SLM:000066250 |
| PC O-18:1;0/20:3;0 | PC O-18:1/20:3 | Phosphatidylcholine (O-18:1_20:3) | SLM:000066257 |
| PC O-18:1;0/20:4;0 | PC O-18:1/20:4 | Phosphatidylcholine (O-18:1_20:4) | SLM:000066258 |
| PC O-18:1;0/22:4;0 | PC O-18:1/22:4 | Phosphatidylcholine (O-18:1_22:4) | SLM:000066265 |
| PC O-18:1;0/22:5;0 | PC O-18:1/22:5 | Phosphatidylcholine (O-18:1_22:5) | SLM:000066266 |
| PC O-18:2;0/16:0;0 | PC O-18:2/16:0 | Phosphatidylcholine (O-18:2_16:0) | SLM:000066309 |
| PC O-18:2;0/18:0;0 | PC O-18:2/18:0 | Phosphatidylcholine (O-18:2_18:0) | SLM:000066313 |
| PC O-18:2;0/18:1;0 | PC O-18:2/18:1 | Phosphatidylcholine (O-18:2_18:1) | SLM:000066314 |
| PC O-18:2;0/18:2;0 | PC O-18:2/18:2 | Phosphatidylcholine (O-18:2_18:2) | SLM:000066315 |
| PC O-18:2;0/20:3;0 | PC O-18:2/20:3 | Phosphatidylcholine (O-18:2_20:3) | SLM:000066322 |
| PC O-18:2;0/20:4;0 | PC O-18:2/20:4 | Phosphatidylcholine (O-18:2_20:4) | SLM:000066323 |
| PE 16:0;0_18:1;0 | PE 16:0_18:1 | Phosphatidylethanolamine (16:0_18:1) | SLM:000067693 |
| PE 16:0;0_18:2;0 | PE 16:0_18:2 | Phosphatidylethanolamine (16:0_18:2) | SLM:000067694 |
| PE 16:0;0_20:3;0 | PE 16:0_20:3 | Phosphatidylethanolamine (16:0_20:3) | SLM:000067701 |
| PE 16:0;0_20:4;0 | PE 16:0_20:4 | Phosphatidylethanolamine (16:0_20:4) | SLM:000067702 |
| PE 16:0;0_22:5;0 | PE 16:0_22:5 | Phosphatidylethanolamine (16:0_22:5) | SLM:000067710 |
| PE 16:0;0_22:6;0 | PE 16:0_22:6 | Phosphatidylethanolamine (16:0_22:6) | SLM:000067711 |
| PE 16:1;0_18:0;0 | PE 16:1_18:0 | Phosphatidylethanolamine (16:1_18:0) | SLM:000067745 |
| PE 18:0;0_18:1;0 | PE 18:0_18:1 | Phosphatidylethanolamine (18:0_18:1) | SLM:000067899 |
| PE 18:0;0_18:2;0 | PE 18:0_18:2 | Phosphatidylethanolamine (18:0_18:2) | SLM:000067900 |

| | | | |
|---|---|---|---|
| PE 18:0;0_18:3;0 | PE 18:0_18:3 | Phosphatidylethanolamine (18:0_18:3) | SLM:000067901 |
| PE 18:0;0_20:2;0 | PE 18:0_20:2 | Phosphatidylethanolamine (18:0_20:2) | SLM:000067906 |
| PE 18:0;0_20:3;0 | PE 18:0_20:3 | Phosphatidylethanolamine (18:0_20:3) | SLM:000067907 |
| PE 18:0;0_20:4;0 | PE 18:0_20:4 | Phosphatidylethanolamine (18:0_20:4) | SLM:000067908 |
| PE 18:0;0_20:5;0 | PE 18:0_20:5 | Phosphatidylethanolamine (18:0_20:5) | SLM:000067909 |
| PE 18:0;0_22:4;0 | PE 18:0_22:4 | Phosphatidylethanolamine (18:0_22:4) | SLM:000067915 |
| PE 18:0;0_22:5;0 | PE 18:0_22:5 | Phosphatidylethanolamine (18:0_22:5) | SLM:000067916 |
| PE 18:0;0_22:6;0 | PE 18:0_22:6 | Phosphatidylethanolamine (18:0_22:6) | SLM:000067917 |
| PE 18:1;0_18:1;0 | PE 18:1_18:1 | Phosphatidylethanolamine (18:1/18:1) | SLM:000094777 |
| PE 18:1;0_18:2;0 | PE 18:1_18:2 | Phosphatidylethanolamine (18:1_18:2) | SLM:000067949 |
| PE 18:1;0_20:3;0 | PE 18:1_20:3 | Phosphatidylethanolamine (18:1_20:3) | SLM:000067956 |
| PE 18:1;0_20:4;0 | PE 18:1_20:4 | Phosphatidylethanolamine (18:1_20:4) | SLM:000067957 |
| PE 18:2;0_18:2;0 | PE 18:2_18:2 | Phosphatidylethanolamine (18:2/18:2) | SLM:000094843 |
| PE O-16:0;0/18:2;0 | PE O-16:0/18:2 | Phosphatidylethanolamine (O-16:0/18:2) | SLM:000098418 |
| PE O-16:0;0/20:4;0 | PE O-16:0/20:4 | Phosphatidylethanolamine (O-16:0/20:4) | SLM:000098426 |
| PE O-16:0;0/22:5;0 | PE O-16:0/22:5 | Phosphatidylethanolamine (O-16:0/22:5) | SLM:000098434 |
| PE O-16:1;0/18:1;0 | PE O-16:1/18:1 | Phosphatidylethanolamine (O-16:1_18:1) | SLM:000069953 |
| PE O-16:1;0/18:2;0 | PE O-16:1/18:2 | Phosphatidylethanolamine (O-16:1_18:2) | SLM:000069954 |
| PE O-16:1;0/20:3;0 | PE O-16:1/20:3 | Phosphatidylethanolamine (O-16:1_20:3) | SLM:000069961 |
| PE O-16:1;0/20:4;0 | PE O-16:1/20:4 | Phosphatidylethanolamine | SLM:000069962 |

| | | | |
|---|---|---|---|
| | | (O-16:1_20:4) | |
| PE O-16:1;0/22:4;0 | PE O-16:1/22:4 | Phosphatidylethanolamine (O-16:1_22:4) | SLM:000069969 |
| PE O-16:1;0/22:5;0 | PE O-16:1/22:5 | Phosphatidylethanolamine (O-16:1_22:5) | SLM:000069970 |
| PE O-16:1;0/22:6;0 | PE O-16:1/22:6 | Phosphatidylethanolamine (O-16:1_22:6) | SLM:000069971 |
| PE O-16:2;0/18:0;0 | PE O-16:2/18:0 | Phosphatidylethanolamine (O-16:2_18:0) | |
| PE O-18:0;0/18:2;0 | PE O-18:0/18:2 | Phosphatidylethanolamine (O-18:0/18:2) | SLM:000098548 |
| PE O-18:0;0/20:4;0 | PE O-18:0/20:4 | Phosphatidylethanolamine (O-18:0/20:4) | SLM:000098556 |
| PE O-18:1;0/18:1;0 | PE O-18:1/18:1 | Phosphatidylethanolamine (O-18:1_18:1) | SLM:000070213 |
| PE O-18:1;0/18:2;0 | PE O-18:1/18:2 | Phosphatidylethanolamine (O-18:1_18:2) | SLM:000070214 |
| PE O-18:1;0/20:3;0 | PE O-18:1/20:3 | Phosphatidylethanolamine (O-18:1_20:3) | SLM:000070221 |
| PE O-18:1;0/20:4;0 | PE O-18:1/20:4 | Phosphatidylethanolamine (O-18:1_20:4) | SLM:000070222 |
| PE O-18:1;0/20:5;0 | PE O-18:1/20:5 | Phosphatidylethanolamine (O-18:1_20:5) | SLM:000070223 |
| PE O-18:1;0/22:4;0 | PE O-18:1/22:4 | Phosphatidylethanolamine (O-18:1_22:4) | SLM:000070229 |
| PE O-18:1;0/22:6;0 | PE O-18:1/22:6 | Phosphatidylethanolamine (O-18:1_22:6) | SLM:000070231 |
| PE O-18:2;0/16:0;0 | PE O-18:2/16:0 | Phosphatidylethanolamine (O-18:2_16:0) | SLM:000070273 |
| PE O-18:2;0/18:0;0 | PE O-18:2/18:0 | Phosphatidylethanolamine (O-18:2_18:0) | SLM:000070277 |
| PE O-18:2;0/18:1;0 | PE O-18:2/18:1 | Phosphatidylethanolamine (O-18:2_18:1) | SLM:000070278 |
| PE O-18:2;0/18:2;0 | PE O-18:2/18:2 | Phosphatidylethanolamine (O-18:2_18:2) | SLM:000070279 |
| PE O-18:2;0/20:3;0 | PE O-18:2/20:3 | Phosphatidylethanolamine (O-18:2_20:3) | SLM:000070286 |
| PE O-18:2;0/20:4;0 | PE O-18:2/20:4 | Phosphatidylethanolamine (O-18:2_20:4) | SLM:000070287 |

| | | | |
|---|---|---|---|
| PE O-18:2;0/22:5;0 | PE O-18:2/22:5 | Phosphatidylethanolamine (O-18:2_22:5) | SLM:000070295 |
| PI 16:0;0_18:1;0 | PI 16:0_18:1 | Phosphatidylinositol (16:0_18:1) | SLM:000073801 |
| PI 16:0;0_18:2;0 | PI 16:0_18:2 | Phosphatidylinositol (16:0_18:2) | SLM:000073802 |
| PI 16:0;0_20:3;0 | PI 16:0_20:3 | Phosphatidylinositol (16:0_20:3) | SLM:000073809 |
| PI 16:0;0_20:4;0 | PI 16:0_20:4 | Phosphatidylinositol (16:0_20:4) | SLM:000073810 |
| PI 16:1;0_18:0;0 | PI 16:1_18:0 | Phosphatidylinositol (16:1_18:0) | SLM:000073853 |
| PI 18:0;0_18:1;0 | PI 18:0_18:1 | Phosphatidylinositol (18:0_18:1) | SLM:000074007 |
| PI 18:0;0_18:2;0 | PI 18:0_18:2 | Phosphatidylinositol (18:0_18:2) | SLM:000074008 |
| PI 18:0;0_20:3;0 | PI 18:0_20:3 | Phosphatidylinositol (18:0_20:3) | SLM:000074015 |
| PI 18:0;0_20:4;0 | PI 18:0_20:4 | Phosphatidylinositol (18:0_20:4) | SLM:000074016 |
| PI 18:0;0_22:5;0 | PI 18:0_22:5 | Phosphatidylinositol (18:0_22:5) | SLM:000074024 |
| PI 18:1;0_18:1;0 | PI 18:1_18:1 | Phosphatidylinositol (18:1/18:1) | SLM:000105305 |
| PI 18:1;0_18:2;0 | PI 18:1_18:2 | Phosphatidylinositol (18:1_18:2) | SLM:000074057 |
| PI 18:1;0_20:3;0 | PI 18:1_20:3 | Phosphatidylinositol (18:1_20:3) | SLM:000074064 |
| PI 18:1;0_20:4;0 | PI 18:1_20:4 | Phosphatidylinositol (18:1_20:4) | SLM:000074065 |
| PI 18:2;0_18:2;0 | PI 18:2_18:2 | Phosphatidylinositol (18:2/18:2) | SLM:000105371 |
| SM 32:1;2 | SM 32:1;O2 | Sphingomyelin (d32:1) | SLM:000390695 |
| SM 32:2;2 | SM 32:2;O2 | Sphingomyelin (d32:2) | SLM:000390694 |
| SM 34:0;2 | SM 34:0;O2 | Sphingomyelin (d34:0) | SLM:000390716 |
| SM 34:1;2 | SM 34:1;O2 | Sphingomyelin (d34:1) | SLM:000390714 |
| SM 34:1;3 | SM 34:1;O3 | Sphingomyelin (t34:1) | SLM:000390728 |
| SM 34:2;2 | SM 34:2;O2 | Sphingomyelin (d34:2) | SLM:000390712 |
| SM 36:1;2 | SM 36:1;O2 | Sphingomyelin (d36:1) | SLM:000390739 |
| SM 36:2;2 | SM 36:2;O2 | Sphingomyelin (d36:2) | SLM:000390737 |
| SM 38:1;2 | SM 38:1;O2 | Sphingomyelin (d38:1) | SLM:000390767 |
| SM 38:2;2 | SM 38:2;O2 | Sphingomyelin (d38:2) | SLM:000390765 |
| SM 40:1;2 | SM 40:1;O2 | Sphingomyelin (d40:1) | SLM:000390797 |
| SM 40:2;2 | SM 40:2;O2 | Sphingomyelin (d40:2) | SLM:000390795 |
| SM 42:1;2 | SM 42:1;O2 | Sphingomyelin (d42:1) | SLM:000390824 |
| SM 42:2;2 | SM 42:2;O2 | Sphingomyelin (d42:2) | SLM:000390823 |
| TAG 44:1;0 | TG 44:1 | Triacylglycerol (44:1) | SLM:000308232 |

| | | | |
|---|---|---|---|
| TAG 46:0;0 | TG 46:0 | Triacylglycerol (46:0) | SLM:000308243 |
| TAG 46:1;0 | TG 46:1 | Triacylglycerol (46:1) | SLM:000308244 |
| TAG 46:2;0 | TG 46:2 | Triacylglycerol (46:2) | SLM:000308245 |
| TAG 48:0;0 | TG 48:0 | Triacylglycerol (48:0) | SLM:000308257 |
| TAG 48:1;0 | TG 48:1 | Triacylglycerol (48:1) | SLM:000308258 |
| TAG 48:2;0 | TG 48:2 | Triacylglycerol (48:2) | SLM:000308259 |
| TAG 48:3;0 | TG 48:3 | Triacylglycerol (48:3) | SLM:000308260 |
| TAG 49:1;0 | TG 49:1 | Triacylglycerol (49:1) | SLM:000308267 |
| TAG 49:2;0 | TG 49:2 | Triacylglycerol (49:2) | SLM:000308268 |
| TAG 49:3;0 | TG 49:3 | Triacylglycerol (49:3) | SLM:000308269 |
| TAG 50:1;0 | TG 50:1 | Triacylglycerol (50:1) | SLM:000308276 |
| TAG 50:2;0 | TG 50:2 | Triacylglycerol (50:2) | SLM:000308277 |
| TAG 50:3;0 | TG 50:3 | Triacylglycerol (50:3) | SLM:000308278 |
| TAG 50:4;0 | TG 50:4 | Triacylglycerol (50:4) | SLM:000308279 |
| TAG 50:5;0 | TG 50:5 | Triacylglycerol (50:5) | SLM:000308280 |
| TAG 51:1;0 | TG 51:1 | Triacylglycerol (51:1) | SLM:000308286 |
| TAG 51:2;0 | TG 51:2 | Triacylglycerol (51:2) | SLM:000308287 |
| TAG 51:3;0 | TG 51:3 | Triacylglycerol (51:3) | SLM:000308288 |
| TAG 51:4;0 | TG 51:4 | Triacylglycerol (51:4) | SLM:000308289 |
| TAG 52:2;0 | TG 52:2 | Triacylglycerol (52:2) | SLM:000308298 |
| TAG 52:3;0 | TG 52:3 | Triacylglycerol (52:3) | SLM:000308299 |
| TAG 52:4;0 | TG 52:4 | Triacylglycerol (52:4) | SLM:000308300 |
| TAG 52:5;0 | TG 52:5 | Triacylglycerol (52:5) | SLM:000308301 |
| TAG 52:6;0 | TG 52:6 | Triacylglycerol (52:6) | SLM:000308302 |
| TAG 53:2;0 | TG 53:2 | Triacylglycerol (53:2) | SLM:000308309 |
| TAG 53:3;0 | TG 53:3 | Triacylglycerol (53:3) | SLM:000308310 |
| TAG 53:4;0 | TG 53:4 | Triacylglycerol (53:4) | SLM:000308311 |
| TAG 53:5;0 | TG 53:5 | Triacylglycerol (53:5) | SLM:000308312 |
| TAG 54:3;0 | TG 54:3 | Triacylglycerol (54:3) | SLM:000308323 |
| TAG 54:4;0 | TG 54:4 | Triacylglycerol (54:4) | SLM:000308324 |
| TAG 54:5;0 | TG 54:5 | Triacylglycerol (54:5) | SLM:000308325 |

| | | | |
|---|---|---|---|
| TAG 54:6;0 | TG 54:6 | Triacylglycerol (54:6) | SLM:000308326 |
| TAG 54:7;0 | TG 54:7 | Triacylglycerol (54:7) | SLM:000308327 |
| TAG 56:3;0 | TG 56:3 | Triacylglycerol (56:3) | SLM:000308349 |
| TAG 56:5;0 | TG 56:5 | Triacylglycerol (56:5) | SLM:000308351 |
| TAG 56:6;0 | TG 56:6 | Triacylglycerol (56:6) | SLM:000308352 |
| TAG 56:7;0 | TG 56:7 | Triacylglycerol (56:7) | SLM:000308353 |
| TAG 56:8;0 | TG 56:8 | Triacylglycerol (56:8) | SLM:000308354 |
| TAG 56:9;0 | TG 56:9 | Triacylglycerol (56:9) | SLM:000308355 |
| TAG 58:7;0 | TG 58:7 | Triacylglycerol (58:7) | SLM:000308381 |
| TAG 58:8;0 | TG 58:8 | Triacylglycerol (58:8) | SLM:000308382 |
| TAG 58:9;0 | TG 58:9 | Triacylglycerol (58:9) | SLM:000308383 |

**Table S5.** Lipid species showing alterations within or between eTRE and lTRE interventions.

| Lipid species | Before eTRE | After eTRE | Change eTRE after – before (95% CI) | P[a)] | P BH[b)] | Before lTRE | After lTRE | Change lTRE After – before (95% CI) | P[a)] | P BH[b)] | Difference between lTRE vs. eTRE (95% CI)[a] | P[a)] | P BH[b)] |
|---|---|---|---|---|---|---|---|---|---|---|---|---|---|
| **PC 18:0;0_20:3;0** | 35.6 | 27.5 | -8.1 (4.6 to 11) | **0.000002** | **0.0006** | 35.9 | 32.9 | -3.1 (0.86 to 6) | **0.015** | 0.85 | 4.8 (-9.1 to -0.49) | **0.028** | 0.24 |
| **LPE 20:2;0** | 0.53 | 0.39 | -0.13 (0.069 to 0.17) | **0.0000057** | **0.00076** | 0.47 | 0.5 | 0.034 (-0.077 to 0.043) | 0.73 | 1 | 0.2 (-0.29 to -0.066) | **0.001** | 0.17 |
| **PC 18:2;0_20:3;0** | 4 | 3.1 | -0.96 (0.56 to 1.3) | **0.0000076** | **0.00076** | 3.9 | 3.6 | -0.3 (-0.14 to 0.72) | 0.19 | 1 | 0.82 (-1.4 to -0.19) | **0.0065** | 0.17 |
| **PC 18:0;0_18:2;0** | 234.9 | 191.4 | -43 (23 to 60) | **0.000024** | **0.0014** | 234 | 225.9 | -8.1 (-8 to 26) | 0.43 | 1 | 35 (-65 to 1.1) | 0.067 | 0.33 |
| **PE 18:0;0_20:3;0** | 2.5 | 2.1 | -0.41 (0.26 to 0.54) | **0.000024** | **0.0014** | 2.4 | 2.3 | -0.16 (-0.11 to 0.46) | 0.3 | 1 | 0.24 (-0.56 to 0.11) | 0.16 | 0.38 |
| **LPE 18:1;0** | 0.72 | 0.52 | -0.2 (0.068 to 0.26) | **0.000028** | **0.0014** | 0.67 | 0.7 | 0.032 (-0.13 to 0.078) | 0.67 | 1 | 0.25 (-0.38 to -0.093) | **0.0022** | 0.17 |
| **PE O-18:1;0/20:5;0** | 0.72 | 0.53 | -0.19 (0.14 to 0.32) | **0.000044** | **0.0019** | 0.71 | 0.74 | 0.038 (-0.18 to 0.11) | 0.79 | 1 | 0.34 (-0.55 to -0.099) | **0.0074** | 0.17 |
| **PC 16:0;0_18:3;0** | 11.2 | 7.3 | -4 (1.1 to 3.3) | **0.000063** | **0.0023** | 9.5 | 9.9 | 0.39 (-0.95 to 1.5) | 0.54 | 1 | 4.3 (-4.4 to -0.019) | 0.05 | 0.29 |
| **PE 18:0;0_18:3;0** | 0.3 | 0.22 | -0.08 (0.036 to 0.1) | **0.000068** | **0.0023** | 0.29 | 0.28 | -0.016 (-0.005 to 0.056) | 0.086 | 1 | 0.085 (-0.096 to 0.011) | 0.17 | 0.39 |
| **PC 18:0;0_22:5;0** | 7.2 | 5.8 | -1.4 (0.69 to 2) | **0.000089** | **0.0024** | 7.4 | 6.8 | -0.57 (0.12 to 1.2) | **0.016** | 0.85 | 0.83 (-1.7 to 0.082) | 0.088 | 0.33 |

| | | | | | | | | | | | | | |
|---|---|---|---|---|---|---|---|---|---|---|---|---|---|
| **CE 18:3;0** | 121.4 | 92.3 | -29 (12 to 40) | **0.000089** | **0.0024** | 122 | 117.2 | -4.8 (-8.6 to 17) | 0.52 | 1 | 24 (-50 to 3.9) | 0.12 | 0.35 |
| **PC 18:0;0_18:1;0** | 39 | 30.8 | -8.2 (3.3 to 12) | **0.00011** | **0.0028** | 39.4 | 38.3 | -1.2 (-2.1 to 5.1) | 0.38 | 1 | 7.1 (-11 to -0.45) | **0.04** | 0.25 |
| **PC 18:0;0_20:2;0** | 2.6 | 2.1 | -0.49 (0.29 to 0.81) | **0.00012** | **0.0028** | 2.8 | 2.4 | -0.33 (0.053 to 0.51) | **0.02** | 0.85 | 0.28 (-0.74 to 0.089) | 0.12 | 0.35 |
| **PC 18:0;0_18:3;0** | 4.5 | 2.8 | -1.7 (0.64 to 2.5) | **0.00013** | **0.0028** | 3.8 | 3.8 | 0.039 (-0.42 to 0.76) | 0.51 | 1 | 1.4 (-2.3 to 0.022) | 0.076 | 0.33 |
| **PC 16:0;0_18:0;0** | 27 | 21.5 | -5.5 (2 to 6) | **0.00017** | **0.0032** | 25 | 24.1 | -0.89 (-1.1 to 2.5) | 0.49 | 1 | 4.6 (-6.5 to -0.71) | **0.023** | 0.23 |
| **PE O-18:1;0/18:1;0** | 0.63 | 0.5 | -0.13 (0.055 to 0.23) | **0.00017** | **0.0032** | 0.64 | 0.59 | -0.046 (-0.028 to 0.15) | 0.18 | 1 | 0.11 (-0.22 to 0.022) | 0.15 | 0.37 |
| **SM 42:1;2** | 18.1 | 16.2 | -1.9 (0.82 to 3) | **0.00023** | **0.0037** | 18 | 18.3 | 0.36 (-1.7 to 1.4) | 0.92 | 1 | 2.3 (-4 to 0.31) | 0.084 | 0.33 |
| **PC 18:1;0_20:2;0** | 1.5 | 1.2 | -0.28 (0.15 to 0.42) | **0.00023** | **0.0037** | 1.5 | 1.4 | -0.12 (-0.093 to 0.23) | 0.42 | 1 | 0.2 (-0.48 to 0.075) | 0.11 | 0.35 |
| **PE O-18:1;0/18:2;0** | 2.6 | 2.0 | -0.56 (0.28 to 0.82) | **0.00023** | **0.0037** | 2.5 | 2.3 | -0.2 (-0.073 to 0.45) | 0.18 | 1 | 0.38 (-0.88 to 0.097) | 0.12 | 0.35 |
| **PE 18:0;0_20:2;0** | 3.5 | 2.9 | -0.66 (0.33 to 1.1) | **0.00026** | **0.0038** | 3.3 | 3.2 | -0.063 (-0.35 to 0.33) | 0.94 | 1 | 0.72 (-1.5 to -0.039) | **0.035** | 0.24 |
| **PC 18:2;0_18:2;0** | 28.9 | 21.7 | -7.2 (3.5 to 10) | **0.00028** | **0.004** | 28.7 | 27.6 | -1.1 (-2.6 to 5) | 0.48 | 1 | 6.1 (-12 to 1.3) | 0.12 | 0.36 |

| | | | | | | | | | | | | | |
|---|---|---|---|---|---|---|---|---|---|---|---|---|---|
| **PC 18:2;0_18:3;0** | 2.6 | 1.6 | -0.97 (0.43 to 1.9) | **0.00033** | **0.0045** | 2.2 | 2.1 | -0.042 (-0.54 to 0.42) | 0.75 | 1 | 1.5 (-2.4 to -0.25) | **0.011** | 0.17 |
| **Cer 40:1;2** | 1.5 | 1.3 | -0.2 (0.084 to 0.29) | **0.00038** | **0.005** | 1.5 | 1.4 | -0.063 (-0.031 to 0.18) | 0.16 | 1 | 0.14 (-0.28 to 0.08) | 0.27 | 0.49 |
| **PE O-18:1;0/20:3;0** | 0.69 | 0.54 | -0.15 (0.07 to 0.26) | **0.00042** | **0.0053** | 0.71 | 0.59 | -0.12 (0.006 to 0.19) | **0.036** | 0.97 | 0.098 (-0.26 to 0.035) | 0.14 | 0.37 |
| **PC 18:2;0_20:4;0** | 11.6 | 10 | -1.5 (0.46 to 2.1) | **0.00067** | **0.0077** | 11.5 | 11.9 | 0.43 (-1.3 to 0.69) | 0.56 | 1 | 2 (-3.2 to 0.11) | 0.088 | 0.33 |
| **PC 16:1;0_18:0;0** | 2.4 | 1.9 | -0.46 (0.16 to 0.62) | **0.00067** | **0.0077** | 2.3 | 2.3 | -0.035 (-0.16 to 0.32) | 0.49 | 1 | 0.42 (-0.79 to 0.13) | 0.16 | 0.37 |
| **SM 40:1;2** | 31.3 | 27.9 | -3.4 (1.6 to 4.9) | **0.0008** | **0.0087** | 30.8 | 31.2 | 0.44 (-2.5 to 2.2) | 0.98 | 1 | 3.9 (-6.8 to 0.17) | 0.055 | 0.31 |
| **PE O-16:1;0/18:2;0** | 1.6 | 1.3 | -0.32 (0.15 to 0.57) | **0.00081** | **0.0087** | 1.5 | 1.4 | -0.095 (-0.11 to 0.29) | 0.26 | 1 | 0.32 (-0.72 to 0.073) | 0.096 | 0.34 |
| **PE O-18:2;0/18:1;0** | 0.59 | 0.46 | -0.12 (0.05 to 0.19) | **0.00087** | **0.009** | 0.58 | 0.52 | -0.065 (0.0069 to 0.12) | **0.025** | 0.92 | 0.062 (-0.14 to 0.049) | 0.31 | 0.52 |
| **PC 18:1;0_18:3;0** | 3.1 | 2 | -1.1 (0.36 to 1.4) | **0.00094** | **0.0092** | 2.7 | 2.8 | 0.12 (-0.49 to 0.39) | 1 | 1 | 1.3 (-1.8 to -0.26) | **0.0034** | 0.17 |
| **PC O-18:1;0/20:3;0** | 1.2 | 0.98 | -0.21 (0.079 to 0.34) | **0.00095** | **0.0092** | 1.2 | 1.1 | -0.13 (-0.049 to 0.29) | 0.12 | 1 | 0.095 (-0.27 to 0.036) | 0.11 | 0.35 |
| **PE 16:0;0_20:3;0** | 0.47 | 0.39 | -0.079 (0.042 to 0.13) | **0.001** | **0.0094** | 0.45 | 0.46 | 0.01 (-0.048 to 0.062) | 0.8 | 1 | 0.13 (-0.24 to -0.025) | **0.011** | 0.17 |
| **PI 18:1;0_18:1;0** | 0.97 | 0.65 | -0.31 (0.11 to 0.45) | **0.001** | **0.0094** | 0.88 | 0.85 | -0.033 (-0.092 to 0.17) | 0.66 | 1 | 0.29 (-0.53 to -0.065) | **0.016** | 0.22 |

| | | | | | | | | | | | | | |
|---|---|---|---|---|---|---|---|---|---|---|---|---|---|
| **PC O-16:2;0/18:1;0** | 0.37 | 0.3 | -0.068 (0.034 to 0.12) | **0.0013** | **0.011** | 0.33 | 0.35 | 0.019 (-0.056 to 0.027) | 0.56 | 1 | 0.085 (-0.16 to -0.026) | **0.011** | 0.17 |
| **LPE 20:0;0** | 0.5 | 0.41 | -0.088 (0.02 to 0.11) | **0.0013** | **0.011** | 0.5 | 0.48 | -0.021 (-0.025 to 0.07) | 0.42 | 1 | 0.089 (-0.16 to -0.0095) | **0.022** | 0.22 |
| **PE O-16:1;0/18:1;0** | 0.53 | 0.44 | -0.091 (0.042 to 0.17) | **0.0013** | **0.011** | 0.55 | 0.51 | -0.042 (-0.014 to 0.1) | 0.11 | 1 | 0.071 (-0.15 to 0.042) | 0.35 | 0.55 |
| **PC O-18:2;0/20:3;0** | 0.59 | 0.46 | -0.13 (0.048 to 0.2) | **0.0015** | **0.011** | 0.58 | 0.49 | -0.084 (-0.061 to 0.13) | 0.26 | 1 | 0.086 (-0.16 to 0.0095) | 0.15 | 0.37 |
| **PC 18:1;0_18:2;0** | 64.4 | 54.2 | -10 (4 to 16) | **0.0015** | **0.011** | 63.3 | 62.4 | -0.9 (-4.3 to 5.4) | 0.87 | 1 | 9.2 (-20 to 2.1) | 0.15 | 0.37 |
| **PE O-16:1;0/22:5;0** | 2 | 1.6 | -0.35 (0.13 to 0.46) | **0.0015** | **0.011** | 2 | 1.9 | -0.11 (-0.099 to 0.32) | 0.37 | 1 | 0.23 (-0.54 to 0.17) | 0.37 | 0.57 |
| **PC 18:0;0_20:5;0** | 10.3 | 8 | -2.3 (0.99 to 3.5) | **0.0016** | **0.012** | 9.7 | 10.4 | 0.64 (-1.9 to 1) | 0.72 | 1 | 2.8 (-5.3 to -0.5) | **0.021** | 0.22 |
| **PC O-16:0;0/18:3;0** | 0.31 | 0.23 | -0.077 (0.021 to 0.16) | **0.0017** | **0.012** | 0.27 | 0.28 | 0.0011 (-0.055 to 0.049) | 0.99 | 1 | 0.21 (-0.37 to -0.042) | **0.0063** | 0.17 |
| **SM 38:1;2** | 17.8 | 16.1 | -1.7 (0.61 to 2.6) | **0.0017** | **0.012** | 17.3 | 17.7 | 0.41 (-1.5 to 0.97) | 0.65 | 1 | 2.1 (-3.6 to -0.16) | **0.029** | 0.24 |
| **Cer 42:1;2** | 3.8 | 3.3 | -0.49 (0.2 to 0.76) | **0.0017** | **0.012** | 3.7 | 3.7 | -0.077 (-0.097 to 0.43) | 0.14 | 1 | 0.42 (-0.75 to 0.17) | 0.27 | 0.49 |
| **PC 17:0;0_18:2;0** | 16.3 | 14.5 | -1.8 (0.48 to 2.9) | **0.0019** | **0.012** | 16.4 | 16.7 | 0.3 (-2.3 to 0.96) | 0.41 | 1 | 2.1 (-4.5 to 0.45) | 0.11 | 0.35 |
| **PE O-18:2;0/16:0;0** | 0.21 | 0.18 | -0.03 (0.015 to 0.06) | **0.0019** | **0.012** | 0.21 | 0.2 | -0.01 (-0.014 to 0.044) | 0.32 | 1 | 0.025 (-0.056 to 0.02) | 0.37 | 0.57 |
| **LPE 18:2;0** | 1.2 | 0.91 | -0.29 (0.084 to 0.44) | **0.002** | **0.013** | 1.2 | 1.1 | -0.068 (-0.12 to 0.21) | 0.52 | 1 | 0.23 (-0.45 to 0.016) | 0.07 | 0.33 |

| | | | | | | | | | | | | | |
|---|---|---|---|---|---|---|---|---|---|---|---|---|---|
| **PC O-18:0;0/18:3;0** | 0.17 | 0.11 | -0.055 (0.015 to 0.074) | **0.002** | **0.013** | 0.14 | 0.14 | 0.0042 (-0.023 to 0.029) | 0.75 | 1 | 0.067 (-0.11 to 0.0052) | 0.074 | 0.33 |
| **PC 17:0;0_18:3;0** | 0.55 | 0.4 | -0.14 (0.025 to 0.16) | **0.002** | **0.013** | 0.79 | 0.95 | 0.16 (-0.066 to 0.069) | 0.96 | 1 | 0.088 (-0.19 to 0.081) | 0.46 | 0.65 |
| **PC 16:0;0_20:1;0** | 1.8 | 1.6 | -0.18 (0.094 to 0.34) | **0.0022** | **0.013** | 1.8 | 1.8 | 0.014 (-0.16 to 0.14) | 0.92 | 1 | 0.2 (-0.4 to -0.054) | **0.02** | 0.22 |
| **PE 18:1;0_20:3;0** | 0.83 | 0.68 | -0.15 (0.072 to 0.22) | **0.0023** | **0.014** | 0.81 | 0.82 | 0.0051 (-0.11 to 0.11) | 0.94 | 1 | 0.13 (-0.3 to 0.035) | 0.13 | 0.37 |
| **PC O-18:2;0/18:2;0** | 2.7 | 2.2 | -0.45 (0.15 to 0.63) | **0.0026** | **0.015** | 2.6 | 2.6 | -0.023 (-0.3 to 0.37) | 0.66 | 1 | 0.44 (-0.85 to 0.12) | 0.1 | 0.35 |
| **PC O-17:2;0/17:0;0** | 0.16 | 0.13 | -0.024 (0.0088 to 0.039) | **0.0029** | **0.017** | 0.16 | 0.16 | -0.0056 (-0.01 to 0.026) | 0.32 | 1 | 0.012 (-0.037 to 0.015) | 0.37 | 0.57 |
| **LPC 18:2;0** | 17.2 | 13.1 | -4.1 (0.84 to 4.6) | **0.003** | **0.017** | 16.5 | 16.9 | 0.48 (-2.5 to 2) | 0.93 | 1 | 4.8 (-7.3 to -1.1) | **0.015** | 0.22 |
| **PC O-16:1;0/18:2;0** | 6.9 | 5.8 | -1.1 (0.36 to 1.7) | **0.003** | **0.017** | 6.5 | 6.5 | -0.02 (-0.63 to 0.7) | 0.88 | 1 | 1.2 (-2.2 to -0.033) | **0.039** | 0.25 |
| **PC 18:2;0_20:0;0** | 0.84 | 0.68 | -0.16 (0.06 to 0.29) | **0.0032** | **0.017** | 0.76 | 0.79 | 0.032 (-0.079 to 0.1) | 0.6 | 1 | 0.26 (-0.4 to -0.081) | **0.0043** | 0.17 |
| **SM 32:1;2** | 11.9 | 10.8 | -1.1 (0.34 to 1.8) | **0.0032** | **0.017** | 11.8 | 12 | 0.18 (-1.1 to 0.7) | 0.85 | 1 | 1.4 (-2.5 to 0.076) | 0.067 | 0.33 |
| **CE 20:3;0** | 45.2 | 38.4 | -6.7 (2.3 to 11) | **0.0035** | **0.018** | 46.5 | 44.9 | -1.6 (-2 to 6.4) | 0.26 | 1 | 5.1 (-12 to 1.3) | 0.15 | 0.37 |
| **PC 18:2;0_20:2;0** | 2.2 | 1.8 | -0.32 (0.15 to 0.58) | **0.0037** | **0.019** | 2.2 | 2.1 | -0.16 (-0.11 to 0.43) | 0.21 | 1 | 0.23 (-0.74 to 0.24) | 0.28 | 0.49 |

| | | | | | | | | | | | | | |
|---|---|---|---|---|---|---|---|---|---|---|---|---|---|
| **PC 14:0;0_20:4;0** | 2.8 | 2.5 | -0.32 (0.16 to 0.91) | **0.0039** | **0.02** | 2.8 | 2.9 | 0.099 (-0.33 to 0.27) | 0.86 | 1 | 0.71 (-1.2 to -0.27) | **0.006** | 0.17 |
| **PC 16:0;0_20:2;0** | 8.5 | 7.5 | -1 (0.3 to 1.8) | **0.0043** | **0.022** | 8.7 | 8.2 | -0.55 (-0.11 to 1.4) | 0.066 | 1 | 0.55 (-1.6 to 0.63) | 0.53 | 0.69 |
| **PI 18:0;0_18:1;0** | 2.2 | 1.7 | -0.41 (0.14 to 0.63) | **0.0047** | **0.023** | 2.3 | 2.1 | -0.15 (-0.11 to 0.42) | 0.31 | 1 | 0.27 (-0.61 to 0.09) | 0.18 | 0.4 |
| **PC O-16:2;0/16:0;0** | 2.8 | 2.2 | -0.54 (0.13 to 0.9) | **0.005** | **0.024** | 2.6 | 2.7 | 0.074 (-0.27 to 0.33) | 0.81 | 1 | 0.69 (-1.2 to -0.094) | **0.025** | 0.24 |
| **PI 16:0;0_18:1;0** | 1 | 0.75 | -0.3 (0.096 to 0.45) | **0.005** | **0.024** | 1.1 | 1.1 | 0.021 (-0.25 to 0.19) | 0.82 | 1 | 0.34 (-0.59 to -0.07) | **0.026** | 0.24 |
| **PI 18:0;0_20:3;0** | 2.6 | 2.2 | -0.32 (0.1 to 0.52) | **0.005** | **0.024** | 2.8 | 2.6 | -0.21 (-0.048 to 0.53) | 0.09 | 1 | 0.1 (-0.48 to 0.28) | 0.53 | 0.69 |
| **CE 17:1;0** | 12.8 | 11 | -1.8 (0.53 to 3) | **0.0054** | **0.025** | 13.1 | 13.1 | -0.024 (-1.7 to 1.5) | 0.82 | 1 | 1.8 (-4.1 to 0.55) | 0.12 | 0.35 |
| **PE O-18:2;0/22:5;0** | 1.8 | 1.5 | -0.34 (0.099 to 0.62) | **0.0056** | **0.025** | 1.9 | 1.6 | -0.25 (0.028 to 0.53) | **0.033** | 0.97 | 0.1 (-0.48 to 0.37) | 0.94 | 0.97 |
| **PC 14:0;0_18:1;0** | 4.4 | 3.6 | -0.86 (0.27 to 1.4) | **0.0058** | **0.025** | 4.5 | 4.5 | 0.044 (-0.52 to 0.45) | 0.79 | 1 | 0.9 (-1.5 to -0.18) | **0.011** | 0.17 |
| **LPC 18:1;0** | 13.3 | 10.8 | -2.5 (0.4 to 3) | **0.0058** | **0.025** | 13 | 13.3 | 0.24 (-1.6 to 1.5) | 0.96 | 1 | 2.9 (-4.6 to -0.48) | **0.011** | 0.17 |
| **PC O-18:0;0/20:5;0** | 0.86 | 0.75 | -0.11 (0.049 to 0.24) | **0.0059** | **0.026** | 0.87 | 0.93 | 0.056 (-0.16 to 0.078) | 0.45 | 1 | 0.18 (-0.35 to -0.053) | **0.007** | 0.17 |
| **PC 18:1;0_20:5;0** | 6.5 | 5.4 | -1.1 (0.35 to 2) | **0.0061** | **0.026** | 6.5 | 6.6 | 0.099 (-0.99 to 0.74) | 0.9 | 1 | 1.2 (-3 to 0.44) | 0.14 | 0.37 |

| | | | | | | | | | | | | | |
|---|---|---|---|---|---|---|---|---|---|---|---|---|---|
| **PC O-16:2;0/18:0;0** | 0.32 | 0.25 | -0.069 (0.015 to 0.1) | **0.0062** | **0.026** | 0.32 | 0.31 | -0.019 (-0.032 to 0.062) | 0.57 | 1 | 0.058 (-0.11 to -0.01) | **0.02** | 0.22 |
| **PC 14:0;0_20:3;0** | 0.82 | 0.7 | -0.12 (0.046 to 0.33) | **0.0066** | **0.027** | 0.79 | 0.81 | 0.015 (-0.12 to 0.2) | 0.7 | 1 | 0.17 (-0.37 to 0.026) | 0.098 | 0.34 |
| **PC O-17:0;0/17:1;0** | 0.44 | 0.38 | -0.069 (0.017 to 0.11) | **0.0066** | **0.027** | 0.46 | 0.45 | -0.014 (-0.049 to 0.068) | 0.84 | 1 | 0.063 (-0.12 to 0.019) | 0.18 | 0.39 |
| **CE 14:0;0** | 26.7 | 22 | -4.6 (1.6 to 9.2) | **0.008** | **0.031** | 27 | 26.8 | -0.12 (-2.8 to 3.3) | 0.98 | 1 | 5.8 (-11 to -1) | **0.018** | 0.22 |
| **PC O-16:0;0/18:1;0** | 2.7 | 2.4 | -0.28 (0.068 to 0.44) | **0.0081** | **0.031** | 2.7 | 2.7 | 0.068 (-0.33 to 0.25) | 0.98 | 1 | 0.38 (-0.61 to 0.025) | 0.07 | 0.33 |
| **PC O-16:0;0/16:0;0** | 7.4 | 6.7 | -0.71 (0.2 to 1.2) | **0.0081** | **0.031** | 7.8 | 7.6 | -0.17 (-0.37 to 0.96) | 0.42 | 1 | 0.65 (-1.5 to 0.28) | 0.23 | 0.45 |
| **PC 18:1;0_20:3;0** | 11.9 | 10.6 | -1.3 (0.41 to 2.2) | **0.0081** | **0.031** | 12.3 | 11.9 | -0.47 (-0.34 to 1.5) | 0.18 | 1 | 0.83 (-2.1 to 0.63) | 0.3 | 0.51 |
| **CE 18:2;0** | 2741.2 | 2489.3 | -250 (74 to 440) | **0.0081** | **0.031** | 2756.7 | 2744.5 | -12 (-200 to 250) | 0.68 | 1 | 240 (-660 to 160) | 0.3 | 0.51 |
| **PC 16:0;0_20:3;0** | 66.6 | 59.1 | -7.5 (1.8 to 12) | **0.0081** | **0.031** | 69 | 63.9 | -5.2 (-0.34 to 12) | 0.069 | 1 | 2.4 (-10 to 6.7) | 0.7 | 0.79 |
| **PI 18:1;0_20:3;0** | 0.34 | 0.28 | -0.064 (0.017 to 0.12) | **0.0083** | **0.031** | 0.32 | 0.31 | -0.012 (-0.035 to 0.073) | 0.68 | 1 | 0.076 (-0.14 to 0.01) | 0.083 | 0.33 |
| **PC O-16:1;0/20:3;0** | 1.5 | 1.2 | -0.28 (0.074 to 0.44) | **0.0087** | **0.032** | 1.5 | 1.5 | -0.032 (-0.18 to 0.19) | 0.96 | 1 | 0.25 (-0.57 to 0.051) | 0.11 | 0.35 |

| | | | | | | | | | | | | | |
|---|---|---|---|---|---|---|---|---|---|---|---|---|---|
| **CE 16:1;0** | 150.1 | 129.8 | -20 (5.7 to 30) | **0.0087** | **0.032** | 156 | 156.1 | 0.072 (-11 to 16) | 0.76 | 1 | 21 (-43 to 6) | 0.14 | 0.37 |
| **PC O-16:0;0/18:0;0** | 0.12 | 0.11 | -0.013 (0.0038 to 0.051) | **0.009** | **0.033** | 0.14 | 0.11 | -0.026 (-0.002 to 0.019) | 0.09 | 1 | 0.0079 (-0.037 to 0.016) | 0.49 | 0.67 |
| **PE O-16:1;0/20:3;0** | 0.39 | 0.31 | -0.084 (0.02 to 0.15) | **0.0092** | **0.033** | 0.41 | 0.43 | 0.012 (-0.047 to 0.068) | 0.56 | 1 | 0.088 (-0.2 to 0.0085) | 0.096 | 0.34 |
| **LPC 18:0;0** | 18.4 | 15.5 | -2.9 (0.44 to 3.9) | **0.0093** | **0.033** | 19.3 | 18.4 | -0.98 (-1.1 to 3) | 0.37 | 1 | 2.2 (-4.4 to 0.075) | 0.058 | 0.32 |
| **PC O-16:0;0/20:3;0** | 1.4 | 1.2 | -0.2 (0.064 to 0.45) | **0.0095** | **0.033** | 1.5 | 1.4 | -0.11 (-0.026 to 0.29) | 0.079 | 1 | 0.18 (-0.45 to -0.021) | **0.034** | 0.24 |
| **PC 17:1;0_18:2;0** | 3 | 2.7 | -0.25 (0.073 to 0.88) | **0.0096** | **0.033** | 3.1 | 2.8 | -0.23 (-0.17 to 0.51) | 0.44 | 1 | 0.56 (-0.79 to 0.072) | 0.15 | 0.37 |
| **PC 17:0;0_20:5;0** | 1 | 0.68 | -0.35 (0.031 to 0.54) | **0.01** | **0.034** | 0.89 | 1.4 | 0.48 (-0.21 to 0.048) | 0.41 | 1 | 0.66 (-0.76 to -0.026) | **0.02** | 0.22 |
| **PE O-18:0;0/18:2;0** | 0.29 | 0.25 | -0.04 (0.013 to 0.094) | **0.01** | **0.035** | 0.29 | 0.27 | -0.015 (-0.015 to 0.055) | 0.23 | 1 | 0.035 (-0.089 to 0.019) | 0.27 | 0.49 |
| **PE 18:0;0_20:5;0** | 0.59 | 0.47 | -0.12 (0.011 to 0.2) | **0.011** | **0.035** | 0.58 | 0.61 | 0.024 (-0.066 to 0.1) | 0.79 | 1 | 0.052 (-0.18 to 0.076) | 0.24 | 0.47 |
| **PC O-16:1;0/22:5;0** | 1 | 0.87 | -0.13 (0.037 to 0.28) | **0.012** | **0.039** | 0.93 | 0.98 | 0.052 (-0.25 to 0.085) | 0.43 | 1 | 0.34 (-0.56 to -0.091) | **0.0096** | 0.17 |
| **PE 18:1;0_18:1;0** | 0.9 | 0.75 | -0.16 (0.028 to 0.26) | **0.012** | **0.039** | 0.88 | 0.94 | 0.056 (-0.18 to 0.068) | 0.39 | 1 | 0.22 (-0.35 to -0.018) | **0.028** | 0.24 |

| | | | | | | | | | | | | | |
|---|---|---|---|---|---|---|---|---|---|---|---|---|---|
| **PC 18:1;0_18:1;0** | 24.7 | 21 | -3.7 (0.67 to 6.1) | **0.014** | **0.044** | 23.6 | 24.4 | 0.78 (-2.7 to 1.2) | 0.47 | 1 | 4.5 (-8.1 to -0.17) | **0.036** | 0.24 |
| **PC 16:1;0_20:3;0** | 1.8 | 1.7 | -0.15 (0.044 to 0.39) | **0.014** | **0.044** | 1.9 | 1.8 | -0.063 (-0.059 to 0.3) | 0.25 | 1 | 0.17 (-0.51 to 0.19) | 0.34 | 0.54 |
| **PC O-16:1;0/18:0;0** | 0.44 | 0.39 | -0.05 (0.009 to 0.089) | **0.015** | **0.046** | 0.43 | 0.44 | 0.0094 (-0.048 to 0.047) | 0.72 | 1 | 0.062 (-0.12 to -0.0032) | **0.035** | 0.24 |
| **PI 16:1;0_18:0;0** | 1.1 | 0.98 | -0.098 (0.02 to 0.17) | **0.015** | **0.046** | 1.1 | 1.1 | -0.014 (-0.073 to 0.11) | 0.52 | 1 | 0.087 (-0.21 to 0.014) | 0.081 | 0.33 |
| **PC O-16:0;0/18:2;0** | 5.2 | 4.4 | -0.84 (0.15 to 1.4) | **0.015** | **0.046** | 5.3 | 5.2 | -0.094 (-0.6 to 0.77) | 0.68 | 1 | 0.78 (-1.9 to 0.24) | 0.12 | 0.35 |
| **PE O-18:2;0/18:2;0** | 1.3 | 1.1 | -0.25 (0.059 to 0.45) | **0.015** | **0.046** | 1.3 | 1.2 | -0.082 (-0.13 to 0.29) | 0.43 | 1 | 0.24 (-0.57 to 0.11) | 0.19 | 0.4 |
| **PI 16:0;0_20:4;0** | 2 | 1.7 | -0.27 (0.05 to 0.44) | **0.015** | **0.046** | 2 | 2 | 0.0083 (-0.19 to 0.23) | 0.82 | 1 | 0.29 (-0.52 to 0.1) | 0.21 | 0.43 |
| **PC 16:0;0_18:2;0** | 466 | 420 | -46 (12 to 70) | **0.015** | **0.046** | 470 | 461 | -8.9 (-25 to 45) | 0.61 | 1 | 38 (-100 to 43) | 0.64 | 0.77 |
| **PC 18:2;0_20:1;0** | 2 | 1.7 | -0.36 (0.084 to 0.67) | **0.016** | **0.048** | 1.8 | 1.9 | 0.11 (-0.34 to 0.11) | 0.28 | 1 | 0.47 (-0.93 to -0.034) | **0.032** | 0.24 |
| **CE 18:1;0** | 798 | 720 | -78 (19 to 130) | **0.016** | **0.048** | 813 | 833 | 20 (-78 to 57) | 0.78 | 1 | 97 (-200 to 28) | 0.15 | 0.37 |
| **PI 16:0;0_20:3;0** | 0.46 | 0.4 | -0.062 (0.015 to 0.11) | **0.017** | **0.049** | 0.47 | 0.45 | -0.023 (-0.014 to 0.065) | 0.18 | 1 | 0.053 (-0.13 to 0.02) | 0.16 | 0.38 |
| **TAG 48:0;0** | 6.8 | 4.4 | -2.4 (0.27 to 3.1) | **0.018** | 0.052 | 7 | 6.7 | -0.24 (-0.93 to 2.3) | 0.24 | 1 | 1.8 (-3.7 to 1.4) | 0.5 | 0.67 |

| | | | | | | | | | | | | | |
|---|---|---|---|---|---|---|---|---|---|---|---|---|---|
| **Cer 40:0;2** | 0.27 | 0.24 | -0.031 (0.0045 to 0.078) | **0.019** | 0.055 | 0.28 | 0.3 | 0.019 (-0.045 to 0.018) | 0.57 | 1 | 0.082 (-0.14 to -0.025) | **0.0052** | 0.17 |
| **PC 18:2;0_19:0;0** | 1.4 | 1.2 | -0.18 (0.023 to 0.37) | **0.019** | 0.055 | 1.3 | 1.4 | 0.069 (-0.31 to 0.071) | 0.23 | 1 | 0.43 (-0.76 to -0.15) | **0.0095** | 0.17 |
| **PC O-16:1;0/18:1;0** | 1.2 | 1 | -0.17 (0.023 to 0.24) | **0.02** | 0.055 | 1.2 | 1.2 | 0.0036 (-0.17 to 0.14) | 0.9 | 1 | 0.18 (-0.35 to 0.048) | 0.15 | 0.37 |
| **PE O-18:2;0/20:3;0** | 0.71 | 0.53 | -0.18 (0.021 to 0.25) | **0.021** | 0.059 | 0.73 | 0.61 | -0.11 (0.0011 to 0.21) | **0.048** | 1 | 0.066 (-0.3 to 0.16) | 0.95 | 0.98 |
| **PC O-17:0;0/17:0;0** | 0.54 | 0.5 | -0.039 (0.0047 to 0.12) | **0.022** | 0.059 | 0.56 | 0.54 | -0.014 (-0.064 to 0.055) | 0.79 | 1 | 0.064 (-0.18 to 0.012) | 0.096 | 0.34 |
| **PC 18:0;0_22:4;0** | 2.4 | 2.2 | -0.22 (0.035 to 0.54) | **0.023** | 0.063 | 2.4 | 2.4 | 0.006 (-0.16 to 0.17) | 0.64 | 1 | 0.32 (-0.65 to -0.07) | **0.02** | 0.22 |
| **PE 18:1;0_18:2;0** | 0.95 | 0.77 | -0.19 (0.028 to 0.36) | **0.023** | 0.063 | 0.94 | 1 | 0.071 (-0.21 to 0.079) | 0.48 | 1 | 0.26 (-0.51 to 0.024) | 0.096 | 0.34 |
| **PE 18:0;0_18:2;0** | 10.8 | 9.7 | -1.1 (0.15 to 1.8) | **0.023** | 0.063 | 11.1 | 10.7 | -0.44 (-0.44 to 1.4) | 0.35 | 1 | 0.66 (-2.2 to 0.97) | 0.43 | 0.63 |
| **PC O-18:1;0/16:0;0** | 1.6 | 1.5 | -0.16 (0.02 to 0.24) | **0.025** | 0.066 | 1.6 | 1.6 | 0.0041 (-0.16 to 0.19) | 0.56 | 1 | 0.17 (-0.3 to 0.094) | 0.49 | 0.67 |
| **PC 14:0;0_18:2;0** | 16.5 | 14.1 | -2.4 (0.49 to 4.5) | **0.026** | 0.068 | 16.6 | 15.8 | -0.78 (-1 to 3) | 0.37 | 1 | 1.7 (-4.3 to 0.49) | 0.088 | 0.33 |
| **TAG 56:5;0** | 8.2 | 9.2 | 1.1 (-1.9 to -0.068) | **0.026** | 0.068 | 8.7 | 8.3 | -0.43 (-0.88 to 1.8) | 0.43 | 1 | -0.91 (-0.63 to 2.7) | 0.16 | 0.37 |

| | | | | | | | | | | | | | |
|---|---|---|---|---|---|---|---|---|---|---|---|---|---|
| **PI 18:1;0_18:2;0** | 0.57 | 0.48 | -0.097 (0.011 to 0.21) | **0.026** | 0.068 | 0.56 | 0.56 | -7.7e-05 (-0.065 to 0.093) | 0.67 | 1 | 0.13 (-0.3 to 0.06) | 0.21 | 0.43 |
| **Cer 42:0;2** | 0.33 | 0.28 | -0.041 (0.0042 to 0.091) | **0.027** | 0.07 | 0.33 | 0.33 | -0.004 (-0.028 to 0.045) | 0.75 | 1 | 0.054 (-0.1 to 0.013) | 0.18 | 0.39 |
| **PC 15:0;0_18:2;0** | 16.5 | 14.5 | -2 (0.13 to 3) | **0.028** | 0.071 | 16.1 | 16.2 | 0.088 (-1.7 to 1.5) | 0.61 | 1 | 2 (-4.4 to 0.9) | 0.21 | 0.42 |
| **PC 17:1;0_18:1;0** | 2.3 | 2.1 | -0.29 (0.02 to 0.45) | **0.029** | 0.073 | 2.3 | 2.4 | 0.084 (-0.19 to 0.12) | 0.67 | 1 | 0.24 (-0.58 to 0.12) | 0.21 | 0.43 |
| **PC O-18:1;0/18:2;0** | 2.5 | 2.1 | -0.32 (0.045 to 0.61) | **0.03** | 0.075 | 2.5 | 2.4 | -0.1 (-0.32 to 0.38) | 0.56 | 1 | 0.37 (-0.85 to 0.015) | 0.06 | 0.32 |
| **PC O-16:0;0/20:5;0** | 0.71 | 0.58 | -0.13 (0.015 to 0.28) | **0.03** | 0.075 | 0.69 | 0.75 | 0.058 (-0.16 to 0.15) | 0.89 | 1 | 0.15 (-0.46 to 0.15) | 0.26 | 0.48 |
| **PC 18:1;0_20:1;0** | 0.65 | 0.59 | -0.055 (0.0073 to 0.18) | **0.031** | 0.076 | 0.63 | 0.67 | 0.043 (-0.11 to 0.082) | 0.66 | 1 | 0.16 (-0.34 to -0.0094) | **0.033** | 0.24 |
| **PC 16:0;0_20:5;0** | 24.6 | 21 | -3.6 (0.25 to 6.8) | **0.031** | 0.076 | 23.7 | 25.2 | 1.5 (-4.8 to 2.8) | 0.79 | 1 | 4.8 (-11 to 0.91) | 0.088 | 0.33 |
| **PC O-16:1;0/16:0;0** | 4.3 | 3.8 | -0.42 (0.025 to 0.69) | **0.033** | 0.079 | 4.1 | 4.2 | 0.06 (-0.35 to 0.42) | 0.82 | 1 | 0.48 (-0.99 to 0.073) | 0.08 | 0.33 |
| **PE 16:0;0_18:2;0** | 2 | 1.8 | -0.18 (0.022 to 0.4) | **0.036** | 0.087 | 2.1 | 2 | -0.12 (-0.14 to 0.32) | 0.56 | 1 | 0.067 (-0.43 to 0.23) | 0.58 | 0.72 |
| **PC 16:0;0_19:1;0** | 1.2 | 1.1 | -0.12 (0.013 to 0.25) | **0.037** | 0.087 | 1.3 | 1.3 | -0.01 (-0.16 to 0.14) | 0.99 | 1 | 0.12 (-0.31 to 0.092) | 0.26 | 0.48 |

| | | | | | | | | | | | | | |
|---|---|---|---|---|---|---|---|---|---|---|---|---|---|
| **TAG 50:5;0** | 3 | 2.4 | -0.61 (0.047 to 1.1) | **0.038** | 0.09 | 2.6 | 3.2 | 0.62 (-0.66 to 0.43) | 0.85 | 1 | 1.5 (-1.6 to -0.07) | **0.02** | 0.22 |
| **CE 18:0;0** | 18.6 | 16.4 | -2.2 (0.13 to 4.1) | **0.038** | 0.09 | 19.7 | 19.5 | -0.24 (-2 to 2) | 0.93 | 1 | 2 (-4.6 to 0.91) | 0.18 | 0.39 |
| **PC 17:0;0_22:4;0** | 1.6 | 1.4 | -0.22 (0.0076 to 0.35) | **0.039** | 0.09 | 1.4 | 1.6 | 0.21 (-0.55 to 0.06) | 0.14 | 1 | 0.51 (-1.1 to 0.11) | 0.12 | 0.36 |
| **PC 16:0;0_22:6;0** | 62.6 | 68.9 | 6.2 (-11 to -0.42) | **0.04** | 0.093 | 64.6 | 68.7 | 4.1 (-13 to 4.8) | 0.46 | 1 | -2.1 (-8 to 12) | 0.58 | 0.72 |
| **PC O-16:0;0/22:4;0** | 0.61 | 0.56 | -0.05 (0.0031 to 0.11) | **0.041** | 0.093 | 0.61 | 0.58 | -0.031 (-0.068 to 0.11) | 0.61 | 1 | 0.028 (-0.15 to 0.096) | 0.69 | 0.79 |
| **PC 17:1;0_18:0;0** | 0.64 | 0.54 | -0.098 (0.0047 to 0.16) | **0.043** | 0.097 | 0.64 | 0.65 | 0.011 (-0.12 to 0.059) | 0.43 | 1 | 0.14 (-0.23 to 0.013) | 0.064 | 0.33 |
| **TAG 52:3;0** | 202.8 | 223.1 | 20 (-49 to -1.6) | **0.043** | 0.096 | 222.4 | 218.1 | -4.2 (-26 to 37) | 0.69 | 1 | -25 (-7.8 to 71) | 0.11 | 0.35 |
| **PE O-18:0;0/20:4;0** | 0.6 | 0.52 | -0.082 (0.0033 to 0.19) | **0.043** | 0.096 | 0.63 | 0.56 | -0.069 (-0.022 to 0.13) | 0.21 | 1 | 0.079 (-0.21 to 0.076) | 0.37 | 0.57 |
| **Cer 40:2;2** | 0.37 | 0.36 | -0.012 (0.00091 to 0.056) | **0.044** | 0.097 | 0.35 | 0.38 | 0.025 (-0.04 to 0.0092) | 0.27 | 1 | 0.042 (-0.083 to -0.016) | **0.012** | 0.18 |
| **PC 17:0;0_20:6;0** | 2.3 | 2.2 | -0.081 (0.01 to 0.56) | **0.045** | 0.098 | 2.3 | 2.2 | -0.043 (-0.36 to 0.34) | 0.95 | 1 | 0.41 (-0.85 to 0.14) | 0.15 | 0.37 |
| **PC 14:0;0_16:0;0** | 3.6 | 3.1 | -0.52 (0.0027 to 1.3) | **0.046** | 0.099 | 3.8 | 3.8 | 0.00088 (-0.61 to 0.41) | 0.66 | 1 | 0.68 (-1.5 to 0.12) | 0.076 | 0.33 |
| **CE 20:2;0** | 5.8 | 5.2 | -0.57 (0.11 to 1.4) | **0.046** | 0.099 | 7.2 | 5.7 | -1.5 (-0.66 to 1.1) | 0.62 | 1 | 0.15 (-2.1 to 0.4) | 0.14 | 0.37 |
| **SM 34:0;2** | 3.7 | 3.4 | -0.38 (0.0063 to 0.68) | **0.047** | 0.1 | 3.7 | 4 | 0.22 (-0.53 to 0.11) | 0.16 | 1 | 0.61 (-1.1 to -0.036) | **0.038** | 0.25 |

| | | | | | | | | | | | | | |
|---|---|---|---|---|---|---|---|---|---|---|---|---|---|
| **PC O-18:2;0/18:1;0** | 0.62 | 0.55 | -0.074 (-0.0039 to 0.16) | 0.06 | 0.12 | 0.61 | 0.61 | 0.008 (-0.095 to 0.079) | 0.95 | 1 | 0.11 (-0.22 to -0.0054) | **0.034** | 0.24 |
| **TAG 52:6;0** | 5.7 | 4.2 | -1.5 (-0.084 to 1.6) | 0.063 | 0.13 | 4.3 | 5.6 | 1.2 (-1 to 0.63) | 0.71 | 1 | 3 (-2.4 to -0.016) | **0.045** | 0.28 |
| **LPE 20:1;0** | 0.39 | 0.31 | -0.074 (-0.004 to 0.13) | 0.08 | 0.15 | 0.33 | 0.36 | 0.032 (-0.079 to 0.032) | 0.54 | 1 | 0.16 (-0.25 to -0.069) | **0.0024** | 0.17 |
| **PC O-17:1;0/17:0;0** | 0.12 | 0.1 | -0.013 (-0.0015 to 0.027) | 0.088 | 0.16 | 0.11 | 0.11 | -0.0052 (-0.005 to 0.01) | 0.59 | 1 | 0.011 (-0.029 to -0.00036) | **0.048** | 0.29 |
| **PE O-16:2;0/18:0;0** | 0.074 | 0.062 | -0.012 (-0.0023 to 0.021) | 0.12 | 0.2 | 0.079 | 0.07 | -0.0097 (4e-04 to 0.02) | **0.041** | 0.97 | 0.00017 (-0.022 to 0.018) | 0.95 | 0.97 |
| **LPE 16:0;0** | 0.71 | 0.63 | -0.076 (-0.012 to 0.13) | 0.13 | 0.21 | 0.69 | 0.72 | 0.027 (-0.1 to 0.057) | 0.69 | 1 | 0.12 (-0.24 to -0.017) | **0.029** | 0.24 |
| **PC O-18:0;0/20:6;0** | 1.1 | 1 | -0.079 (-0.098 to 0.24) | 0.36 | 0.46 | 1.2 | 1.1 | -0.1 (0.0026 to 0.3) | **0.042** | 0.97 | -0.02 (-0.27 to 0.31) | 0.89 | 0.94 |
| **PC 17:0;0_20:4;0** | 7.8 | 7.9 | 0.12 (-0.82 to 0.62) | 0.63 | 0.72 | 7.9 | 8.4 | 0.51 (-1.3 to -0.084) | **0.018** | 0.85 | 0.41 (-1.3 to 0.58) | 0.56 | 0.71 |
| **SM 34:1;3** | 1 | 1 | 0.031 (-0.12 to 0.16) | 0.78 | 0.83 | 0.89 | 1.1 | 0.16 (-0.3 to -0.084) | **0.0013** | 0.39 | 0.26 (-0.56 to 0.017) | 0.067 | 0.33 |
| **PC 18:1;0_20:4;0** | 19.9 | 19.6 | -0.34 (-1.6 to 1.9) | 1 | 1 | 20.1 | 21.7 | 1.6 (-2.5 to -0.068) | **0.039** | 0.97 | 1.9 (-3.6 to 1.1) | 0.37 | 0.57 |

Lipid species showing significant changes within eTRE, lTRE, and/or between interventions are shown. Data are shown as mean or mean (95% CI).

a) P-values without correction for multiple testing.

b) P-values with correction for multiple testing.

**Table S6.** Lipid classes showing alterations within or between eTRE and lTRE interventions.

| Lipid classes | Before eTRE | After eTRE | Change eTRE after – before (95% CI) | P[a)] | P BH[b)] | Before lTRE | After lTRE | Change lTRE After – before (95% CI) | P[a)] | P BH[b)] | Difference between lTRE vs. eTRE (95% CI)[a] | P[a)] | P BH[b)] |
|---|---|---|---|---|---|---|---|---|---|---|---|---|---|
| Cer | 8.5 | 7.6 | -0.88 (0.26 to 1.4) | **0.0054** | **0.043** | 8.3 | 8.2 | -0.11 (-0.39 to 0.75) | 0.3 | 0.99 | 0.78 (-1.5 to 0.43) | 0.31 | 0.62 |
| PC | 1894.6 | 1711.8 | -180 (43 to 260) | **0.0062** | **0.043** | 1911.7 | 1909.9 | -1.8 (-110 to 140) | 0.61 | 0.99 | 180 (-380 to 94) | 0.48 | 0.67 |
| PE O- | 52.3 | 44.4 | -7.9 (2.1 to 12) | **0.013** | 0.06 | 52.8 | 50.5 | -2.2 (-3.2 to 7.2) | 0.58 | 0.99 | 6.1 (-14 to 4.3) | 0.29 | 0.62 |
| CE | 4870 | 4453.8 | -420 (76 to 740) | **0.021** | 0.061 | 4940.1 | 5004.1 | 64 (-420 to 390) | 0.9 | 0.99 | 480 (-1200 to 280) | 0.3 | 0.62 |
| LPC | 105 | 90 | -15 (1.4 to 18) | **0.023** | 0.061 | 107.3 | 106.5 | -0.77 (-11 to 13) | 0.93 | 0.99 | 16 (-28 to -0.67) | 0.04 | 0.28 |
| LPE | 8.1 | 6.9 | -1.3 (0.13 to 1.9) | **0.026** | 0.061 | 7.9 | 8 | 0.12 (-0.94 to 0.88) | 0.99 | 0.99 | 1.5 (-2.4 to -0.27) | **0.015** | 0.22 |
| PC O- | 96.2 | 86.6 | -9.6 (0.81 to 18) | **0.036** | 0.073 | 97.6 | 98.3 | 0.74 (-10 to 9.2) | 0.96 | 0.99 | 11 (-24 to 2.3) | 0.1 | 0.47 |

Data are shown as mean or mean (95% CI).

a) P-values without correction for multiple testing.

b) P-values with correction for multiple testing.

**Table S7.** Fatty acids showing alterations within or between eTRE and lTRE interventions.

| Fatty acid | Before eTRE | After eTRE | Change eTRE after – before (95% CI) | P a) | P BH b) | Before lTRE | After lTRE | Change lTRE After – before (95% CI) | P a) | P BH b) | Difference between lTRE vs. | P a) | P BH b) |
|---|---|---|---|---|---|---|---|---|---|---|---|---|---|
| 18:0;0 | 633.2 | 550.2 | -83 (35 to 120) | **0.00021** | **0.010** | 638.3 | 632.5 | -5.8 (-32 to 54) | 0.62 | 1 | 78 (-140 to 8.6) | 0.088 | 0.51 |
| 20:3;0 | 206.8 | 176.5 | -30 (14 to 43) | **0.00061** | **0.012** | 211.9 | 199.9 | -12 (-1.9 to 32) | 0.079 | 1 | 18 (-41 to 7.6) | 0.16 | 0.51 |
| 20:2;0 | 30.9 | 26.4 | -4.5 (2.3 to 6.9) | **0.00073** | **0.012** | 32.1 | 29.6 | -2.5 (-1.3 to 6.1) | 0.17 | 1 | 2 (-6.3 to 2.8) | 0.4 | 0.64 |
| O-17:2;0 | 0.16 | 0.13 | -0.024 (0.0088 to 0.039) | **0.0029** | **0.026** | 0.16 | 0.16 | -0.0056 (-0.01 to 0.026) | 0.32 | 1 | 0.012 (-0.037 to 0.015) | 0.37 | 0.63 |
| 18:3;0 | 242.4 | 182.4 | -60 (14 to 76) | **0.003** | **0.026** | 227 | 240.7 | 14 (-21 to 35) | 0.98 | 1 | 74 (-90 to 13) | 0.16 | 0.51 |
| O-16:2;0 | 3.4 | 2.7 | -0.74 (0.22 to 1.1) | **0.0032** | **0.026** | 3.2 | 3.4 | 0.18 (-0.42 to 0.36) | 0.96 | 1 | 0.98 (-1.5 to -0.22) | **0.005** | 0.22 |
| 20:0;0 | 2 | 1.7 | -0.33 (0.099 to 0.6) | **0.008** | 0.05 | 1.9 | 2.2 | 0.35 (-0.47 to 0.23) | 0.57 | 1 | 0.81 (-1 to -0.11) | **0.018** | 0.39 |
| O-18:2;0 | 19.4 | 16.2 | -3.3 (0.77 to 4.5) | **0.0087** | 0.05 | 19.5 | 18.6 | -0.82 (-1.5 to 3.1) | 0.48 | 1 | 2.6 (-5.4 to 1.4) | 0.16 | 0.51 |
| O-18:1;0 | 38.6 | 34 | -4.6 (0.73 to 7.3) | **0.0093** | 0.05 | 39.3 | 38.8 | -0.44 (-3.3 to 3.7) | 0.9 | 1 | 4.6 (-9.9 to 1.6) | 0.2 | 0.51 |
| 14:0;0 | 152.2 | 123.8 | -28 (4.9 to 49) | **0.016** | 0.079 | 156.2 | 156 | -0.26 (-8.9 to 24) | 0.43 | 1 | 28 (-46 to -0.85) | **0.043** | 0.51 |
| O-17:0;0 | 1.8 | 1.5 | -0.28 (0.033 to 0.49) | **0.023** | 0.1 | 1.8 | 1.8 | 0.035 (-0.24 to 0.12) | 0.54 | 1 | 0.33 (-0.65 to 0.029) | 0.061 | 0.51 |
| 14:1;0 | 10.2 | 7.6 | -2.6 (0.2 to 4.4) | **0.029** | 0.12 | 10.7 | 10.6 | -0.12 (-1.6 to 2.5) | 0.66 | 1 | 2.5 (-5.1 to 1.1) | 0.3 | 0.6 |
| 18:2;0 | 4238.2 | 3891.9 | -350 (22 to 580) | **0.035** | 0.12 | 4390.6 | 4268.4 | -120 (-240 to 470) | 0.5 | 1 | 220 (-840 to 490) | 0.75 | 0.94 |
| O-16:0;0 | 30.8 | 27.5 | -3.3 (0.33 to 6.3) | **0.035** | 0.12 | 31.7 | 31.6 | -0.12 (-3.5 to 3.5) | 0.66 | 1 | 3.5 (-8.1 to 0.94) | 0.1 | 0.51 |

| | | | | | | | | | | | | | |
|---|---|---|---|---|---|---|---|---|---|---|---|---|---|
| 20:5;0 | 148.4 | 127.5 | -21 (1.4 to 41) | **0.043** | 0.13 | 150.8 | 159.3 | 8.6 (-32 to 17) | 0.72 | 1 | 29 (-66 to 6.6) | 0.088 | 0.51 |
| O-16:1;0 | 42 | 37.4 | -4.6 (0.28 to 8.3) | **0.043** | 0.13 | 42 | 41.5 | -0.48 (-3.6 to 4.6) | 0.81 | 1 | 4.4 (-12 to 3) | 0.33 | 0.6 |
| O-17:1;0 | 0.12 | 0.1 | -0.013 (-0.0015 to 0.027) | 0.088 | 0.2 | 0.11 | 0.11 | -0.0052 (-0.005 to 0.01) | 0.59 | 1 | 0.011 (-0.029 to -0.00036) | **0.048** | 0.51 |

Data are shown as mean or mean (95% CI).

[a)] P-values without correction for multiple testing.

[b)] P-values with correction for multiple testing.

**Table S8.** Indices of enzyme activity and their calculation.

| Index | Description | Calculation |
|---|---|---|
| D5D/FADS1 | Delta-5 Desaturase | Sum 20:4 FA* / 20:3 FA* |
| D6D/FADS2 | Delta-6 Desaturase | Sum 18:3 FA* / 18:2 FA* |
| D9D/SCD1 (C16) | Delta-9 Desaturase / Stearoyl-CoA Desaturase 1 - C16 | Sum 16:1 FA* / 16:0 FA* |
| D9D/SCD1 (C18) | Delta-9 Desaturase / Stearoyl-CoA Desaturase 1 - C18 | Sum 18:1 FA* / 18:0 FA* |
| D9D/SCD1 (C18+16) | Delta-9 Desaturase / Stearoyl-CoA Desaturase 1 - C18+16 | Sum (16:1 + 18:1) FA* / (16:0 + 18:0) FA* |
| ELOVL5 | Elongation of Very Long Chain Fatty Acids Protein 5 | Sum 20:3 FA* / 18:3 FA* |
| ELOVL6 | Elongation of Very Long Chain Fatty Acids Protein 6 | Sum 18:0 FA* / 16:0 FA * |

* for all lipids in which fatty acids (FA) were measured

**Table S9.** Changes of enzyme activity indices within or between eTRE and lTRE interventions.

| Indices | Before eTRE | After eTRE | Change eTRE after – before (95% CI) | P[a)] | P BH[b)] | Before lTRE | After lTRE | Change lTRE After – before (95% CI) | P[a)] | P BH[b)] | Difference between lTRE vs. eTRE (95% CI)[a] | P[a)] | P BH[b)] |
|---|---|---|---|---|---|---|---|---|---|---|---|---|---|
| ELOVL6 | 0.26 | 0.23 | -0.025 (0.016 to 0.033) | **0.000006** | **0.000096** | 0.25 | 0.25 | -0.0012 (-0.0097 to 0.016) | 0.62 | 0.82 | 0.023 (-0.034 to -0.0071) | **0.0054** | 0.062 |
| D5D | 4.7 | 5.6 | 0.85 (-1.2 to -0.37) | **0.00034** | **0.0024** | 4.7 | 5.5 | 0.82 (-1.1 to -0.41) | **0.000045** | **0.00073** | -0.0062 (-0.51 to 0.56) | 0.92 | 0.94 |
| SCD1/D9D (C18) | 4.2 | 4.6 | 0.42 (-0.68 to -0.17) | **0.0016** | **0.0073** | 4.3 | 4.3 | 0.0012 (-0.37 to 0.26) | 0.65 | 0.82 | -0.42 (0.031 to 0.83) | 0.026 | 0.15 |
| D6D | 0.056 | 0.047 | -0.009 (0.0013 to 0.011) | **0.015** | **0.036** | 0.052 | 0.054 | 0.0014 (-0.0031 to 0.0042) | 0.62 | 0.82 | 0.01 (-0.011 to 0.00087) | 0.14 | 0.4 |
| ELOVL5 | 0.96 | 1 | 0.076 (-0.18 to 0.011) | 0.088 | 0.14 | 0.97 | 0.96 | -0.015 (-0.061 to 0.082) | 0.53 | 0.82 | -0.087 (-0.053 to 0.2) | 0.21 | 0.46 |
| SCD1/D9D (C18+16) | 0.97 | 0.98 | 0.013 (-0.055 to 0.02) | 0.34 | 0.43 | 0.97 | 0.98 | 0.0063 (-0.048 to 0.026) | 0.53 | 0.82 | -0.0086 (-0.038 to 0.06) | 0.7 | 0.85 |
| SCD1/D9D (C16) | 0.16 | 0.15 | -0.0037 (-0.0072 to 0.014) | 0.53 | 0.62 | 0.16 | 0.16 | 0.0028 (-0.0097 to 0.0047) | 0.49 | 0.82 | 0.0066 (-0.018 to 0.0058) | 0.3 | 0.53 |

Data are shown as mean or mean (95% CI).

a) P-values without correction for multiple testing.

b) P-values with correction for multiple testing.

**Table S10.** Total length of carbon chain per lipid class showing alterations within or between eTRE and lTRE interventions.

| **Total length of carbon chain per** | **Before eTRE** | **After eTRE** | **Change eTRE after – before (95% CI)** | **P[a)]** | **P BH[b)]** | **Before lTRE** | **After lTRE** | **Change lTRE After – before (95% CI)** | **P[a)]** | **P BH[b)]** | **Difference between lTRE vs. eTRE (95% CI)[a]** | **P[a)]** | **P BH[b)]** |
|---|---|---|---|---|---|---|---|---|---|---|---|---|---|
| PC_C_36 | 35.4 | 34.5 | -0.87 (0.48 to 1.2) | **0.00003** | **0.00095** | 35.3 | 35.1 | -0.26 (-0.15 to 0.6) | 0.29 | 0.49 | 0.59 (-1.1 to -0.22) | **0.015** | 0.19 |
| PI_C_38 | 64.8 | 68.4 | 3.6 (-5.2 to -2.3) | **0.00003** | **0.00095** | 65.7 | 67.3 | 1.6 (-2.9 to -0.26) | **0.021** | 0.19 | -2 (-0.045 to 4.3) | 0.058 | 0.4 |
| LPC_C_18 | 45.9 | 43.7 | -2.2 (1.1 to 3.1) | **0.000044** | **0.00095** | 45.4 | 45.5 | 0.11 (-1.2 to 1) | 0.99 | 0.99 | 2.2 (-3.5 to -0.74) | **0.0032** | 0.052 |
| PI_C_36 | 25.3 | 22.8 | -2.5 (1.5 to 3.6) | **0.000079** | **0.0013** | 24.7 | 23.6 | -1 (0.059 to 2.1) | **0.041** | 0.28 | 1.5 (-3.4 to 0.092) | 0.067 | 0.4 |
| SM_C_40 | 17.6 | 16.8 | -0.74 (0.33 to 1.1) | **0.00011** | **0.0014** | 17.2 | 17 | -0.19 (-0.21 to 0.61) | 0.36 | 0.54 | 0.53 (-1.1 to 0.033) | 0.073 | 0.4 |
| TAG_C_52 | 47.3 | 50.4 | 3.1 (-4.6 to -1.7) | **0.00014** | **0.0015** | 47.1 | 47.8 | 0.72 (-2 to 0.74) | 0.38 | 0.55 | -2.4 (0.29 to 4.2) | **0.029** | 0.29 |
| LPC_C_16 | 49.5 | 51.9 | 2.4 (-3.5 to -1.1) | **0.00019** | **0.0018** | 50.1 | 49.5 | -0.53 (-0.67 to 1.5) | 0.46 | 0.63 | -2.8 (1 to 4.4) | **0.0013** | **0.044** |
| SM_C_36 | 10.2 | 10.9 | 0.69 (-1.1 to -0.34) | **0.00026** | **0.0021** | 10.5 | 10.4 | -0.15 (-0.16 to 0.47) | 0.23 | 0.43 | -0.84 (0.3 to 1.3) | **0.00019** | **0.012** |
| PI_C_34 | 8 | 6.9 | -1.1 (0.43 to 1.6) | **0.00067** | **0.0048** | 7.8 | 7.4 | -0.39 (-0.17 to 0.91) | 0.12 | 0.31 | 0.72 (-1.5 to -0.0097) | **0.047** | 0.38 |
| CE_C_22 | 0.89 | 0.99 | 0.096 (-0.14 to -0.041) | **0.0012** | **0.008** | 0.9 | 0.95 | 0.053 (-0.11 to 0.00064) | 0.055 | 0.31 | -0.04 (-0.035 to 0.087) | 0.28 | 0.61 |
| SM_C_32 | 4 | 3.8 | -0.26 (0.11 to 0.41) | **0.0019** | **0.011** | 3.9 | 3.9 | -0.09 (-0.023 to 0.2) | 0.098 | 0.31 | 0.17 (-0.37 to 0.031) | 0.096 | 0.42 |
| PC_C_38 | 15.5 | 16.3 | 0.76 (-1.1 to -0.3) | **0.0026** | **0.013** | 15.6 | 16.1 | 0.52 (-0.89 to 0.051) | 0.094 | 0.31 | -0.27 (-0.5 to 0.86) | 0.57 | 0.82 |
| SM_C_38 | 7.5 | 7.3 | -0.24 (0.086 to 0.41) | **0.0028** | **0.013** | 7.4 | 7.3 | -0.083 (-0.1 to 0.22) | 0.52 | 0.69 | 0.15 (-0.44 to 0.073) | 0.13 | 0.43 |

| | | | | | | | | | | | | | |
|---|---|---|---|---|---|---|---|---|---|---|---|---|---|
| TAG_C_48 | 5.4 | 4.2 | -1.3 (0.43 to 2) | **0.0028** | **0.013** | 5.7 | 5.1 | -0.62 (-0.12 to 1.1) | 0.1 | 0.31 | 0.67 (-1.7 to 0.66) | 0.5 | 0.78 |
| PC O-_C_34 | 24.3 | 23.1 | -1.2 (0.34 to 1.8) | **0.003** | **0.013** | 23.8 | 23.4 | -0.38 (-0.26 to 1.1) | 0.3 | 0.49 | 0.8 (-1.7 to 0.072) | 0.1 | 0.42 |
| CE_C_20 | 12.1 | 12.9 | 0.88 (-1.2 to -0.23) | **0.0035** | **0.014** | 12.3 | 13.2 | 0.93 (-1.4 to -0.33) | **0.0014** | **0.029** | 0.075 (-0.85 to 0.83) | 0.78 | 0.93 |
| PE O-_C_34 | 5 | 4.4 | -0.57 (0.17 to 0.83) | **0.0052** | **0.02** | 4.6 | 4.4 | -0.26 (-0.11 to 0.57) | 0.22 | 0.43 | 0.39 (-0.94 to 0.26) | 0.35 | 0.64 |
| Cer_C_40 | 24.6 | 23.6 | -1 (0.23 to 1.4) | **0.0076** | **0.026** | 24 | 24.2 | 0.14 (-1.1 to 1) | 0.88 | 0.94 | 1.1 (-2.1 to 0.44) | 0.14 | 0.43 |
| Cer_C_42 | 75.4 | 76.4 | 1 (-1.4 to -0.23) | **0.0076** | **0.026** | 76 | 75.8 | -0.14 (-1 to 1.1) | 0.88 | 0.94 | -1.1 (-0.44 to 2.1) | 0.14 | 0.43 |
| LPE_C_22 | 9.2 | 10.2 | 1 (-1.7 to -0.2) | **0.013** | **0.042** | 8.9 | 9.2 | 0.34 (-1.1 to 0.56) | 0.53 | 0.69 | -0.75 (-0.55 to 1.9) | 0.38 | 0.65 |
| TAG_C_46 | 1.3 | 0.84 | -0.49 (0.082 to 0.78) | **0.014** | **0.043** | 1.5 | 1.2 | -0.22 (-0.03 to 0.45) | 0.079 | 0.31 | 0.26 (-0.62 to 0.38) | 0.72 | 0.88 |
| PC O-_C_38 | 26.2 | 27.1 | 0.94 (-1.9 to -0.29) | **0.015** | **0.043** | 26.2 | 26.3 | 0.14 (-0.92 to 0.64) | 0.64 | 0.77 | -0.81 (-0.4 to 2) | 0.1 | 0.42 |
| TAG_C_49 | 0.9 | 0.75 | -0.15 (0.025 to 0.23) | **0.02** | 0.056 | 0.94 | 0.83 | -0.11 (0.0034 to 0.2) | **0.039** | 0.28 | 0.041 (-0.19 to 0.21) | 0.92 | 0.98 |
| PE_C_34 | 6.2 | 5.8 | -0.38 (0.042 to 0.57) | **0.025** | 0.067 | 6.5 | 6.1 | -0.32 (0.012 to 0.62) | **0.043** | 0.28 | 0.061 (-0.36 to 0.45) | 0.9 | 0.98 |
| PE O-_C_38 | 55.4 | 56.9 | 1.5 (-2.1 to -0.14) | **0.028** | 0.072 | 56.3 | 56.3 | -0.056 (-0.92 to 0.66) | 0.82 | 0.91 | -1.5 (-0.46 to 2.7) | 0.18 | 0.46 |
| PE_C_36 | 36.9 | 35.4 | -1.5 (0.093 to 2.4) | **0.029** | 0.073 | 37.1 | 36.2 | -0.93 (-0.55 to 2.4) | 0.19 | 0.41 | 0.61 (-2.2 to 1.5) | 0.72 | 0.88 |
| PE_C_38 | 46.1 | 47.5 | 1.4 (-2.8 to -0.22) | **0.038** | 0.092 | 45.3 | 46.3 | 0.93 (-2.1 to 0.34) | 0.12 | 0.31 | -0.55 (-1.1 to 2.3) | 0.5 | 0.78 |
| CE_C_18 | 75.5 | 74.7 | -0.87 (-0.07 to 1.4) | 0.096 | 0.22 | 75.2 | 74.1 | -1.2 (0.45 to 1.6) | **0.00076** | **0.029** | -0.36 (-0.58 to 1.4) | 0.38 | 0.65 |

| | | | | | | | | | | | | | |
|---|---|---|---|---|---|---|---|---|---|---|---|---|---|
| PC_C_37 | 0.76 | 0.81 | 0.045 (-0.1 to 0.0074) | 0.11 | 0.23 | 0.75 | 0.81 | 0.056 (-0.094 to -0.017) | **0.0058** | 0.095 | 0.011 (-0.066 to 0.074) | 0.87 | 0.96 |
| LPC_C_20 | 4.7 | 4.6 | -0.1 (-0.14 to 0.44) | 0.27 | 0.43 | 4.5 | 4.9 | 0.42 (-0.69 to -0.16) | **0.0014** | **0.029** | 0.57 (-0.91 to -0.25) | **0.0029** | 0.052 |
| PC_C_35 | 1.9 | 1.9 | 0.016 (-0.075 to 0.03) | 0.4 | 0.58 | 1.8 | 1.9 | 0.14 (-0.18 to -0.013) | **0.014** | 0.15 | 0.12 (-0.21 to -0.013) | **0.031** | 0.29 |
| SM_C_42 | 23.1 | 23.3 | 0.24 (-0.61 to 0.24) | 0.48 | 0.64 | 23.1 | 23.6 | 0.46 (-0.82 to -0.14) | **0.0097** | 0.13 | 0.19 (-0.86 to 0.29) | 0.21 | 0.48 |

Data are shown as mean or mean (95% CI).

Lipid classes & total length of carbon chain are shown as, for example, “PC_C_36” meaning all measured phosphatidylcholines with a total carbon chain length of 36 atoms.

[a)] P-values without correction for multiple testing.

[b)] P-values with correction for multiple testing.

**Table S11.** Total double bonds per lipid class showing alterations within and between eTRE and/or lTRE interventions.

| Total double bonds per lipid class | Before eTRE | After eTRE | Change eTRE after – before (95% CI) | P[a)] | P BH[b)] | Before lTRE | After lTRE | Change lTRE After – before (95% CI) | P[a)] | P BH[b)] | Difference between lTRE vs. eTRE | P[a)] | P BH[b)] |
|---|---|---|---|---|---|---|---|---|---|---|---|---|---|
| PC_db_4 | 17.5 | 19.1 | 1.6 (-2.2 to -1.1) | **0.000006** | **0.00022** | 17.8 | 18.7 | 0.93 (-1.3 to -0.42) | **0.00045** | **0.011** | -0.66 (-0.097 to 1.4) | 0.08 | 0.34 |
| PI_db_4 | 61.3 | 65 | 3.7 (-5.3 to -2.6) | **0.000008** | **0.00022** | 61.7 | 63.8 | 2.1 (-3.3 to -0.92) | **0.0015** | **0.021** | -1.6 (-0.18 to 3.5) | 0.07 | 0.33 |
| PE_db_3 | 7.4 | 6.4 | -0.98 (0.65 to 1.4) | **0.0000092** | **0.00022** | 7.1 | 6.8 | -0.28 (-0.16 to 0.7) | 0.27 | 0.5 | 0.68 (-1.2 to -0.25) | **0.0099** | 0.13 |
| PC_db_6 | 6 | 6.7 | 0.76 (-1 to -0.38) | **0.000063** | **0.0011** | 6 | 6.3 | 0.32 (-0.7 to 0.15) | 0.23 | 0.46 | -0.45 (-0.22 to 0.89) | 0.17 | 0.47 |
| CE_db_4 | 9.3 | 10.3 | 1 (-1.4 to -0.54) | **0.00012** | **0.0015** | 9.4 | 10.3 | 0.88 (-1.2 to -0.48) | **0.00012** | **0.0042** | -0.12 (-0.49 to 0.73) | 0.69 | 0.78 |
| LPE_db_4 | 11.9 | 13.3 | 1.5 (-1.8 to -0.63) | **0.00012** | **0.0015** | 12.3 | 12.2 | -0.013 (-0.56 to 0.51) | 0.87 | 0.92 | -1.3 (0.42 to 2) | **0.0013** | **0.045** |
| CE_db_3 | 3.4 | 2.9 | -0.45 (0.2 to 0.66) | **0.00023** | **0.0023** | 3.4 | 3.2 | -0.23 (0.0021 to 0.4) | **0.043** | 0.23 | 0.21 (-0.52 to 0.14) | 0.24 | 0.52 |
| PC_db_3 | 10.5 | 9.8 | -0.77 (0.33 to 1.2) | **0.00034** | **0.0027** | 10.4 | 9.9 | -0.5 (0.14 to 0.8) | **0.0071** | 0.056 | 0.25 (-0.83 to 0.26) | 0.3 | 0.53 |
| PI_db_1 | 10.5 | 9.1 | -1.4 (0.6 to 2.1) | **0.00038** | **0.0027** | 10.5 | 9.9 | -0.52 (-0.19 to 1.3) | 0.16 | 0.4 | 0.92 (-2 to 0.043) | 0.058 | 0.33 |
| LPE_db_2 | 21.1 | 18.7 | -2.4 (1.2 to 3.6) | **0.00042** | **0.0027** | 20.7 | 20 | -0.63 (-0.43 to 1.7) | 0.36 | 0.58 | 1.8 (-3.4 to -0.018) | 0.045 | 0.33 |
| PI_db_3 | 8.9 | 8 | -0.9 (0.46 to 1.4) | **0.00042** | **0.0027** | 9 | 8.4 | -0.59 (0.28 to 0.95) | **0.0012** | **0.021** | 0.28 (-0.9 to 0.34) | 0.43 | 0.54 |
| Cer_db_2 | 30.7 | 32.8 | 2.1 (-2.9 to -0.72) | **0.00055** | **0.0031** | 31.3 | 31.7 | 0.43 (-1.5 to 0.61) | 0.4 | 0.6 | -1.7 (0.43 to 3) | 0.014 | 0.14 |
| PE O-_db_5 | 47.2 | 50 | 2.8 (-4.1 to -0.99) | **0.00061** | **0.0031** | 48.8 | 49.4 | 0.67 (-2.8 to 1.4) | 0.56 | 0.7 | -2.1 (-0.86 to 4.8) | 0.21 | 0.49 |
| SM_db_2 | 35.5 | 36.3 | 0.84 (-1.2 to -0.35) | **0.00061** | **0.0031** | 35.6 | 36.2 | 0.56 (-0.94 to -0.22) | **0.0051** | 0.052 | -0.31 (-0.42 to 0.98) | 0.35 | 0.53 |

| | | | | | | | | | | | | | |
|---|---|---|---|---|---|---|---|---|---|---|---|---|---|
| PE_db_6 | 13.5 | 16.4 | 2.9 (-4.2 to -1.6) | **0.00074** | **0.0031** | 13.8 | 14.6 | 0.78 (-2.7 to 0.93) | 0.41 | 0.6 | -1.8 (-0.66 to 4.3) | 0.18 | 0.47 |
| TAG_db_0 | 0.83 | 0.48 | -0.34 (0.12 to 0.39) | **0.00075** | **0.0031** | 0.84 | 0.75 | -0.085 (-0.019 to 0.29) | 0.095 | 0.29 | 0.15 (-0.36 to 0.2) | 0.6 | 0.7 |
| LPE_db_0 | 45.9 | 48.9 | 3 (-3.7 top -0.96) | **0.0008** | **0.0031** | 46.7 | 46.3 | -0.36 (-0.85 to 1.8) | 0.49 | 0.68 | -3.3 (0.78 to 4.4) | **0.0058** | 0.13 |
| PE_db_2 | 29.5 | 27.1 | -2.4 (1 to 3.8) | **0.0008** | **0.0031** | 29.9 | 28.2 | -1.7 (0.22 to 2.5) | **0.022** | 0.14 | 0.66 (-2.6 to 1.1) | 0.31 | 0.53 |
| LPC_db_0 | 65.2 | 67.7 | 2.6 (-3.7 to -0.89) | **0.00087** | **0.0033** | 66.3 | 65.1 | -1.2 (-0.11 to 2.2) | 0.076 | 0.26 | -3.8 (1.7 to 5.4) | **0.00014** | **0.0098** |
| SM_db_1 | 63.3 | 62.5 | -0.77 (0.33 to 1.2) | **0.0011** | **0.004** | 63.2 | 62.6 | -0.58 (0.22 to 0.96) | **0.002** | **0.024** | 0.23 (-0.93 to 0.45) | 0.43 | 0.54 |
| CE_db_6 | 0.89 | 0.99 | 0.096 (-0.14 to -0.041) | **0.0012** | **0.0041** | 0.9 | 0.95 | 0.053 (-0.11 to 0.00064) | 0.055 | 0.23 | -0.04 (-0.035 to 0.087) | 0.28 | 0.53 |
| LPC_db_1 | 14 | 13.2 | -0.87 (0.34 to 1.4) | **0.0013** | **0.0041** | 13.6 | 13.8 | 0.28 (-0.59 to 0.33) | 0.64 | 0.75 | 1.2 (-1.9 to -0.3) | **0.011** | 0.13 |
| PI_db_2 | 15 | 13.6 | -1.3 (0.72 to 2.2) | **0.0013** | **0.0041** | 14.5 | 13.6 | -0.93 (-0.022 to 1.7) | 0.058 | 0.23 | 0.44 (-1.7 to 0.82) | 0.42 | 0.54 |
| LPE_db_1 | 12 | 10 | -1.9 (0.64 to 3) | **0.002** | **0.0057** | 11.5 | 12.2 | 0.67 (-1.7 to 0.5) | 0.25 | 0.46 | 2.5 (-4.5 to -0.69) | **0.016** | 0.14 |
| PC O-_db_2 | 14.4 | 13.2 | -1.2 (0.31 to 1.8) | **0.002** | **0.0057** | 14 | 14 | -0.019 (-0.6 to 0.88) | 0.57 | 0.7 | 1.2 (-1.9 to 0.039) | 0.07 | 0.33 |
| LPC_db_2 | 16.3 | 14.7 | -1.6 (0.47 to 2.3) | **0.0024** | **0.0065** | 15.6 | 16.2 | 0.52 (-1.2 to 0.5) | 0.41 | 0.6 | 2.1 (-3.5 to -0.41) | **0.0099** | 0.13 |
| PC O-_db_3 | 12.5 | 11.6 | -0.85 (0.25 to 1.3) | **0.003** | **0.0079** | 12.3 | 11.9 | -0.42 (-0.016 to 0.8) | 0.061 | 0.23 | 0.46 (-1.1 to 0.21) | 0.18 | 0.47 |
| PE O-_db_3 | 9.5 | 8.5 | -1 (0.3 to 1.4) | **0.0043** | **0.011** | 9.1 | 8.6 | -0.51 (-0.18 to 1) | 0.25 | 0.46 | 0.47 (-1.4 to 0.63) | 0.33 | 0.53 |
| PC O-_db_5 | 24 | 25.4 | 1.4 (-2.5 to -0.48) | **0.0058** | **0.014** | 23.8 | 24.6 | 0.79 (-1.9 to 0.3) | 0.1 | 0.3 | -0.61 (-0.98 to 2.4) | 0.45 | 0.56 |
| PE O-_db_4 | 6.4 | 5.8 | -0.65 (0.18 to 1) | **0.0065** | **0.015** | 6.3 | 6.1 | -0.16 (-0.47 to 0.75) | 0.58 | 0.7 | 0.55 (-1.4 to 0.24) | 0.21 | 0.49 |
| TAG_db_3 | 29.4 | 30.9 | 1.4 (-2.4 to -0.35) | **0.0076** | **0.017** | 29.1 | 29.6 | 0.53 (-1.2 to 0.16) | 0.11 | 0.32 | -0.93 (-0.58 to 2.2) | 0.29 | 0.53 |

| | | | | | | | | | | | | | |
|---|---|---|---|---|---|---|---|---|---|---|---|---|---|
| LPC_db_4 | 3.5 | 3.8 | 0.24 (-0.44 to -0.051) | **0.012** | **0.026** | 3.5 | 3.9 | 0.4 (-0.54 to -0.2) | **0.0000092** | **0.00065** | 0.12 (-0.36 to 0.15) | 0.34 | 0.53 |
| LPE_db_6 | 9.2 | 10.2 | 1 (-1.7 to -0.2) | **0.013** | **0.028** | 8.9 | 9.2 | 0.34 (-1.1 to 0.56) | 0.53 | 0.7 | -0.75 (-0.55 to 1.9) | 0.38 | 0.54 |
| PC_db_2 | 42.1 | 40.7 | -1.3 (0.37 to 2.5) | **0.015** | **0.029** | 41.9 | 40.9 | -1 (0.006 to 1.6) | **0.046** | 0.23 | 0.27 (-1.8 to 1.3) | 0.97 | 0.97 |
| PE_db_4 | 31.2 | 33 | 1.8 (-3.2 to -0.42) | **0.015** | **0.029** | 31.7 | 32.4 | 0.62 (-2.6 to 0.72) | 0.22 | 0.44 | -1.1 (-0.96 to 3.4) | 0.22 | 0.49 |
| DAG_db_4 | 6.4 | 5.4 | -1 (0.17 to 1.7) | **0.016** | **0.032** | 6.1 | 6.8 | 0.7 (-1.5 to 0.72) | 0.95 | 0.98 | 1.6 (-2.7 to 0.25) | 0.097 | 0.36 |
| Cer_db_1 | 63.1 | 61.7 | -1.4 (0.18 to 2.6) | **0.017** | **0.033** | 62.4 | 62.2 | -0.2 (-0.47 to 1.4) | 0.29 | 0.51 | 1.2 (-2.8 to 0.65) | 0.2 | 0.48 |
| TAG_db_1 | 9.5 | 8.1 | -1.4 (0.22 to 2.3) | **0.017** | **0.033** | 9.9 | 9 | -0.9 (-0.069 to 1.8) | 0.063 | 0.23 | 0.49 (-1.9 to 1.4) | 0.86 | 0.88 |
| PE O-_db_2 | 3.7 | 3.4 | -0.38 (0.032 to 0.55) | **0.025** | **0.045** | 3.6 | 3.4 | -0.23 (0.0047 to 0.48) | 0.05 | 0.23 | 0.18 (-0.63 to 0.31) | 0.43 | 0.54 |
| Cer_db_0 | 6.6 | 6.1 | -0.51 (0.036 to 1.6) | **0.039** | 0.07 | 6.7 | 6.7 | -0.016 (-0.86 to 0.9) | 0.97 | 0.99 | 0.5 (-1.9 to 0.4) | 0.14 | 0.46 |
| SM_db_0 | 1.2 | 1.1 | -0.063 (0.0031 to 0.12) | **0.043** | 0.074 | 1.2 | 1.2 | 0.02 (-0.096 to 0.032) | 0.32 | 0.54 | 0.083 (-0.16 to -0.00029) | 0.05 | 0.33 |
| CE_db_2 | 56.3 | 55.9 | -0.32 (-0.81 to 1.2) | 0.67 | 0.7 | 55.9 | 54.7 | -1.2 (0.37 to 2) | **0.0066** | 0.056 | -0.9 (-0.56 to 2.5) | 0.18 | 0.47 |
| TAG_db_9 | 0.41 | 0.32 | -0.094 (-0.015 to 0.15) | 0.11 | 0.18 | 0.29 | 0.4 | 0.11 (-0.21 to -0.027) | **0.014** | 0.099 | 0.16 (-0.29 to 0.005) | 0.074 | 0.33 |

Data are shown as mean or mean (95% CI).

Lipid classes & total double bonds per class are shown as, for example, “PC_db_4” meaning all measured phosphatidylcholines with 4 double bonds.

[a)] P-values without correction for multiple testing.

[b)] P-values with correction for multiple testing.

**Table S13.** Genes which mRNA expression was analyzed by the real-time PCR and primer sequences.

| Gene symbol | Gene ID | Full name | Role | Circadian (tissue) | Forward primer | Reverse primer |
|---|---|---|---|---|---|---|
| FADS1/D5D | 3998 | Fatty acid desaturase 1 | Desaturase | Yes[1] (liver) | 5´-TGCCTTCAATGACTGGTTCA-3´ | 5´- ATGCTTGGCACACAAGGACT-3´ |
| FADS2/D6D | 9415 | Fatty acid desaturase 2 | Desaturase | Yes[1] (liver) | 5´-AAGGGTGCCTCTGCCAACT-3´ | 5´-GATTGTAGGGCAGGTATTTCAGC-3´ |
| SCD1/D9D | 6319 | Stearoyl-CoA desaturase | Desaturase | Yes[2] (liver) | 5´-CATAACAGCAGGAGCTCATCGT-3´ | 5´-ACGAGCCCATTCATAGACATCA-3´ |
| ELOVL5 | 60481 | ELOVL fatty acid elongase 5 | Elongase | Yes[1] (liver) | 5´-TAACAGGAGTATGGGAAGGCA-3´ | 5´-ACGAGCCCATTCATAGACATCA-3´ |
| ELOVL6 | 79071 | ELOVL fatty acid elongase 6 | Elongase | Yes[3] (liver) | 5´-AACGAGCAAAGTTTGAACTGAGG-3´ | 5´-TCGAAGAGCACCGAATATACTGA-3´ |
| FASN | 2194 | Fatty acid synthase | FA synthesis | Yes[4, 5] (adipose tissue and liver) | 5´-AGACACTCGTGGCCTACAGCAT-3´ | 5´-ATGGCCTGGTAGGCGTTCT-3´ |
| ACACA/ACC1 | 31 | Acetyl-CoA carboxylase alpha | FA synthesis | Yes[4, 5] (adipose tissue and liver) | 5´-TCGCTTTGGGGGAAATAAAGTG-3´ | 5´-TCGCTTTGGGGGAAATAAAGTG-3´, |

| | | | | | | |
|---|---|---|---|---|---|---|
| ACSL1 | 2180 | Acyl-CoA synthetase long chain family member 1 | FA synthesis | Yes[6] (adipose tissue) | 5´-AACAGACGGAAGCCCAAGC-3’ | 5´-TCGGTGAGTGACCATTGCTC-3´ |
| CHPT1 | 56994 | Choline phosphotransferase 1 | Biosynthesis of PC | ye [7] (muscle) | 5’-TCCAGTTCTTGGATTTCTAGGTGGAGT-3’ | 5´-ACACTGGTGCCTGCTATAGTGGA-3´ |
| CEPT1 | 10390 | Choline/ethanolamine phosphotransferase 1 | Biosynthesis of PC | yes [7] (muscle) | 5´-CAGTGATTGGAGGACCACCT-3´ | 5´-AGGACACTTGTTCCTGCTATTGT-3´ |
| SPTLC2 | 9517 | Serine palmitoyltransferase long chain base subunit 2 | Biosynthesis of Cer | yes[7] (muscle) | 5´-GAGACGCCTGAAAGAGATGG-3´ | 5´-TGGTATGAGCTGCTGACAGG-3´ |
| CERS6 | 253782 | Ceramide synthase 6 | Biosynthesis of Cer | No[8] (estimated activity, serum) | 5´-CGGACCTGAAGAACACGGAGGA-3´ | 5´-ATGGCGCACGGTTTGGCTAC-3´ |
| GUSB | 2990 | Glucuronidase beta | Housekeeper | | 5´-CTCATTTGGAATTTTGCCGATT-3´ | 5´-CCGAGTGAAGATCCCCTTTTTA-3´ |
| RPLP0 | 6175 | Ribosomal protein lateral stalk subunit P0 | Housekeeper | | 5´-GCTTCCTGGAGGGTGTCC-3´ | 5´-GGACTCGTTTGTACCCGTTG-3´ |

**Supplemental References to the Table S13.**

1. Chen, M., Y. Lin, Y. Dang, Y. Xiao, F. Zhang, G. Sun, X. Jiang, L. Zhang, J. Du, S. Duan, *et al.* "Reprogramming of rhythmic liver metabolism by intestinal clock." *J Hepatol* 79 (2023): 741-57. 10.1016/j.jhep.2023.04.040.
2. Zhou, X., D. Wan, Y. Zhang, Y. Zhang, C. Long, S. Chen, L. He, B. Tan, X. Wu and Y. Yin. "Diurnal variations in polyunsaturated fatty acid contents and expression of genes involved in their de novo synthesis in pigs." *Biochem Biophys Res Commun* 483 (2017): 430-34. 10.1016/j.bbrc.2016.12.126.
3. Chou, C. F., X. Zhu, Y. Y. Lin, K. L. Gamble, W. T. Garvey and C. Y. Chen. "Ksrp is critical in governing hepatic lipid metabolism through controlling per2 expression." *J Lipid Res* 56 (2015): 227-40. 10.1194/jlr.M050724.
4. Kohsaka, A., A. D. Laposky, K. M. Ramsey, C. Estrada, C. Joshu, Y. Kobayashi, F. W. Turek and J. Bass. "High-fat diet disrupts behavioral and molecular circadian rhythms in mice." *Cell Metab* 6 (2007): 414-21. 10.1016/j.cmet.2007.09.006.
5. Kudo, T., T. Tamagawa, M. Kawashima, N. Mito and S. Shibata. "Attenuating effect of clock mutation on triglyceride contents in the icr mouse liver under a high-fat diet." *J Biol Rhythms* 22 (2007): 312-23. 10.1177/0748730407302625.
6. Stenvers, D. J., A. Jongejan, S. Atiqi, J. P. Vreijling, E. J. Limonard, E. Endert, F. Baas, P. D. Moerland, E. Fliers, A. Kalsbeek, *et al.* "Diurnal rhythms in the white adipose tissue transcriptome are disturbed in obese individuals with type 2 diabetes compared with lean control individuals." *Diabetologia* 62 (2019): 704-16. 10.1007/s00125-019-4813-5.
7. Loizides-Mangold, U., L. Perrin, B. Vandereycken, J. A. Betts, J. P. Walhin, I. Templeman, S. Chanon, B. D. Weger, C. Durand, M. Robert, *et al.* "Lipidomics reveals diurnal lipid oscillations in human skeletal muscle persisting in cellular myotubes cultured in vitro." *Proc Natl Acad Sci U S A* 114 (2017): E8565-e74. 10.1073/pnas.1705821114.
8. Poolman, T. M., J. Gibbs, A. L. Walker, S. Dickson, L. Farrell, J. Hensman, A. C. Kendall, R. Maidstone, S. Warwood, A. Loudon, *et al.* "Rheumatoid arthritis reprograms circadian output pathways." *Arthritis Res Ther* 21 (2019): 47. 10.1186/s13075-019-1825-y.
9. Peters, B., D. A. Koppold-Liebscher, B. Schuppelius, N. Steckhan, A. F. H. Pfeiffer, A. Kramer, A. Michalsen and O. Pivovarova-Ramich. "Effects of early vs. Late time-restricted eating on cardiometabolic health, inflammation, and sleep in overweight and obese women: A study protocol for the chronofast trial." *Front Nutr* 8 (2021): 765543. 10.3389/fnut.2021.765543. https://www.ncbi.nlm.nih.gov/pubmed/34869534.
10. Peters, B., J. Schwarz, B. Schuppelius, A. Ottawa, D. A. Koppold, D. Weber, N. Steckhan, K. Mai, T. Grune, A. F. H. Pfeiffer, *et al.* "Effects of isocaloric early vs. Late time-restricted eating on insulin sensitivity, cardiometabolic health, and internal circadian time in women with overweight or obesity." *medRxiv* (2024): 2024.10.05.24314120. 10.1101/2024.10.05.24314120. https://www.medrxiv.org/content/medrxiv/early/2024/10/07/2024.10.05.24314120.full.pdf.

**Table S14.** Lipid pathway enrichment analysis of the plasma lipidome changes within eTRE intervention.

| **Pathway name** | **Pathway lipids** | **P-value[a)]** | **P-value[b)]** | **Signif.** |
|---|---|---|---|---|
| Glycerophospholipid metabolism | 26 | 0.0000137 | 0.00041 | *** |
| Sphingolipid signaling pathway | 9 | 0.0032 | 0.0290 | N.S. |
| Glycosylphosphatidylinositol (GPI)-anchor biosynthesis | 3 | 0.0039 | 0.0290 | N.S. |
| Autophagy - other | 3 | 0.0039 | 0.0290 | N.S. |
| Ferroptosis | 11 | 0.0060 | 0.0324 | N.S. |
| Autophagy - animal | 4 | 0.0076 | 0.0324 | N.S. |
| Necroptosis | 4 | 0.0076 | 0.0324 | N.S. |
| Choline metabolism in cancer | 5 | 0.012 | 0.046 | N.S. |
| Retrograde endocannabinoid signaling | 8 | 0.032 | 0.104 | N.S. |
| Pathogenic Escherichia coli infection | 1 | 0.037 | 0.104 | N.S. |
| Sphingolipid metabolism | 21 | 0.038 | 0.104 | N.S. |
| AGE-RAGE signaling pathway in diabetic complications | 2 | 0.073 | 0.183 | N.S. |
| Neurotrophin signaling pathway | 3 | 0.108 | 0.216 | N.S. |
| Adipocytokine signaling pathway | 3 | 0.108 | 0.216 | N.S. |
| Kaposi's sarcoma-associated herpesvirus infection | 3 | 0.108 | 0.216 | N.S. |
| Ether lipid metabolism | 16 | 0.116 | 0.217 | N.S. |
| Insulin resistance | 4 | 0.141 | 0.235 | N.S. |
| Leishmaniasis | 4 | 0.141 | 0.235 | N.S. |
| Tuberculosis | 5 | 0.173 | 0.274 | N.S. |
| Fat digestion and absorption | 8 | 0.263 | 0.376 | N.S. |
| Cholesterol metabolism | 8 | 0.263 | 0.376 | N.S. |
| Inositol phosphate metabolism | 9 | 0.291 | 0.397 | N.S. |
| Phosphatidylinositol signaling system | 11 | 0.344 | 0.449 | N.S. |
| Vitamin digestion and absorption | 15 | 0.438 | 0.548 | N.S. |
| Ovarian steroidogenesis | 18 | 0.501 | 0.601 | N.S. |
| alpha-Linolenic acid metabolism | 23 | 0.590 | 0.666 | N.S. |
| Linoleic acid metabolism | 25 | 0.621 | 0.666 | N.S. |

| | | | | |
|---|---|---|---|---|
| Bile secretion | 25 | 0.621 | 0.666 | N.S. |
| Steroid biosynthesis | 41 | 0.802 | 0.829 | N.S. |
| Arachidonic acid metabolism | 75 | 0.953 | 0.953 | N.S. |

Lipid pathway enrichment analysis was conducted using LIPEA software as described in *Experimental Section/Methods*.

[a] P-values without correction for multiple testing.

[b] P-values with Bonferroni correction for multiple testing.

**Table S15.** Key resources table.

| *Reagent or Resource* | *Source* | *Identifier* |
|---|---|---|
| **Biological samples** | | |
| Plasma and adipose tissue samples from women with overweight or obesity (the ChronoFast cohort) | German Institute of Human Nutrition Potsdam-Rebruecke [9, 10] | Clinicaltrial.gov Identifier: NCT04351672 |
| **Chemicals, peptides, and recombinant proteins** | | |
| RNase-free Water | MP Biomedicals llc. | Cat.Nr: 7732-18-5 |
| High-Capacity cDNA Reverse Transcription Kit | ThermoFisher Scientific / Applied Biosystems™ | Cat.Nr: 4368814 |
| RNase-Inhibitor | ThermoFisher Scientific / Applied Biosystems™ | Cat.Nr: N8080119 |
| 384-Well Multiply®-PCR Plate | ThermoFisher Scientific / Applied Biosystems™ | Cat.Nr: AB1384 |
| *Power*SYBR® Green PCR Master Mix | ThermoFisher Scientific / Applied Biosystems™ | Cat.Nr: 4367659 |
| qRCR Primers | Invitrogen | N/A |
| **Deposited data** | | |
| Raw RNAseq data for adipose tissue samples | | GSE287198 |
| **Software and algorithms** | | |
| MinimPy | Open source software | https://sourceforge.net/projects/minimpy/ |
| SPSS Statistics 25.0 | IBM | https://www.ibm.com/de-de/products/spss-statistics |
| GraphPad Prism Version 5.0 | GraphPad | https://www.graphpad.com/ |
| Bodygram™ | AKERN | https://www.akern.com/en/products-and-solutions/data-analysis-software/bodygram-software/ |
| Fddb App and Food Database | Food Database GmbH | https://fddb.info/ |
| ActiLife Version 6.13.4 | ActiGraph | https://theactigraph.com/ |

| | | |
|---|---|---|
| QuantStudio™ Real-Time PCR Software | ThermoFisher Scientific | https://www.thermofisher.com/de/de/home/global/forms/life-science/quantstudio-6-7-flex-software.html |
| FastQC v0.12.1 | Open source http://www.bioinformatics.babraham.ac.uk/projects/fastqc/ | RRID:SCR_014583 |
| STAR v2.7.11a | Open source https://github.com/alexdobin/STAR | RRID:SCR_004463 |
| STRINGTIE v2.2.1 | Open source https://ccb.jhu.edu/software/stringtie/ | RRID:SCR_016323 |
| DESeq2 v1.34.0 | Open source https://bioconductor.org/packages/release/bioc/html/DESeq2.html | RRID:SCR_015687 |
| Python v3.10.9 | Open source https://www.python.org/ | RRID:SCR_008394 |
| R v4.1.2 | Open source https://www.r-project.org/ | RRID:SCR_001905 |
| LIPEA | Biomedical Cybernetics Group https://hyperlipea.org/ | N/A |

**A**

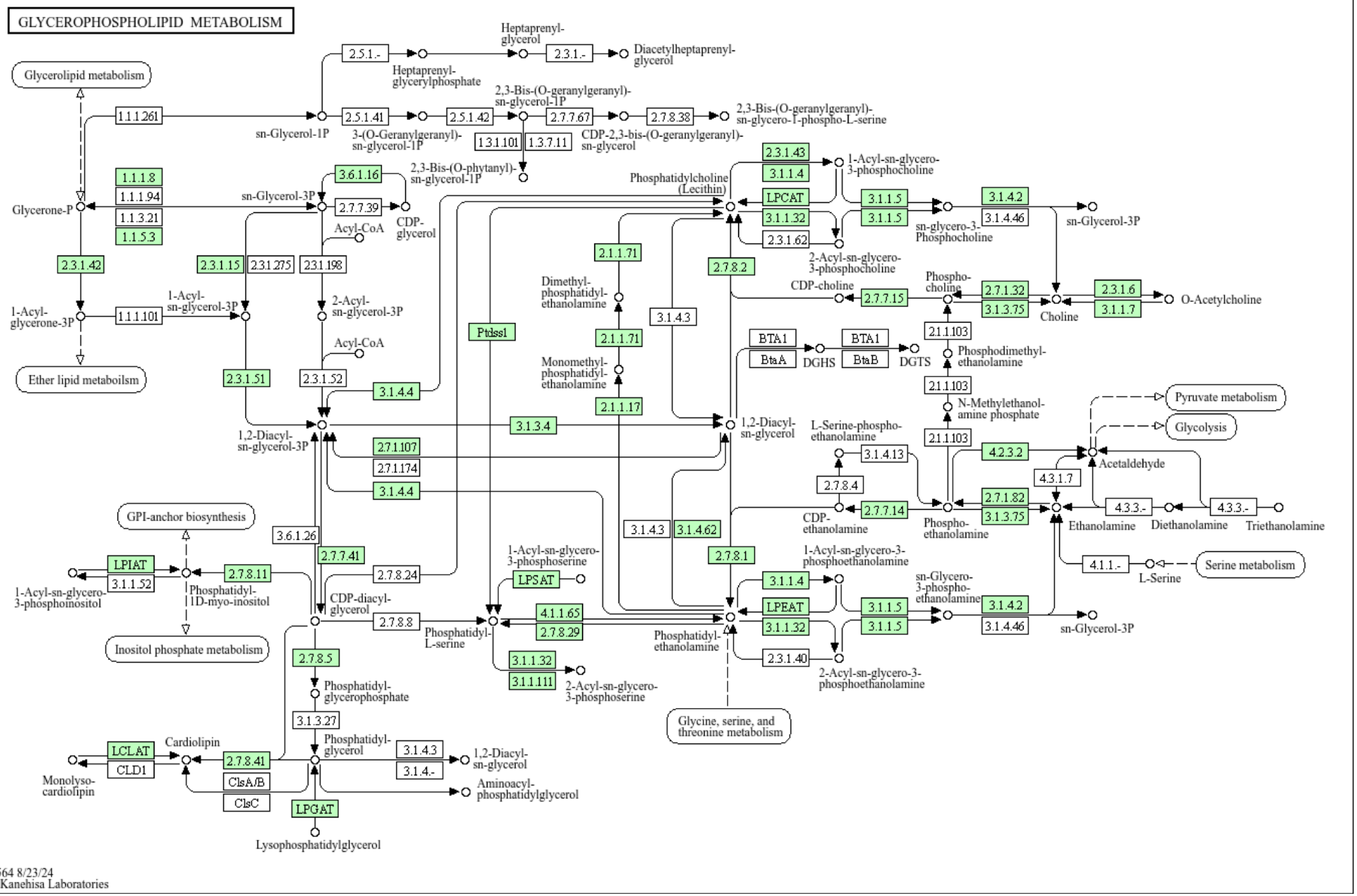

GLYCEROPHOSPHOLIPID METABOLISM
Glycerolipid metabolism
Ether lipid metaboilsm
GPI-anchor biosynthesis
Inositol phosphate metabolism
Glycine, serine, and threonine metabolism
Pyruvate metabolism
Glycolysis
Serine metabolism
Heptaprenyl-glycerol
Heptaprenyl-glycerylphosphate
Diacetylheptaprenyl-glycerol
sn-Glycerol-1P
3-(O-Geranylgeranyl)-sn-glycerol-1P
2,3-Bis-(O-geranylgeranyl)-sn-glycerol-1P
CDP-2,3-bis-(O-geranylgeranyl)-sn-glycerol
2,3-Bis-(O-geranylgeranyl)-sn-glycero-1-phospho-L-serine
2,3-Bis-(O-phytanyl)-sn-glycerol-1P
Glycerone-P
sn-Glycerol-3P
CDP-glycerol
Acyl-CoA
1-Acyl-glycerone-3P
1-Acyl-sn-glycerol-3P
2-Acyl-sn-glycerol-3P
1,2-Diacyl-sn-glycerol-3P
Phosphatidylcholine (Lecithin)
1-Acyl-sn-glycero-3-phosphocholine
2-Acyl-sn-glycero-3-phosphocholine
sn-glycero-3-Phosphocholine
sn-Glycerol-3P
CDP-choline
Phospho-choline
Choline
O-Acetylcholine
Dimethyl-phosphatidyl-ethanolamine
Monomethyl-phosphatidyl-ethanolamine
DGHS
DGTS
Phosphodimethyl-ethanolamine
N-Methylethanol-amine phosphate
1,2-Diacyl-sn-glycerol
L-Serine-phospho-ethanolamine
Acetaldehyde
CDP-ethanolamine
Phospho-ethanolamine
Ethanolamine
Diethanolamine
Triethanolamine
L-Serine
1-Acyl-sn-glycero-3-phosphoinositol
Phosphatidyl-1D-myo-inositol
CDP-diacyl-glycerol
1-Acyl-sn-glycero-3-phosphoserine
Phosphatidyl-L-serine
2-Acyl-sn-glycero-3-phosphoserine
Phosphatidyl-ethanolamine
1-Acyl-sn-glycero-3-phosphoethanolamine
sn-Glycero-3-phospho-ethanolamine
2-Acyl-sn-glycero-3-phosphoethanolamine
sn-Glycerol-3P
Phosphatidyl-glycerophosphate
Phosphatidyl-glycerol
Cardiolipin
Monolyso-cardiolipin
1,2-Diacyl-sn-glycerol
Aminoacyl-phosphatidylglycerol
Lysophosphatidylglycerol
2.5.1.-
2.3.1.-
1.1.1.261
2.5.1.41
2.5.1.42
2.7.7.67
2.7.8.38
1.3.1.101
1.3.7.11
1.1.1.8
1.1.1.94
1.1.3.21
1.1.5.3
3.6.1.16
2.7.7.39
2.3.1.42
2.3.1.15
2.3.1.275
2.3.1.198
1.1.1.101
2.3.1.51
2.3.1.52
3.1.4.4
3.1.3.4
2.7.1.107
2.7.1.174
3.1.4.4
2.3.1.43
3.1.1.4
LPCAT
3.1.1.32
2.3.1.62
3.1.1.5
3.1.1.5
3.1.4.2
3.1.4.46
2.1.1.71
2.1.1.71
2.1.1.17
2.7.8.2
Ptdss1
3.1.4.3
2.7.7.15
2.7.1.32
3.1.3.75
2.3.1.6
3.1.1.7
2.1.1.103
2.1.1.103
2.1.1.103
BTA1
BtaA
BTA1
BtaB
3.1.4.13
2.7.8.4
4.2.3.2
4.3.1.7
2.7.1.82
3.1.3.75
2.7.7.14
4.3.3.-
4.3.3.-
4.1.1.-
3.6.1.26
2.7.7.41
3.1.4.3
3.1.4.62
2.7.8.1
LPIAT
3.1.1.52
2.7.8.11
2.7.8.24
LPSAT
3.1.1.4
LPEAT
3.1.1.32
3.1.1.5
3.1.1.5
3.1.4.2
3.1.4.46
2.7.8.8
4.1.1.65
2.7.8.29
3.1.1.32
3.1.1.111
2.3.1.40
2.7.8.5
3.1.3.27
LCLAT
CLD1
2.7.8.41
ClsA/B
ClsC
3.1.4.3
3.1.4.-
LPGAT
00564 8/23/24
(c) Kanehisa Laboratories

B

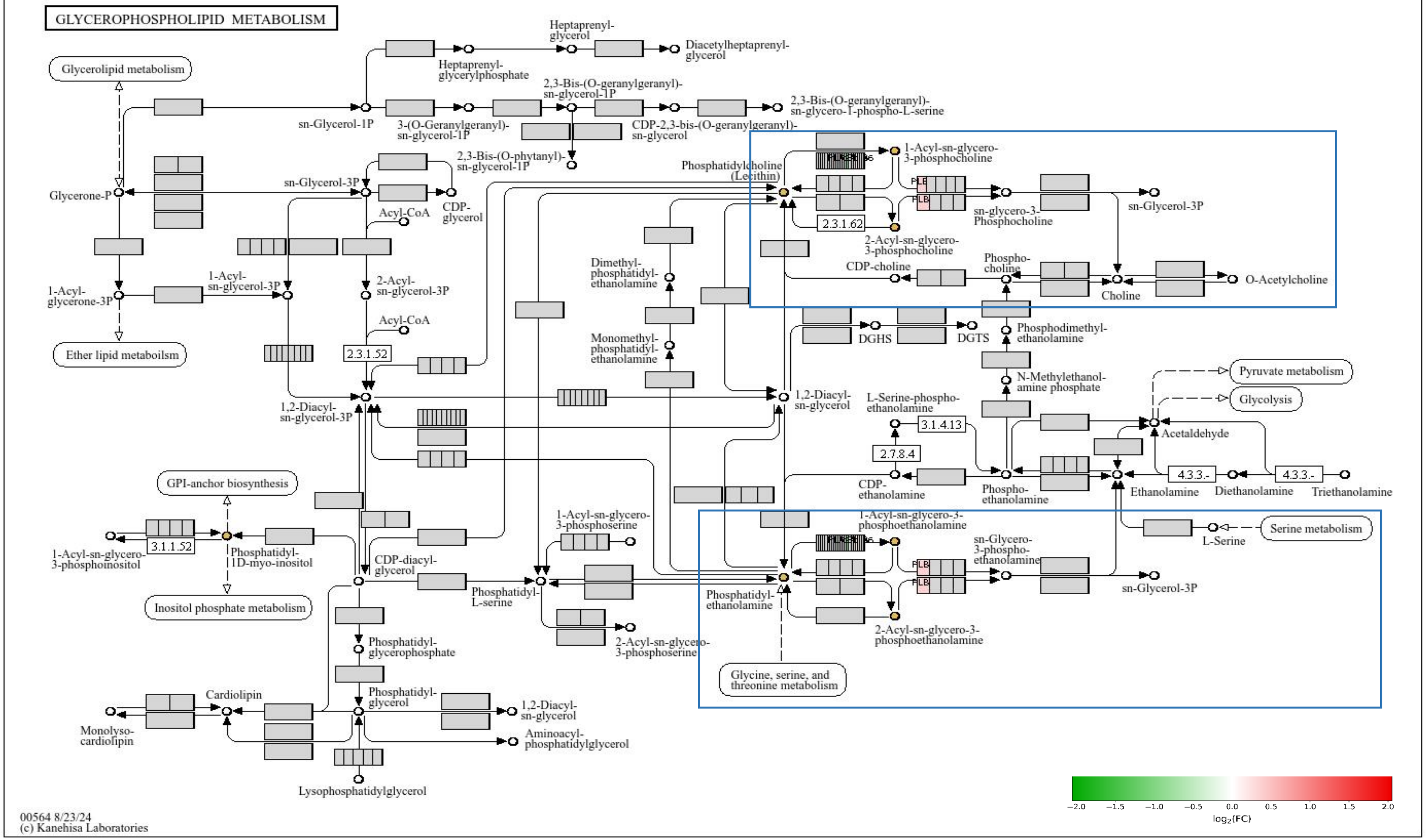

GLYCEROPHOSPHOLIPID METABOLISM
Glycerolipid metabolism
Ether lipid metaboilsm
GPI-anchor biosynthesis
Inositol phosphate metabolism
Glycine, serine, and threonine metabolism
Serine metabolism
Pyruvate metabolism
Glycolysis
Heptaprenyl-glycerol
Diacetylheptaprenyl-glycerol
Heptaprenyl-glycerylphosphate
sn-Glycerol-1P
3-(O-Geranylgeranyl)-sn-glycerol-1P
2,3-Bis-(O-geranylgeranyl)-sn-glycerol-1P
CDP-2,3-bis-(O-geranylgeranyl)-sn-glycerol
2,3-Bis-(O-geranylgeranyl)-sn-glycero-1-phospho-L-serine
2,3-Bis-(O-phytanyl)-sn-glycerol-1P
Glycerone-P
sn-Glycerol-3P
CDP-glycerol
Acyl-CoA
1-Acyl-glycerone-3P
1-Acyl-sn-glycerol-3P
2-Acyl-sn-glycerol-3P
2.3.1.52
1,2-Diacyl-sn-glycerol-3P
Phosphatidylcholine (Lecithin)
1-Acyl-sn-glycero-3-phosphocholine
2-Acyl-sn-glycero-3-phosphocholine
2.3.1.62
sn-glycero-3-Phosphocholine
sn-Glycerol-3P
CDP-choline
Phospho-choline
Choline
O-Acetylcholine
Dimethyl-phosphatidyl-ethanolamine
Monomethyl-phosphatidyl-ethanolamine
DGHS
DGTS
Phosphodimethyl-ethanolamine
N-Methylethanol-amine phosphate
1,2-Diacyl-sn-glycerol
L-Serine-phospho-ethanolamine
3.1.4.13
2.7.8.4
Acetaldehyde
CDP-ethanolamine
Phospho-ethanolamine
Ethanolamine
4.3.3.-
Diethanolamine
Triethanolamine
L-Serine
1-Acyl-sn-glycero-3-phosphoinositol
3.1.1.52
Phosphatidyl-1D-myo-inositol
CDP-diacyl-glycerol
1-Acyl-sn-glycero-3-phosphoserine
Phosphatidyl-L-serine
2-Acyl-sn-glycero-3-phosphoserine
1-Acyl-sn-glycero-3-phosphoethanolamine
sn-Glycero-3-phospho-ethanolamine
sn-Glycerol-3P
Phosphatidyl-ethanolamine
2-Acyl-sn-glycero-3-phosphoethanolamine
Phosphatidyl-glycerophosphate
Phosphatidyl-glycerol
1,2-Diacyl-sn-glycerol
Aminoacyl-phosphatidylglycerol
Cardiolipin
Monolyso-cardiolipin
Lysophosphatidylglycerol
PLB
00564 8/23/24
(c) Kanehisa Laboratories
−2.0 −1.5 −1.0 −0.5 0.0 0.5 1.0 1.5 2.0
log2(FC)

**Figure S1. Full map of the glycerophospholipid pathway combining lipid and gene expression data.**

(A) Glycerophospholipid pathway identified by the lipid pathway enrichment analysis of plasma lipidome before and after the eTRE intervention. Pathway enzymes which activity were potentially altered by this intervention are highlighted in green.

(B) Glycerophospholipid pathway combining lipid and gene expression data. The output of the glycerophospholipid pathway revealed by the lipid pathway enrichment analysis was subjected to the metaKEGG tool together with the SAT transcriptome dataset. Genes found in the transcriptome dataset were directly mapped on the pathway, colored according to their $log_2FC$ translated to a color scale, while KEGG compounds were assigned a single color to highlight their presence in the pathway. In the color scale, red color means a positive log2FC, i.e. the transcript was upregulated in ITRF compared to the eTRE, whereas green color means a negative log2FC. Areas zoomed in Figure3 are highlighted with a blue frame.
